# Brain-charting autism and attention deficit hyperactivity disorder reveals distinct and overlapping neurobiology

**DOI:** 10.1101/2023.12.06.23299587

**Authors:** Saashi A. Bedford, Meng-Chuan Lai, Michael V. Lombardo, Bhismadev Chakrabarti, Amber Ruigrok, John Suckling, Evdokia Anagnostou, Jason P. Lerch, Margot Taylor, Rob Nicolson, Georgiades Stelios, Jennifer Crosbie, Russell Schachar, Elizabeth Kelley, Jessica Jones, Paul D. Arnold, Eric Courchesne, Karen Pierce, Lisa T. Eyler, Kathleen Campbell, Cynthia Carter Barnes, Jakob Seidlitz, Aaron F. Alexander-Bloch, Edward T. Bullmore, Simon Baron-Cohen, Richard A.I. Bethlehem, MRC AIMS Consortium and Lifespan Brain Chart Consortium

**Affiliations:** Autism Research Centre, Department of Psychiatry, University of Cambridge, Cambridge, CB2 8AH, UK; Brain Mapping Unit, Department of Psychiatry, University of Cambridge, Cambridge, CB2 0SZ, UK; The Margaret and Wallace McCain Centre for Child, Youth & Family Mental Health and Azrieli Adult Neurodevelopmental Centre, Campbell Family Mental Health Research Institute, Centre for Addiction and Mental Health, Toronto, ON M6J 1H4, Canada; Department of Psychiatry, The Hospital for Sick Children, Toronto, ON M5G 1X8, Canada; Department of Psychiatry, Temerty Faculty of Medicine, University of Toronto, Toronto, ON M5T 1R8, Canada; Department of Psychiatry, National Taiwan University Hospital and College of Medicine, Taipei 100229, Taiwan; Laboratory for Autism and Neurodevelopmental Disorders, Center for Neuroscience and Cognitive Systems, Istituto Italiano di Tecnologia, Rovereto, Italy; Centre for Autism, School of Psychology and Clinical Language Sciences, University of Reading, Reading RG6 6ES, UK; Division of Psychology and Mental Health, School of Health Sciences, Faculty of Biology, Medicine and Health, University of Manchester; Bloorview Research Institute, Holland Bloorview Kids Rehabilitation Hospital, Toronto, Ontario, Canada; Department of Pediatrics, Temerty Faculty of Medicine, University of Toronto, Toronto, Canada; Program in Neurosciences and Mental Health, Research Institute, Hospital for Sick Children, Toronto, ON M5G 1X8, Canada; Mouse Imaging Centre, Hospital for Sick Children, Toronto, ON M5G 1X8, Canada; Wellcome Centre for Integrative Neuroimaging, FMRIB, Nuffield Department of Clinical Neurosciences, University of Oxford, Oxford OX3 9DU, UK; Department of Diagnostic Imaging, Hospital for Sick Children, Toronto, ON M5G 1X8, Canada; Department of Psychiatry, University of Western Ontario, London, Ontario, Canada; McMaster University, Hamilton, Ontario, Canada; Genetics & Genome Biology, The Hospital for Sick Children, Toronto, Ontario, Canada; Department of Psychology, Queen’s University, Kingston, ON K7L 3N6 Canada; Centre for Neuroscience Studies, Queen’s University, Kingston, ON K7L 3N6 Canada; Department of Psychiatry, Queen’s University, Kingston, ON K7L 3N6 Canada; The Mathison Centre for Mental Health Research & Education, Hotchkiss Brain Institute, Cumming School of Medicine, University of Calgary, Calgary, Alberta, Canada; Departments of Psychiatry and Medical Genetics, Cumming School of Medicine, University of Calgary, Calgary, Alberta, Canada; Department of Neurosciences, University of California San Diego, La Jolla, California, USA; Department of Psychiatry, University of California San Diego, La Jolla, California, USA; Department of Psychiatry, University of Pennsylvania, Philadelphia, PA 19104, USA; Department of Child and Adolescent Psychiatry and Behavioral Science, The Children’s Hospital of Philadelphia, Philadelphia, PA 19104, USA; Lifespan Brain Institute, The Children’s Hospital of Philadelphia and Penn Medicine, Philadelphia, PA 19104, USA; Cambridge Lifetime Autism Spectrum Service (CLASS), Cambridgeshire and Peterborough NHS Foundation Trust, Cambridge, UK; Department of Psychology, University of Cambridge, Cambridge, UK

## Abstract

**Background:** Autism and attention deficit hyperactivity disorder (ADHD) are heterogeneous neurodevelopmental conditions with complex underlying neurobiology. Despite overlapping presentation and sex-biased prevalence, autism and ADHD are rarely studied together, and sex differences are often overlooked. Normative modelling provides a unified framework for studying age-specific and sex-specific divergences in neurodivergent brain development.

**Methods:** Here we use normative modelling and a large, multi-site neuroimaging dataset to characterise cortical anatomy associated with autism and ADHD, benchmarked against models of typical brain development based on a sample of over 75,000 individuals. We also examined sex and age differences, relationship with autistic traits, and explored the co-occurrence of autism and ADHD (autism+ADHD).

**Results:** We observed robust neuroanatomical signatures of both autism and ADHD. Overall, autistic individuals showed greater cortical thickness and volume localised to the superior temporal cortex, whereas individuals with ADHD showed more global effects of cortical thickness increases but lower cortical volume and surface area across much of the cortex. The autism+ADHD group displayed a unique pattern of widespread increases in cortical thickness, and certain decreases in surface area. We also found evidence that sex modulates the neuroanatomy of autism but not ADHD, and an age-by-diagnosis interaction for ADHD only.

**Conclusions:** These results indicate distinct cortical differences in autism and ADHD that are differentially impacted by age, sex, and potentially unique patterns related to their co-occurrence.

## Introduction

Neurodevelopmental conditions such as autism and attention deficit hyperactivity disorder (ADHD) are the products of altered neurodevelopmental trajectories ^1^, but their specific neurobiological underpinnings remain poorly understood. Both autism and ADHD display significant variability in trajectory, associated traits, aetiology and neurobiology ^2–8^, which can hamper efforts to better understand these conditions as a whole. Sex and gender modulations of presentation, prevalence and neuroanatomy ^9–15^, and the high rate of clinical and aetiological overlap between autism and ADHD ^16–19^, add substantial complexity. Importantly, most studies have been based on male-dominant samples and therefore might not be representative or generalisable ^15^.

One of the most commonly reported neuroanatomical findings in autism is of increased total brain volume in young autistic children ^20–22^, although evidence suggests this might only hold true for a subset ^23–25^, and only for boys ^26,27^. Increased cortical thickness is often associated with autism ^28–31^, although reductions have been reported ^32,33^, as well as alterations in cortical surface area and volume ^23,32,34–36^. Cortical alterations, including both increases and decreases, have been reported in the superior temporal gyrus, inferior and prefrontal cortex, and multiple sensory and motor regions ^29–34,37,38^, along with structural and volumetric differences in the cerebellum and subcortex ^39–42^. Importantly, these differences appear to be moderated by age, sex, and co-occurring conditions or traits ^25,30,31,43–48^. Complementary work has suggested that multiple subgroups with distinct patterns of neuroanatomical alterations and clinical and demographic characteristics exist in this multi-dimensional space ^25,40,48–50^. Sex differences, in particular, have been reported in cortical thickness ^31,44^, gyrification ^51^ and mean curvature alterations ^52^, as well as relationships between cortical morphology and autistic traits ^53,54^, with some studies reporting additional regions and greater magnitude of difference in autistic females ^41,55–57^.

Recent meta-analyses have highlighted a similar lack of convergent findings in ADHD ^58,59^. Reduced total brain volume, grey matter volume and cortical surface area in individuals with ADHD have been rather consistently reported ^59–65^. However, while earlier studies reported decreases in cortical thickness ^66–70^, more recent studies with bigger sample sizes have found no or only very minimal differences ^60–64,71–73^. Cortical alterations have most commonly been reported in frontal (primarily dorsolateral and ventrolateral prefrontal, and orbitofrontal), parietal, anterior cingulate and occipital cortices ^59,71^. Volumetric reductions of multiple subcortical structures and cerebellum have also been reported ^59,65,74^, in particular in the basal ganglia ^75,76^, likely related to atypicality in the frontostriatal network ^65,77–79^. As in autism, differences are highly dependent on age, sex, and co-occurring conditions ^59,66,69,80^. Sex-specific alterations have been observed in grey matter volume and surface area in prefrontal and premotor regions in children ^81,82^, as well as reduced pallidum and amygdala volumes in boys, but not girls with ADHD ^83^. Sex differences in white matter microstructure ^84^, caudate volume ^85^, and the anterior cingulate cortex ^86^ have also been reported, while others have found no sex differences ^71,87–89^.

The few studies that have examined structural brain differences in autism and ADHD together have reported largely distinct, with some overlapping, alterations ^90–97^. Importantly, a review by Rommelse and colleagues ^98^ highlights the challenges of comparing these two groups, including limited sample sizes, heterogeneity within conditions, often arbitrary distinctions between clinical groups (e.g. co-occurring ADHD in autistic individuals is often ignored in research studies), and overlap in behavioural presentations. Even fewer studies have specifically examined the co-occurrence of autism and ADHD. Again, both similarities and differences have been observed between those with co-occurring compared to only one diagnosis, at times with more pronounced differences in those with dual diagnoses ^47,95,99^, and evidence that ADHD diagnosis modulates the effect of autism on neuroanatomy ^97^.

While this lack of consistency in the literature is likely due in part to differences in methodology and sample size, another significant contributor is the heterogeneity within, as well as the overlap between, the two conditions. To identify average patterns of alterations, large datasets are needed, as well as techniques to harmonise multi-site data. Critically, these alterations have to be contextualised in light of typical brain development given the neurodevelopmental nature of autism and ADHD, and age must be taken into account ^31,43,46,100–102^.

Normative modelling is a technique that has proven effective for examining and characterising age-dependent variation in brain development ^103,104^, and has recently been employed in studying autism or ADHD ^48,54,96,105^. Normative modelling provides a framework for studying diverse neurodevelopmental conditions in reference to a common baseline, allowing us to better quantify individual differences and address heterogeneity, sex differences and multi-site datasets. Normative modelling also provides a potential route toward clinical applications and translational potential of neuroimaging work ^106^. Similar to the use of paediatric growth charts, by characterising typical brain development across the lifespan, we can identify individually specific alterations or divergence from these trajectories that may be associated with neurodevelopmental conditions, even before associated traits manifest clinically.

Here we leveraged normative models of typical brain development previously characterised by our group ^103^ to quantify divergence in brain development related to autism and ADHD, in comparison to a common reference sample. Furthermore, we examined sources of variability related to sex, age, and continuous measures of autistic and ADHD traits. Finally, we examined a subset of individuals with co-occurring autism and ADHD, to explore whether this group presents with distinct or overlapping neuroanatomical alterations compared to those with only one diagnosis.

## Methods

### Sample and datasets

T1-weighted scans were combined from 49 sites across 7 datasets, including the Autism Brain Imaging Data Exchange (ABIDE ^107,108^), the Province of Ontario Neurodevelopmental (POND) Network, the Healthy Brain Network (HBN) at the Child Mind Institute (CMI) ^109^, the ADHD200 Consortium, the Multimodal Developmental Neurogenetics of Females with ASD (Female ASD) dataset from the NIMH NDAR initiative, the UK Medical Research Council Autism Imaging Multi-centre Study (MRC-AIMS), and the University of California San Diego (UCSD) Biomarkers of Autism study. The final dataset after quality control (QC) included 4255 individuals, (1869 typically developing (TD) controls [1182 male, 687 female], 987 ADHD [717 male, 270 female] and 1399 autistic [1111 male, 288 female], age range 2-64 years [mean 14.0; median 12.4]; Figure 1). For details of each dataset, and participant demographics by dataset before and after QC, see supplementary methods S1. It is important to note the distinction between biological sex and gender identity, which both might influence presentation ^12^. Here we refer to sex assigned at birth, as this was the only data consistently available. We acknowledge the overlap with and influence of gender socialisation, and the lack of data to speak to gender identity effects. Individuals with MRI data available, and a primary diagnosis of autism or ADHD, or no diagnosis, were included. In the main analyses, individuals were included in the group of their primary diagnosis. A subset of individuals with recorded co-occurring autism and ADHD were examined in further analyses (see “Statistical analysis” section for sample details).

**Figure 1.**
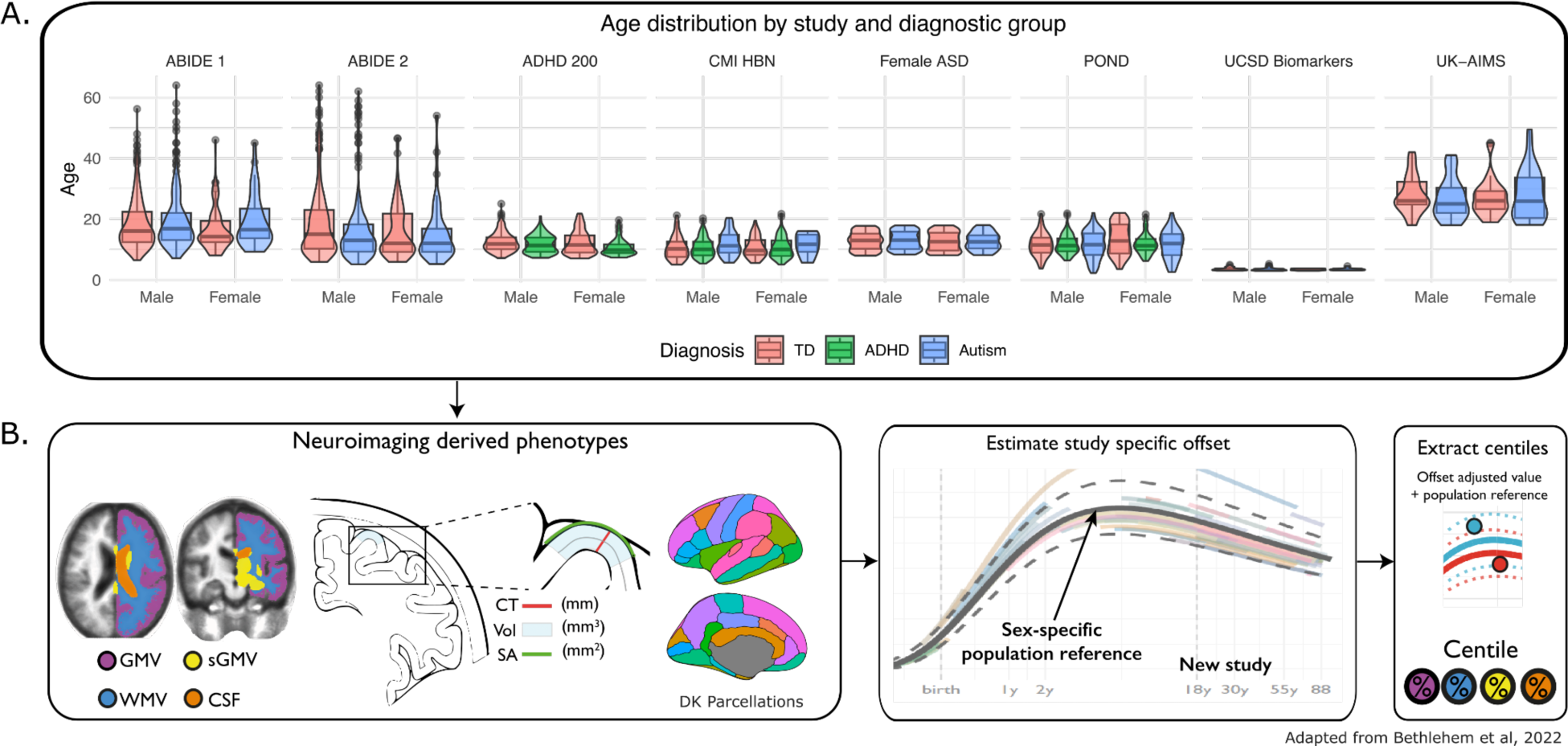
Study demographics and methods overview. **A.** Box and violin plots representing age distribution of each study by diagnostic group and sex. **B.** Methods overview. Global cortical and subcortical grey matter volume (GMV, sGMV), white matter volume (WMV), and ventricular cerebrospinal fluid (CSF) volume, and regional cortical thickness, volume and surface area based on the Desikan-Killiany (DK) parcellations were estimated for each participant. Sex-specific lifespan developmental trajectories for each neuroanatomical measure were estimated using generalised additive models of location scale and shape (GAMLSS) for a sample of 75,241 typically developing individuals, accounting for site and scanner specific variables ^103^. Out-of-sample estimates for the study sample used here were generated based on these reference models, resulting in a (per)centile score for each measure of each participant, indicating where they fall within the sample range (0-1).

Ethical approval and informed consent from all participants was obtained for each dataset in the primary study. The Cambridge Psychology Research Ethics Committee (PRE.2020.104) deemed secondary analysis of de-identified data not to require ethical oversight.

### Data processing

#### Freesurfer and cortical parcellations

T1 images from all datasets were processed individually using the FreeSurfer processing pipeline ^110^ (http://surfer.nmr.mgh.harvard.edu/), version 6.0.1. Regional estimates of each cortical measure were extracted based on the Desikan-Killiany ^111^ atlas, which parcellates the cortex into 34 gyral-based subdivisions per hemisphere. For computational efficiency, and because at the time of analysis BrainChart models were only available for averaged hemispheres, measures were averaged across hemispheres for each parcellation for further analysis.

#### Quality control

All scans underwent manual QC of raw image and FreeSurfer surface reconstructions, using our recently developed FSQC tool ^112^. FSQC generates images of FreeSurfer outputs overlaid on the raw T1 scan, allowing for the evaluation of quality of the surface reconstruction, as well as artefacts in the raw scan, with particular attention paid to motion ^113^. A cut-off point of 2.5 was used for FSQC, based on previous work ^112^. However, as even small variations in quality can bias downstream analyses ^113,114^, we also included the FreeSurfer-derived Euler number ^115^ as a covariate in all analyses.

#### Generation of centile scores using GAMLSS

Our previous work ^103^ generated normative reference models using generalised additive models of location scale and shape (GAMLSS) to map neuroanatomical developmental trajectories across the lifespan. This was based on a sample of 75,241 typically developing individuals, for total grey matter volume (GMV), subcortical grey matter volume (sGMV), white matter volume (WMV), ventricular volume, total surface area, mean cortical thickness, and regional cortical thickness (CT), volume (CV) and surface area (SA), accounting for age, sex and site/scanner. Out-of-sample normative centile scores for our study sample were generated based on these reference models using Brent’s maximum likelihood estimation (see supplementary methods section 1.8 in Bethlehem et al ^103^), quantifying variation in normative brain development in our sample.

#### ComBat and accounting for site variability

GAMLSS has been demonstrated to adequately account for batch effects related to differences between site- and scanner-specific variables ^103^. However, Bethlehem and colleagues ^103^ noted the relatively lower stability of their models to generate accurate out-of-sample centile estimates for samples of N<100. Due to the relatively smaller sample sizes for some sites in our dataset, and higher variability in the clinical samples, we first harmonised our data using ComBat ^116^, consistent with previous work using the same GAMLSS implementation ^117^. ComBat-harmonised data were used as inputs to the out-of-sample maximum likelihood estimation to generate individualised centile scores.

### Statistical analysis

#### Group differences and sex modulation effects

Separate multiple linear regressions were run to examine group differences by diagnosis in centile scores for all global volumes as well as regional cortical volumes, thickness and surface area. The interaction between diagnosis and sex was examined, and given previous evidence of sex-specific neurobiological correlates in autism and ADHD ^31,40,52,66,82,83,118,119^, a priori sex-stratified analyses examined diagnostic differences in males and females separately. We further assessed the similarity of diagnostic effect size maps between males and females by calculating the Spearman correlation between the two maps, and using spin permutation testing to assess significance with appropriate control of the potentially confounding effect of spatial autocorrelation of each map ^120^.

All analyses included age and Euler number as a covariate (see QC section). Multiple comparisons were controlled for using the false discovery rate (FDR ^121^), for each analysis and cortical measure separately.

Multiple sensitivity analyses were conducted, including analyses controlling for global brain measures, using different QC methods, analysis of equal sex-matched subsamples, and Hartigan’s dip tests ^122^ for multi-modality in the distributions of each group (supplementary methods S2-5). We also investigated dimensional associations between cortical measures and autistic and ADHD traits, in subsets of the data with these measures available (supplementary methods S6).

#### Age modulation effects

The interaction between diagnosis and age was examined in separate models, for both global and regional measures, to assess any age-dependent differences related to neurodevelopmental conditions. Due to the narrower age range of the ADHD sample (5-21 years), we conducted sensitivity analyses including only controls within the same age range (supplementary methods S7).

#### Co-occurring autism and ADHD

Finally, we conducted an exploratory analysis to examine whether individuals with co-occurring autism and ADHD had unique neuroanatomical profiles. We compared a subgroup of 203 individuals (post QC) with recorded clinical diagnoses of both autism and ADHD (autism+ADHD) to the control group, as well as examining interactions with sex, and sex-specific diagnostic effects. We also compared the correlation (using spin tests ^120^) and overlap of brain maps between each pair of diagnostic groups (supplementary methods S8.1). We note that data on secondary diagnosis was not available for all datasets, and can be unreliable. While secondary diagnosis at some sites was confirmed by clinician consensus (e.g. HBN ^109^), at other sites, they were community-based. There are likely individuals missed in this analysis; thus, this analysis was exploratory, and we also attempted to replicate this analysis in a subset of autistic individuals who also met the clinical cut-off criteria for ADHD on the Strengths and Weaknesses of Attention-Deficit/Hyperactivity-symptoms and Normal-behaviours ^123^ (SWAN) (N=118; supplementary methods S8.2).

## Results

### Differences in global brain measures

Individuals with ADHD overall had significantly lower total cortical (*d* = -0.14) and subcortical (*d* = -0.13) GMV, total WMV (*d* = -0.16), and total cortical surface area centile scores (*d* = -0.18), but greater mean CT centiles (*d* = 0.09) relative to controls (all q [FDR corrected p-values] < 0.001). Autistic individuals, but not those with ADHD, had significantly greater ventricular volume centiles (*d* = 0.15, q < 0.0001) relative to controls (Figure 2). There were no significant diagnosis-by-sex interactions for any of the global brain measures for ADHD. For autism, we observed trend-level significant interactions for total sGMV (*P* = 0.009) and ventricular volume (*P* = 0.02; neither surviving FDR correction), where autistic males had significantly greater sGMV (*P* = 0.037; *d* = 0.08) and ventricular volume (*P* < 0.0001, *d* = 0.18) than male controls, but autistic females showed no difference to female controls. There was a trend towards a significant interaction between autism diagnosis and age for total WMV, sGMV and SA, and for sGMV and age for ADHD diagnosis (all *P* <0.05), but none survived FDR.

**Figure 2.**
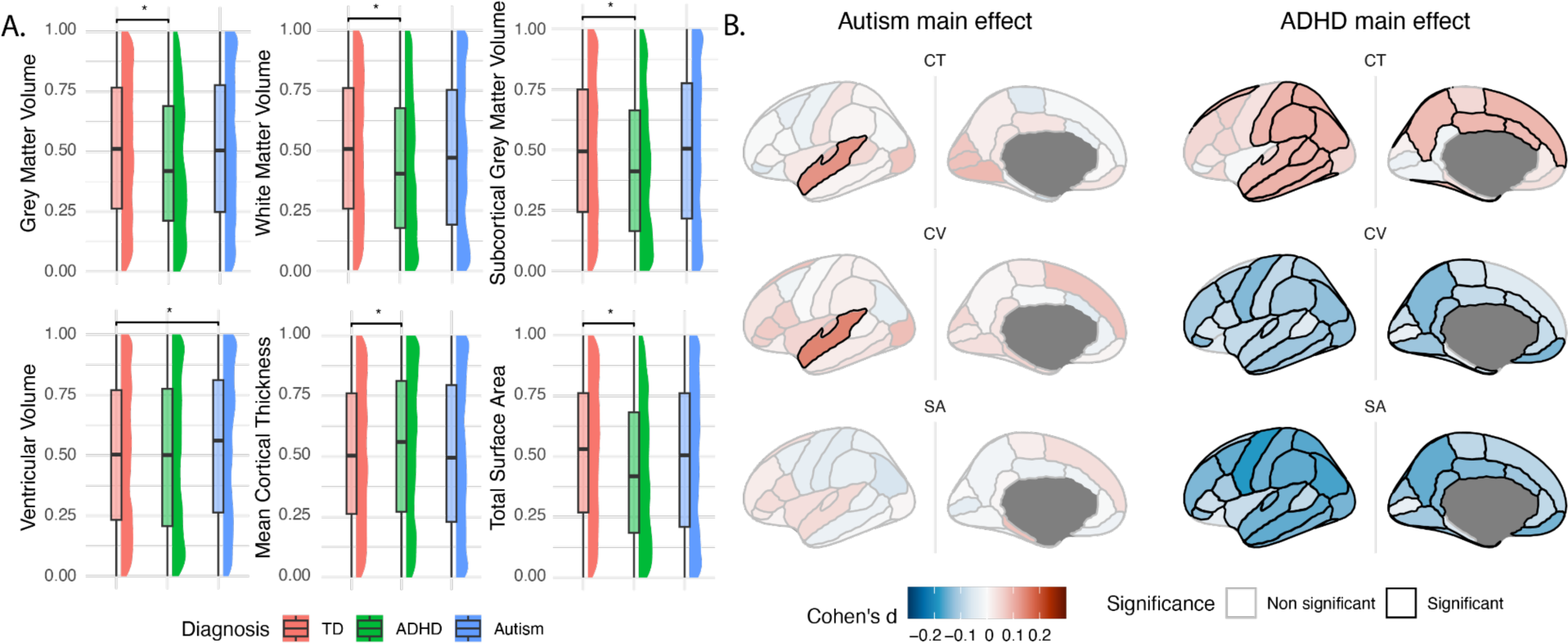
Case-control differences in global and regional centile scores of structural MRI metrics. **A**. Box and raincloud plots showing group differences in global neuroanatomical measures. Raincloud plots show the density distribution of centiles per group. Autistic individuals had significantly larger ventricles than TD individuals, but no differences were observed in any other measures. Individuals with ADHD had significantly lower cortical grey, white, and subcortical grey matter volume and total surface area centiles relative to controls, but greater mean cortical thickness centiles. **B**. Regional group differences. Brain maps show Cohen’s d effect sizes, with significant regions (passing 5% FDR) outlined in black. Red represents positive effect sizes (autism or ADHD > controls), and blue represents negative effect sizes (autism or ADHD < controls). Overall, autistic individuals had significantly greater cortical volume and thickness in the superior temporal gyrus; whereas individuals with ADHD had significant and widespread decreases in cortical volume and surface area, and increases in cortical thickness.

### Regional differences

#### Main effects

Significant group differences in regional centiles were much more widespread in ADHD than autism (Figure 2). All effect sizes were relatively small. In autistic individuals, CT and CV, but not SA, centiles, were increased in the superior temporal gyrus (STG; *d* = 0.13-0.15) only. Individuals with ADHD presented with significantly lower CV and SA centiles across most cortical regions (*d* = -0.07--0.18), but higher CT centiles (*d* = 0.09-0.10). Importantly, controlling for global measures drastically altered effects for ADHD but not for autism, highlighting that the results in ADHD are less specific and driven largely by global effects, but much more localised for autism. In particular, increases in CT were substantially attenuated and no longer significant, and decreases in CV and SA in ADHD were no longer apparent, with increases instead observed in some regions (supplementary results S1). Different methods of accounting for QC did not have a major impact on results (supplementary results S2).

#### Interaction with sex and sex-stratified results

For autism, there was a significant interaction with sex for CV in the STG, insula, and temporal pole (Figure 3A). It is important to note that the significant diagnostic main effect on STG CV must be interpreted in light of this interaction, and appears to apply to autistic males only. There were no significant diagnosis-by-sex interactions for any regions for ADHD.

**Figure 3.**
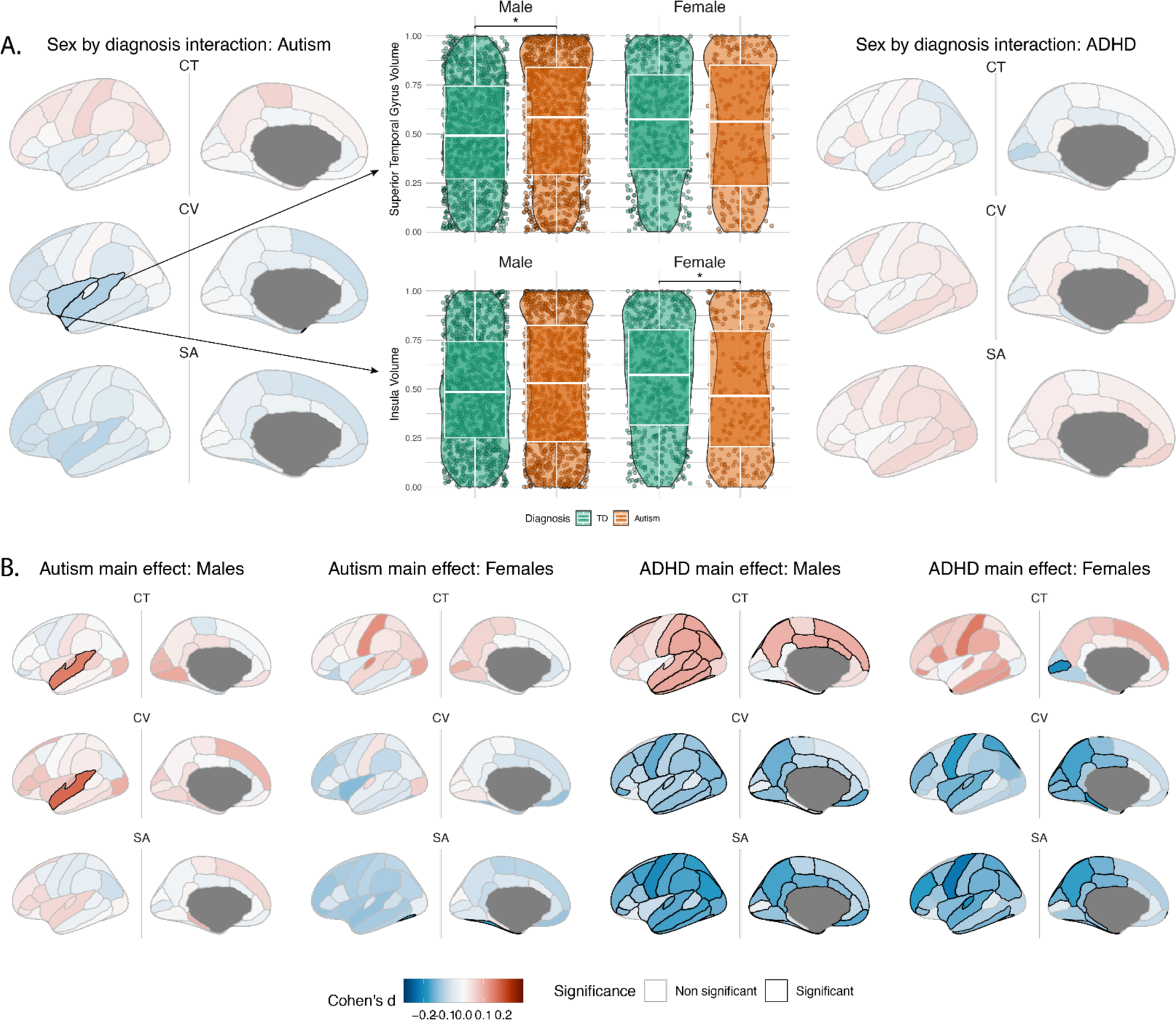
Interactions between sex and diagnostic group on centile scores of regional MRI metrics. **A.** Brain maps showing effect sizes and significance of interaction per brain region, and box and violin plots showing comparison of values broken down by group for two significant regions. **B.** Sex-stratified regional association with diagnosis. All maps show Cohen’s d effect sizes, with significant regions (passing 5% FDR) outlined in black. Red represents positive effect sizes (autism or ADHD > controls), and blue represents negative effect sizes (autism or ADHD < controls).

We also conducted a priori sex-stratified analyses of relationships between regional neuroanatomy and diagnosis (Figure 3B). Compared with same-sex controls, autistic males, but not autistic females, had significantly greater CV and CT centiles in the STG (*d =* 0.15-0.18), whereas autistic females had significantly lower cortical SA centiles only in the fusiform gyrus relatively to controls (*d* = -0.18). Subthreshold effect size maps showed similar spatial patterning between males and females for CT (rho = 0.5, p*_spin_* = 0.024), but were quite different for CV (rho = 0.24, p*_spin_* = 0.13) and SA (rho = -0.06, p*_spin_* = 0.40) (Figure 3B; Supplementary results S3). Males with ADHD had significantly lower CV and SA (*d =* -0.08 - -0.20) and higher CT centiles (*d* = 0.10-0.11) across much of the cortex relative to male controls. Unsurprisingly, given the lack of significant interaction, females with ADHD had very similar patterns of cortical alterations, though with fewer regions reaching significance (CV and SA: *d* = -0.13--0.22; CT: *d* = 0.18). Male and female subthreshold effect size maps were visually similar and showed high spatial overlap for all cortical measures (rho = 0.34-0.59; p*_spin_* = 0.0005-0.029).

Effect sizes and directions remained largely consistent in the sex-matched subsample analyses, though with some differences in the number of significant regions (supplementary results S4). Multimodal distributions of centiles were observed across most of the cortex for the autistic group, compared to only a few regions for ADHD and controls (supplementary results S5). Dimensional analyses of autistic and ADHD traits revealed limited significant but weak associations between some clinical and cortical measures (supplementary results S6).

#### Interactions with age

We also explored whether diagnostic group differences are stable across age, by examining diagnosis-by-age interactions. A significant interaction between age and autism diagnosis was observed only in the superior frontal gyrus for CT centiles, and between age and ADHD diagnosis for CT and CV centiles in superior frontal, temporal, and occipital regions. For autism, there was a slight positive but significant correlation between age and CT centile (partial r = 0.11), with no significant correlation in controls. For ADHD, in the superior frontal, caudal middle frontal cortex, transverse temporal and pericalcarine cortices, for both thickness and volume, there was a significant positive correlation with age in the ADHD group (partial r = 0.07-0.14), but minimal or no significant correlation in the control group. Interestingly, for CV, the regions in which a significant interaction with age was observed were the same regions that displayed a significant effect of ADHD diagnosis when controlling for total GMV (Figure S1.1). In the insula for CT, however, there was a significant negative correlation in the ADHD group (r = -0.14--0.15), but no significant correlation in the control group (Figure 4). The sensitivity analysis for ADHD in the age-matched control sample yielded largely similar results for CT, with more regions reaching significance, but effects for CV were no longer significant (supplementary results section S7).

**Figure 4.**
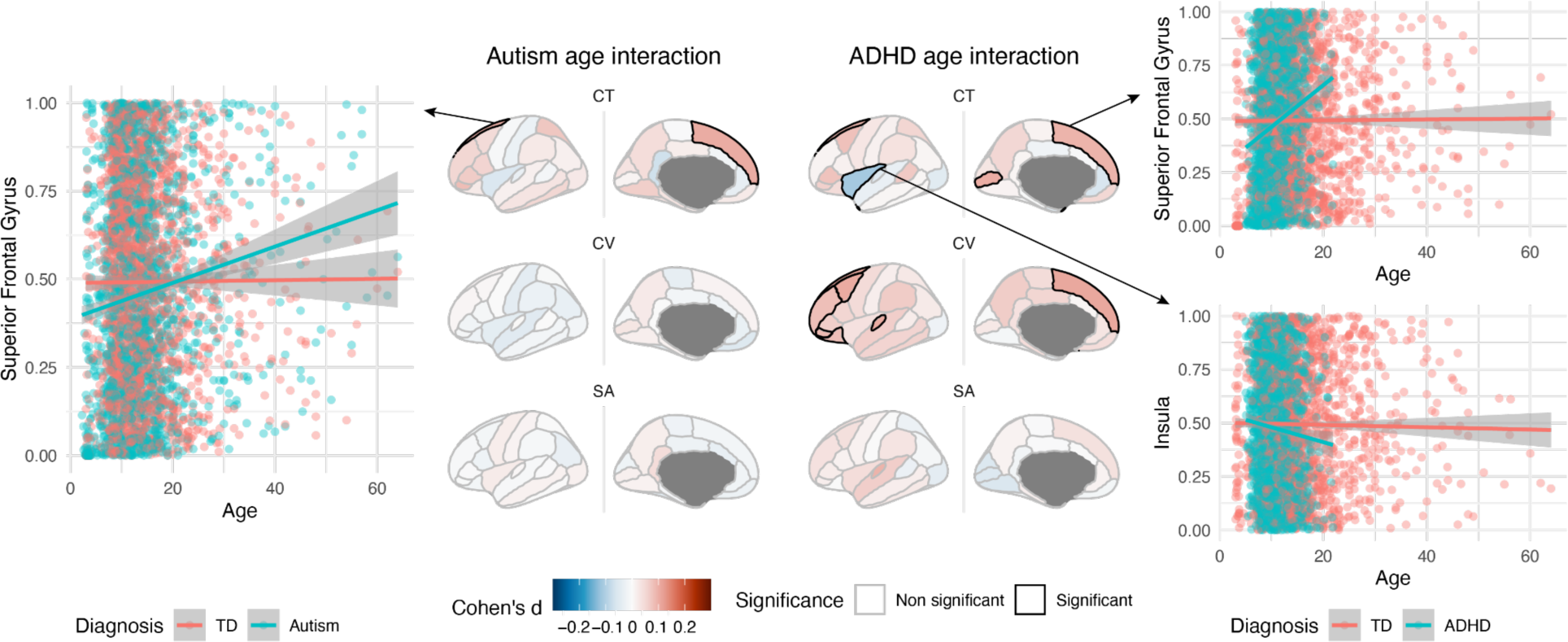
Regional interactions between diagnosis and age. Brain maps show interaction effect sizes and regional significance, and scatter plots show the relationship between CT and age in the autism/ADHD and TD groups in regions where a significant interaction was observed.

#### Co-occurring autism and ADHD

The autism+ADHD group showed a different pattern of alterations, with some overlap, to those with only one diagnosis (Figure 5), with widespread significant increases in CT centiles relative to controls (*d* = 0.10-0.24), decreased SA centiles in frontal and parietal regions (*d =* -0.11--0.14), and no differences in CV centiles. There was no significant interaction with sex. Male effects resembled those observed in the whole group, but there were no significant differences observed in the female group (Figure 5A). Spin tests and an overlap analysis revealed the greatest similarity in effect size maps between the autism+ADHD and ADHD groups, with minimal overlap between the autism and ADHD only groups. CT and SA both showed widespread homology in effect size direction across all three groups, though with minimal overlap of significance, whereas CV primarily showed overlap between autism+ADHD and ADHD. The only region with an overlap of significance between autism and ADHD was the STG, but in opposite directions (Figure 5B).

**Figure 5.**
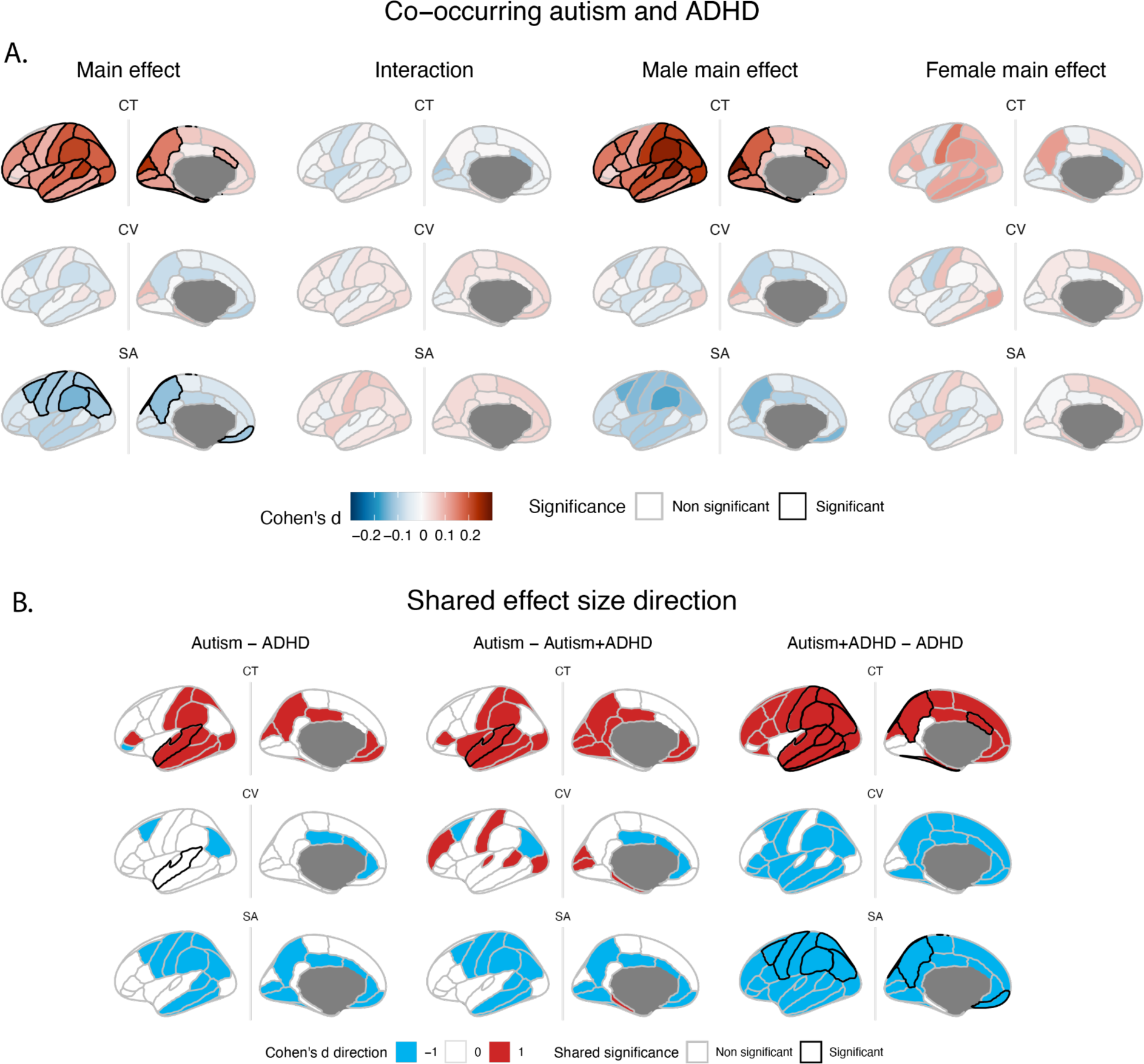
Cortical alterations (relative to controls) in individuals with co-occurring autism and ADHD and overlap of effect size significance and direction. **A.** Main effects of diagnosis relative to controls; interaction with sex; main effects in males, and main effects in females. All maps show Cohen’s d effect sizes, with significant regions (passing 5% FDR) outlined in black. Red represents positive effect sizes (autism+ADHD > controls), and blue represents negative effect sizes (autism+ADHD < controls). **B.** Brain maps showing the pairwise overlap of effect size direction and significance for the autism, ADHD and autism+ADHD groups. Regions which had a positive effect size in both groups’ analysis (in comparison to controls) are shown in red; regions which had a negative effect size in both groups are shown in blue. Regions in white were in discordant directions between groups. Regions which were significantly different from controls in both groups are outlined in black. Note that the superior temporal gyrus showed a significant effect in both autism and ADHD for cortical volume, but in opposite directions.

Most results were no longer significant after controlling for global measures (Figure S8.2). The replication analysis based on the SWAN cut-off yielded similar results, though with slightly fewer regions reaching significance. The biggest difference in results was in the female main effect which more closely resembled the male main effect in the SWAN cut-off subsample (Supplementary Figure S8.3).

## Discussion

Using a large aggregated dataset and existing models of typical brain development, we observed robust neuroanatomical signatures of both autism and ADHD, with evidence for significant sex-modulations in autism, and age-specific effects primarily in ADHD, as well as distinct alterations in individuals with co-occurring autism and ADHD. We also observed some limited significant associations between cortical measures and measures of autistic and ADHD traits, but notably there was no significant association with the ADOS, considered the gold standard measure of autistic traits.

Previous normative modelling studies on a single diagnostic cohort have mainly observed greater divergence from typical brain development in autism or ADHD in highly individualised patterns ^45,54,124^, or multiple autistic subgroups with distinct patterns of deviation and clinical profiles ^48,50^, rather than overall diagnostic group differences. We note that our sample size is considerably larger than previous studies, so we are likely better powered to detect average group differences that are consistent across datasets. However, the results of the studies mentioned above reflect that this is not the whole picture, and highlight the significant heterogeneity of the conditions, and the complexities of characterising the underlying neuroanatomy. It will be interesting to see in future work if a normative modelling approach is more adept at detecting data-driven subtypes.

We did not observe the greater total grey matter volume or surface area in autistic individuals that have been reported previously ^20–22,35,125,126^, although it should be noted that we did not explicitly test the early age range that was the focus of most of these studies. We did however replicate findings of enlarged ventricular volume related to autism ^125–131^, and our findings of significantly greater localised, regional cortical thickness and volume are at least partly consistent with recent, large-sample studies ^30,31,130,132^. Increases in the superior temporal gyrus (STG) have been commonly reported in autism ^30,31,35,54,132–141^. STG and surrounding cortical regions are also repeatedly implicated in autism and known to be involved in cognitive functions that are often impacted (e.g., language) in autistic individuals ^25,142–146^. The present results in ADHD were much more widespread and less localised than in autism. We confirm previous reports of global reductions in grey and white matter volume and total surface area in ADHD, as well as widespread regional decreases in CV and SA ^59–63,65^. We also confirm recent reports of greater, but not lower, CT, which contradict some earlier studies of ADHD ^60–64,72,73^. Of note, head motion poses a critical challenge in neuroimaging studies of ADHD ^72^ (as well as autism ^113,147^). Thus, it is possible that earlier reports of cortical thinning in ADHD were confounded by motion and other aspects of MRI quality ^148^, along with small sample sizes, as has been observed in autism ^112,113^. It will be important in future work to extend these investigations to specific subcortical structures and the cerebellum, which have also been identified as important structures in both autism and ADHD ^74,149–151^.

It is interesting to note the divergent direction of diagnostic effects in autism and ADHD, and the difference in cortical measures between conditions. CT, CV and SA are related to distinct neurodevelopmental processes and genetic underpinnings ^25,152–160^, with volume and surface area more closely related than to thickness ^161^. SA and CT also show different anterior-posterior versus dorsal-ventral patterning gradients that may be linked to underlying molecular processes in prenatal gene expression that create such patterning and cortical regionalization ^25,157–159,162,163^. Such differential cortical patterning of SA and CT linked to differential genomic mechanisms might be particularly important in autism ^25,164^. Thus, these different cortical measures could point to different underlying neurobiological mechanisms or processes related to the emergence of each condition.

The overall main effect of autism appeared to be driven by males, who comprise the majority of the sample, with distinct alterations observed for females, in particular for CV and SA. Critically, this suggests that inferences drawn from the whole sample might not be applicable to autistic females specifically. In contrast, in ADHD we did not observe evidence for such sex modulation, given the lack of significant interaction and the substantial similarity of male and female effect size maps. A still outstanding and important future question is to what extent the observed sex effects in the cortical measures, alongside other presentations of neurodevelopmental conditions, are due to underlying differences in biology versus socialisation, gender identity effects, gender expectations or diagnostic bias ^165^.

Significant associations between the degree of neuroanatomical divergence and autistic traits have also often been reported by previous studies ^45,54,166^, in contrast to the lack of significant association observed here with the ADOS CSS, despite the large sample size. A significant caveat here is that due to the multi-site nature of the data, these analyses were conducted on a subset of participants only, which might partially explain the lack of robust autistic trait associations with cortical measures. Some significant but weak cortical associations were observed for the RBS-R, SRS and SWAN. Interestingly, the effect size maps observed for RBS-R and SRS associations (both autistic trait measures) more closely resembled cortical alterations observed for ADHD (widespread decreases in CV and SA), whereas the effect size maps for the SWAN (an ADHD traits measure) was similar to those for autism (significant increases in STG CT, and CV when controlling for GMV). This perhaps highlights the lack of clear boundaries between diagnostic categories, as well as the non-specificity of some clinical dimensional measures, which might not be picking up on the same things in different diagnostic groups.

The absence of an age-by-diagnosis interaction across global measures and most cortical regions in autism offers limited support for the hypothesis of early brain overgrowth and normalisation with age ^125,167^. However, longitudinal data are needed to properly investigate these relationships and hypotheses. The regional age-interaction for ADHD suggests that the nature of these deviations in ADHD is not static across development, at least in some cortical measures.

Finally, in the exploratory analyses for the autism+ADHD group, we observed CT alterations in the same direction but more widespread than those in either autism or ADHD alone, and SA alterations that resemble those seen in ADHD, but more localised. It might be that these differences in the autism+ADHD group represent a synthesised phenotype, but we caution against a simplified interpretation. Previous studies have not identified significant differences in cortical thickness between an autism+ADHD group and controls; however, the methods used were quite different, and sample sizes relatively small ^168,169^. A significant caveat is that secondary diagnoses were not available for all datasets, and even when available, there are likely missed secondary diagnoses, based on known rates of co-occurrence ^19,170^. For this exploratory analysis we focused on individuals from whom we had clearly recorded secondary diagnoses. Future research could be improved and this picture clarified if co-occurring and secondary diagnoses were reported consistently across studies, along with consistent dimensional clinical data. However, we believe these preliminary findings provide an interesting avenue for future research.

The present results should be interpreted in light of some limitations. First, as is increasingly common, the data come from multiple sources, with different scanners, scanning protocols, recruitment procedures, and demographic characteristics. We have attempted to address this variability as rigorougly as possible: all data was analysed in house, using the same methods and procedures, and data was harmonised in a two-step process, using ComBat and then GAMLSS. While it is impossible to fully eliminate site effects, we believe that the size of this dataset, and in particular, the large female sample and availability of both autism and ADHD data, mitigate these issues. However, we note that the effect sizes observed in most analyses were very small and thus may have limited clinical or practical significance. Second, the lack of consistent phenotypic and diagnostic information across studies and sites leads to limited data in the analyses of relationships with clinical measures, and of co-occurring diagnoses, as well as to investigate the potential confounding impact of medication history in ADHD. Third, despite its large size, the representativeness of the sample is still suboptimal. There is still a large imbalance in the number of diagnosed males and females, as well as a substantial lack of participants with lower IQ and/or high support needs, and insufficient diversity across racial-ethnic groups. Finally, the lack of longitudinal data limits our ability to draw conclusions about developmental trajectories over time, and should be a priority of future studies and data collection initiatives.

## Conclusions

This study identifies distinct profiles of neuroanatomical divergence associated with autism and ADHD, that are differentially modulated by age and sex. These observations offer valuable insights into associated developmental processes and could potentially serve as indicators of biomarkers. We also identified potential differential impacts of co-occurring diagnoses of autism and ADHD, but note that data on secondary diagnosis is not always reliable. Future work should further investigate individual variability and the existence of subgroups within and across diagnoses.

## Data Availability

All data produced in the present study are available upon reasonable request to the authors.

## Acknowledgements

SB is supported by a Trinity College Coutts-Trotter studentship. SBC received funding from the Wellcome Trust 214322\Z\18\Z. For the purpose of Open Access, the author has applied a CC BY public copyright licence to any Author Accepted Manuscript version arising from this submission. The results leading to this publication have received funding from the Innovative Medicines Initiative 2 Joint Undertaking under grant agreement No 777394 for the project AIMS-2-TRIALS. This Joint Undertaking receives support from the European Union’s Horizon 2020 research and innovation programme and EFPIA and AUTISM SPEAKS, Autistica, SFARI. The funders had no role in the design of the study; in the collection, analyses, or interpretation of data; in the writing of the manuscript, or in the decision to publish the results. SBC also received funding from the Autism Centre of Excellence, SFARI, the Templeton World Charitable Fund and the MRC. All research at the Department of Psychiatry in the University of Cambridge was supported by the NIHR Cambridge Biomedical Research Centre (NIHR203312) and the NIHR Applied Research Collaboration East of England. Any views expressed are those of the author(s) and not necessarily those of the funders, IHU-JU2, the NIHR or the Department of Health and Social Care. EB was supported by an NIHR Cambridge Biomedical Research Centre and NIHR Senior Investigator award. PAD receives grant support from the Alberta Innovates Translational Health Chair in Child and Youth Mental Health, CIHR, Canadian Foundation for Innovation, and the Alberta Children’s Hospital Foundation and research support from Biohaven Pharmaceuticals. M-C Lai is supported by the Academic Scholars Award from the Department of Psychiatry, University of Toronto, the Discovery Fund from the Centre for Addiction and Mental Health, and the Canadian Institutes of Health Research Sex and Gender Science Chair (GSB 171373). Funding was provided by the Ontario Brain Institute to EA and JPL. EC and KP were supported by NIDCD grant R01DC016385, and KP was also supported by NIMH grant R01MH104446.

The Medical Research Council Autism Imaging Multicentre Study Consortium (MRC AIMS Consortium) is a UK collaboration between the Institute of Psychiatry, Psychology and Neuroscience (IoPPN) at King’s College, London, the Autism Research Centre, University of Cambridge, and the Autism Research Group, University of Oxford. Members of MRC AIMS Consortium include: Anthony J. Bailey (Oxford), Simon Baron-Cohen (Cambridge), Patrick F. Bolton (IoPPN), Edward T. Bullmore (Cambridge), Sarah Carrington (Oxford), Marco Catani (IoPPN), Bhismadev Chakrabarti (Cambridge), Michael C. Craig (IoPPN), Eileen M. Daly (IoPPN), Sean C. L. Deoni (IoPPN), Christine Ecker (IoPPN), Francesca Happé (IoPPN), Julian Henty (Cambridge), Peter Jezzard (Oxford), Patrick Johnston (IoPPN), Derek K. Jones (IoPPN), Meng-Chuan Lai (Cambridge), Michael V. Lombardo (Cambridge), Anya Madden (IoPPN), Diane Mullins (IoPPN), Clodagh M. Murphy (IoPPN), Declan G. M. Murphy (IoPPN), Greg Pasco (Cambridge), Amber N. V. Ruigrok (Cambridge), Susan A. Sadek (Cambridge), Debbie Spain (IoPPN), Rose Stewart (Oxford), John Suckling (Cambridge), Sally J. Wheelwright (Cambridge) and Steven C. Williams (IoPPN).

For members of the Lifespan Brain Chart Consortium please refer to: https://docs.google.com/spreadsheets/d/1D8YNDcnhwlv2WcUDhreq3fwrkfpfGiFp0OGFVO5d-es/edit?usp=sharing

## References

1. Shaw, P., Gogtay, N. & Rapoport, J. Childhood psychiatric disorders as anomalies in neurodevelopmental trajectories. Hum. Brain Mapp. 31, 917–925 (2010).

2. Luo, Y., Weibman, D., Halperin, J. M. & Li, X. A Review of Heterogeneity in Attention Deficit/Hyperactivity Disorder (ADHD). Front. Hum. Neurosci. 13, 42 (2019).

3. Masi, A., DeMayo, M. M., Glozier, N. & Guastella, A. J. An Overview of Autism Spectrum Disorder, Heterogeneity and Treatment Options. Neuroscience Bulletin vol. 33 183–193 Preprint at 10.1007/s12264-017-0100-y (2017).

4. Lai, M.-C., Lombardo, M. V. & Baron-Cohen, S. Autism. Lancet 383, 896–910 (2014).

5. Kofler, M. J. et al. Heterogeneity in ADHD: Neurocognitive predictors of peer, family, and academic functioning. Child Neuropsychol. 23, 733–759 (2017).

6. Lenroot, R. K. & Yeung, P. K. Heterogeneity within Autism Spectrum Disorders: What have We Learned from Neuroimaging Studies? Front. Hum. Neurosci. 7, 733 (2013).

7. Karalunas, S. L. & Nigg, J. T. Heterogeneity and Subtyping in Attention-Deficit/ Hyperactivity Disorder-Considerations for Emerging Research Using Person-Centered Computational Approaches. Biol. Psychiatry 88, 103–110 (2020).

8. Ecker, C. The neuroanatomy of autism spectrum disorder: An overview of structural neuroimaging findings and their translatability to the clinical setting. Autism 21, 18–28 (2017).

9. Willcutt, E. G. The prevalence of DSM-IV attention-deficit/hyperactivity disorder: a meta-analytic review. Neurotherapeutics 9, 490–499 (2012).

10. Christensen, D. L. et al. Prevalence and characteristics of autism spectrum disorder among children aged 8 years - autism and Developmental Disabilities Monitoring Network, 11 sites, United States, 2012. MMWR Surveill. Summ. 65, 1–23 (2018).

11. Loomes, R., Hull, L. & Mandy, W. P. L. What Is the Male-to-Female Ratio in Autism Spectrum Disorder? A Systematic Review and Meta-Analysis. J. Am. Acad. Child Adolesc. Psychiatry 56, 466–474 (2017).

12. Lai, M.-C., Lombardo, M. V., Auyeung, B., Chakrabarti, B. & Baron-Cohen, S. Sex/gender differences and autism: setting the scene for future research. J. Am. Acad. Child Adolesc. Psychiatry 54, 11–24 (2015).

13. Mandy, W. et al. Sex differences in autism spectrum disorder: Evidence from a large sample of children and adolescents. J. Autism Dev. Disord. 42, 1304–1313 (2012).

14. Rucklidge, J. J. Gender differences in attention-deficit/hyperactivity disorder. Psychiatr. Clin. North Am. 33, 357–373 (2010).

15. Mo, K. et al. Sex/gender differences in the human autistic brains: A systematic review of 20 years of neuroimaging research. NeuroImage: Clinical vol. 32 Preprint at 10.1016/j.nicl.2021.102811 (2021).

16. Ghirardi, L. et al. The familial co-aggregation of ASD and ADHD: A register-based cohort study. Mol. Psychiatry 23, 257–262 (2018).

17. Miller, M. et al. Sibling Recurrence Risk and Cross-aggregation of Attention-Deficit/Hyperactivity Disorder and Autism Spectrum Disorder. JAMA Pediatr. 173, 147–152 (2019).

18. Leitner, Y. The co-occurrence of autism and attention deficit hyperactivity disorder in children - what do we know? Front. Hum. Neurosci. 8, 268 (2014).

19. Hours, C., Recasens, C. & Baleyte, J.-M. ASD and ADHD Comorbidity: What Are We Talking About? Front. Psychiatry 13, 837424 (2022).

20. Courchesne, E., Carper, R. & Akshoomoff, N. Evidence of brain overgrowth in the first year of life in autism. JAMA 290, 337–344 (2003).

21. Hazlett, H. C. et al. Early brain overgrowth in autism associated with an increase in cortical surface area before age 2 years. Arch. Gen. Psychiatry 68, 467–476 (2011).

22. Sacco, R., Gabriele, S. & Persico, A. M. Head circumference and brain size in autism spectrum disorder: A systematic review and meta-analysis. Psychiatry Research - Neuroimaging 234, 239–251 (2015).

23. Ohta, H. et al. Increased Surface Area, but not Cortical Thickness, in a Subset of Young Boys With Autism Spectrum Disorder. Autism Res. 9, 232–248 (2016).

24. Amaral, D. G., et al. In pursuit of neurophenotypes: The consequences of having autism and a big brain. Autism Research vol. 10 711–722 Preprint at 10.1002/aur.1755 (2017).

25. Lombardo, M. V. et al. Atypical genomic cortical patterning in autism with poor early language outcome. Sci Adv 7, eabh1663 (2021).

26. Nordahl, C. W. et al. Brain enlargement is associated with regression in preschool-age boys with autism spectrum disorders. Proc. Natl. Acad. Sci. U. S. A. 108, 20195–20200 (2011).

27. Lee, J. K. et al. Longitudinal Evaluation of Cerebral Growth Across Childhood in Boys and Girls With Autism Spectrum Disorder. Biol. Psychiatry (2020).

28. Hardan, A. Y., Muddasani, S., Vemulapalli, M., Keshavan, M. S. & Minshew, N. J. An MRI study of increased cortical thickness in autism. Am. J. Psychiatry 163, 1290–1292 (2006).

29. Hyde, K. L., Samson, F., Evans, A. C. & Mottron, L. Neuroanatomical differences in brain areas implicated in perceptual and other core features of autism revealed by cortical thickness analysis and voxel-based morphometry. Hum. Brain Mapp. 31, 556–566 (2010).

30. Khundrakpam, B. S., Lewis, J. D., Kostopoulos, P., Carbonell, F. & Evans, A. C. Cortical Thickness Abnormalities in Autism Spectrum Disorders Through Late Childhood, Adolescence, and Adulthood: A Large-Scale MRI Study. Cereb. Cortex 27, 1721–1731 (2017).

31. Bedford, S. A. et al. Large-scale analyses of the relationship between sex, age and intelligence quotient heterogeneity and cortical morphometry in autism spectrum disorder. Mol. Psychiatry (2019) doi:10.1038/s41380-019-0420-6.

32. Ecker, C. et al. The effect of age, diagnosis, and their interaction on vertex-based measures of cortical thickness and surface area in autism spectrum disorder. J. Neural Transm. 121, 1157–1170 (2014).

33. Hadjikhani, N., Joseph, R. M., Snyder, J. & Tager-Flusberg, H. Anatomical differences in the mirror neuron system and social cognition network in autism. Cereb. Cortex 16, 1276– 1282 (2006).

34. Mak-Fan, K. M., Taylor, M. J., Roberts, W. & Lerch, J. P. Measures of cortical grey matter structure and development in children with autism spectrum disorder. J. Autism Dev. Disord. 42, 419–427 (2012).

35. Hazlett, H. C. et al. Early brain development in infants at high risk for autism spectrum disorder. Nature 542, 348–351 (2017).

36. Yang, D. Y.-J., Beam, D., Pelphrey, K. A., Abdullahi, S. & Jou, R. J. Cortical morphological markers in children with autism: a structural magnetic resonance imaging study of thickness, area, volume, and gyrification. Mol. Autism 7, (2016).

37. Valk, S. L., Di Martino, A., Milham, M. P. & Bernhardt, B. C. Multicenter mapping of structural network alterations in autism. Hum. Brain Mapp. 36, 2364–2373 (2015).

38. Scheel, C. et al. Imaging derived cortical thickness reduction in high-functioning autism: Key regions and temporal slope. Neuroimage 58, 391–400 (2011).

39. Herrington, J. D. et al. Amygdala Volume Differences in Autism Spectrum Disorder Are Related to Anxiety. J. Autism Dev. Disord. 47, 3682–3691 (2017).

40. Nordahl, C. W. et al. High Psychopathology Subgroup in Young Children With Autism: Associations With Biological Sex and Amygdala Volume. J. Am. Acad. Child Adolesc. Psychiatry (2020) doi:10.1016/j.jaac.2019.11.022.

41. Schumann, C. M., Barnes, C. C., Lord, C. & Courchesne, E. Amygdala Enlargement in Toddlers with Autism Related to Severity of Social and Communication Impairments. Biol. Psychiatry 66, 942–949 (2009).

42. Li, D., Karnath, H.-O. & Xu, X. Candidate Biomarkers in Children with Autism Spectrum Disorder: A Review of MRI Studies. Neurosci. Bull. 33, 219–237 (2017).

43. Zielinski, B. A. et al. autism and typical development. (2014) doi:10.1093/brain/awu083.

44. Ecker, C. et al. Association Between the Probability of Autism Spectrum Disorder and Normative Sex-Related Phenotypic Diversity in Brain Structure. JAMA Psychiatry 74, 329 (2017).

45. Bethlehem, R. A. I. et al. A normative modelling approach reveals age-atypical cortical thickness in a subgroup of males with autism spectrum disorder. Commun Biol 3, 486 (2020).

46. Lin, H.-Y., Ni, H.-C., Lai, M.-C., Tseng, W.-Y. I. & Gau, S. S.-F. Regional brain volume differences between males with and without autism spectrum disorder are highly age-dependent. Mol. Autism 6, 29 (2015).

47. Mizuno, Y. et al. Structural brain abnormalities in children and adolescents with comorbid autism spectrum disorder and attention-deficit/hyperactivity disorder. Transl. Psychiatry 9, (2019).

48. Zabihi, M. et al. Fractionating autism based on neuroanatomical normative modeling. Transl. Psychiatry 10, (2020).

49. Hong, S.-J., Valk, S. L., Di Martino, A., Milham, M. P. & Bernhardt, B. C. Multidimensional Neuroanatomical Subtyping of Autism Spectrum Disorder. Cereb. Cortex 28, 3578–3588 (2018).

50. Shan, X. et al. Mapping the Heterogeneous Brain Structural Phenotype of Autism Spectrum Disorder Using the Normative Model. Biol. Psychiatry 91, 967–976 (2022).

51. Schaer, M., Kochalka, J., Padmanabhan, A., Supekar, K. & Menon, V. Sex differences in cortical volume and gyrification in autism. Mol. Autism 6, 42 (2015).

52. Hammill, C. et al. Quantitative and Qualitative Sex Modulations in the Brain Anatomy of Autism. Biol Psychiatry Cogn Neurosci Neuroimaging 0, (2021).

53. Van’t Westeinde, A. et al. Sex differences in brain structure: a twin study on restricted and repetitive behaviors in twin pairs with and without autism. Mol. Autism 11, 1 (2020).

54. Zabihi, M. et al. Dissecting the Heterogeneous Cortical Anatomy of Autism Spectrum Disorder Using Normative Models. Biol Psychiatry Cogn Neurosci Neuroimaging 4, 567– 578 (2019).

55. Bloss, C. S. & Courchesne, E. MRI neuroanatomy in young girls with autism. J. Am. Acad. Child Adolesc. Psychiatry 46, 515–523 (2007).

56. Schumann, C. M. et al. Longitudinal magnetic resonance imaging study of cortical development through early childhood in autism. Journal of Neuroscience 30, 4419–4427 (2010).

57. Chen, C. & Van Horn, J. D. Developmental neurogenetics and multimodal neuroimaging of sex differences in autism. Brain Imaging Behav. 11, 38–61 (2017).

58. Samea, F. et al. Brain alterations in children/adolescents with ADHD revisited: A neuroimaging meta-analysis of 96 structural and functional studies. Neurosci. Biobehav. Rev. 100, 1–8 (2019).

59. Albajara Sáenz, A., Villemonteix, T. & Massat, I. Structural and functional neuroimaging in attention-deficit/hyperactivity disorder. Developmental Medicine and Child Neurology vol. 61 399–405 Preprint at 10.1111/dmcn.14050 (2019).

60. Ambrosino, S., de Zeeuw, P., Wierenga, L. M., van Dijk, S. & Durston, S. What can Cortical Development in Attention-Deficit/Hyperactivity Disorder Teach us About the Early Developmental Mechanisms Involved? Cereb. Cortex 27, 4624–4634 (2017).

61. Wolosin, S. M., Richardson, M. E., Hennessey, J. G., Denckla, M. B. & Mostofsky, S. H. Abnormal cerebral cortex structure in children with ADHD. Hum. Brain Mapp. 30, 175–184 (2009).

62. Sarabin, E., Harkness, K. & Murias, K. The Relationship Between Cortical Thickness and Executive Function Measures in Children With and Without ADHD. J. Atten. Disord. 10870547231174036 (2023).

63. Lin, H. et al. Population level multimodal neuroimaging correlates of attention-deficit hyperactivity disorder among children. Front. Neurosci. 17, 1138670 (2023).

64. Maier, S. et al. Left insular cortical thinning differentiates the inattentive and combined subtype of adult attention-deficit/hyperactivity disorder. J. Psychiatr. Res. 159, 196–204 (2023).

65. Kasparek, T., Theiner, P. & Filova, A. Neurobiology of ADHD from childhood to adulthood: Findings of imaging methods. *J*. Atten. Disord. 19, 931–943 (2015).

66. Almeida Montes, L. G., et al. Brain Cortical Thickness in ADHD: Age, Sex, and Clinical Correlations. J. Atten. Disord. 17, 641–654 (2013).

67. Silk, T. J. et al. Cortical morphometry in attention deficit/hyperactivity disorder: Contribution of thickness and surface area to volume. Cortex 82, 1–10 (2016).

68. Shaw, P. et al. Longitudinal mapping of cortical thickness and clinical outcome in children and adolescents with attention-deficit/hyperactivity disorder. Arch. Gen. Psychiatry 63, 540– 549 (2006).

69. Shaw, P. et al. Attention-deficit/hyperactivity disorder is characterized by a delay in cortical maturation. Proc. Natl. Acad. Sci. U. S. A. 104, 19649–19654 (2007).

70. Narr, K. L. et al. Widespread cortical thinning is a robust anatomical marker for attention-deficit/hyperactivity disorder. J. Am. Acad. Child Adolesc. Psychiatry 48, 1014–1022 (2009).

71. Hoogman, M. et al. Brain imaging of the cortex in ADHD: A coordinated analysis of large-scale clinical and population-based samples. Am. J. Psychiatry 176, 531–542 (2019).

72. Dall’Aglio, L. et al. Attention-deficit hyperactivity disorder symptoms and brain morphology: Examining confounding bias. Elife 11, (2022).

73. Bernanke, J. et al. Structural brain measures among children with and without ADHD in the Adolescent Brain and Cognitive Development Study cohort: a cross-sectional US population-based study. Lancet Psychiatry 9, 222–231 (2022).

74. Boedhoe, P. S. W. et al. Subcortical Brain Volume, Regional Cortical Thickness, and Cortical Surface Area Across Disorders: Findings From the ENIGMA ADHD, ASD, and OCD Working Groups. Am. J. Psychiatry 177, 834–843 (2020).

75. Shaw, P. et al. Mapping the development of the basal ganglia in children with attention-deficit/hyperactivity disorder. J. Am. Acad. Child Adolesc. Psychiatry 53, 780–789.e11 (2014).

76. Hoogman, M. et al. Subcortical brain volume differences of participants with ADHD across the lifespan: an ENIGMA collaboration. Lancet Psychiatry **di**, 1–39 (2017).

77. Emond, V., Joyal, C. & Poissant, H. Structural and functional neuroanatomy of attention-deficit hyperactivity disorder (ADHD). Encephale 35, 107–114 (2009).

78. Krain, A. L. & Castellanos, F. X. Brain development and ADHD. Clin. Psychol. Rev. 26, 433–444 (2006).

79. Rubia, K., Alegria, A. & Brinson, H. Imaging the ADHD brain: disorder-specificity, medication effects and clinical translation. Expert Rev. Neurother. 14, 519–538 (2014).

80. Adisetiyo, V. et al. Attention-deficit/hyperactivity disorder without comorbidity is associated with distinct atypical patterns of cerebral microstructural development. Hum. Brain Mapp. 35, 2148–2162 (2014).

81. Mahone, E. M. et al. Comprehensive examination of frontal regions in boys and girls with attention-deficit/hyperactivity disorder. J. Int. Neuropsychol. Soc. 17, 1047–1057 (2011).

82. Dirlikov, B. et al. Distinct frontal lobe morphology in girls and boys with ADHD. NeuroImage: Clinical 7, 222–229 (2014).

83. Seymour, K. E. et al. Anomalous subcortical morphology in boys, but not girls, with ADHD compared to typically developing controls and correlates with emotion dysregulation. Psychiatry Research - Neuroimaging 261, 20–28 (2017).

84. Jacobson, L. A. et al. Sex-Based Dissociation of White Matter Microstructure in Children With Attention-Deficit/Hyperactivity Disorder. J. Am. Acad. Child Adolesc. Psychiatry 54, 938–946 (2015).

85. Onnink, A. M. H. et al. Brain alterations in adult ADHD: effects of gender, treatment and comorbid depression. Eur. Neuropsychopharmacol. 24, 397–409 (2014).

86. Villemonteix, T. et al. Grey matter volume differences associated with gender in children with attention-deficit/hyperactivity disorder: A voxel-based morphometry study. Dev. Cogn. Neurosci. 14, 32–37 (2015).

87. Xavier Castellanos, F., et al. Developmental trajectories of brain volume abnormalities in children and adolescents with attention-deficit/hyperactivity disorder. J. Am. Med. Assoc. 288, 1740–1748 (2002).

88. Shaw, P. et al. Trajectories of cerebral cortical development in childhood and adolescence and adult attention-deficit/hyperactivity disorder. Biol. Psychiatry 74, 599–606 (2013).

89. Kaczkurkin, A. N., Raznahan, A. & Satterthwaite, T. D. Sex differences in the developing brain: insights from multimodal neuroimaging. Neuropsychopharmacology vol. 44 71–85 Preprint at 10.1038/s41386-018-0111-z (2019).

90. Baribeau, D. A. et al. Structural neuroimaging correlates of social deficits are similar in autism spectrum disorder and attention-deficit/hyperactivity disorder: analysis from the POND Network. Transl. Psychiatry 9, (2019).

91. Dougherty, C. C., Evans, D. W., Myers, S. M., Moore, G. J. & Michael, A. M. A Comparison of Structural Brain Imaging Findings in Autism Spectrum Disorder and Attention-Deficit Hyperactivity Disorder. Neuropsychology Review vol. 26 25–43 Preprint at 10.1007/s11065-015-9300-2 (2016).

92. Hoogman, M. et al. Consortium neuroscience of attention deficit/hyperactivity disorder and autism spectrum disorder: The ENIGMA adventure. Hum. Brain Mapp. 43, 37–55 (2022).

93. Ray, S. et al. Structural and functional connectivity of the human brain in autism spectrum disorders and attention-deficit/hyperactivity disorder: A rich club-organization study. Hum. Brain Mapp. 35, 6032–6048 (2014).

94. Bethlehem, R. A. I., Romero-Garcia, R., Mak, E., Bullmore, E. T. & Baron-Cohen, S. Structural covariance networks in children with autism or ADHD. Cereb. Cortex 27, 4267– 4276 (2017).

95. Kangarani-Farahani, M., Izadi-Najafabadi, S. & Zwicker, J. G. How does brain structure and function on MRI differ in children with autism spectrum disorder, developmental coordination disorder, and/or attention deficit hyperactivity disorder? Int. J. Dev. Neurosci. 82, 681–715 (2022).

96. Tung, Y.-H. et al. Whole Brain White Matter Tract Deviation and Idiosyncrasy From Normative Development in Autism and ADHD and Unaffected Siblings Link With Dimensions of Psychopathology and Cognition. Am. J. Psychiatry appiajp202020070999 (2021).

97. Berg, L. M. et al. The neuroanatomical substrates of autism and ADHD and their link to putative genomic underpinnings. Mol. Autism 14, 36 (2023).

98. Rommelse, N., Buitelaar, J. K. & Hartman, C. A. Structural brain imaging correlates of ASD and ADHD across the lifespan: a hypothesis-generating review on developmental ASD– ADHD subtypes. Journal of Neural Transmission vol. 124 259–271 Preprint at 10.1007/s00702-016-1651-1 (2017).

99. Chen, Y. et al. Altered cortical gyrification, sulcal depth, and fractal dimension in the autism spectrum disorder comorbid attention-deficit/hyperactivity disorder than the autism spectrum disorder. Neuroreport 34, 93–101 (2023).

100. Misaki, M., Wallace, G. L., Dankner, N., Martin, A. & Bandettini, P. A. Characteristic cortical thickness patterns in adolescents with autism spectrum disorders: Interactions with age and intellectual ability revealed by canonical correlation analysis. Neuroimage 60, 1890–1901 (2012).

101. Raznahan, A. et al. Cortical anatomy in autism spectrum disorder: An in vivo MRI study on the effect of age. Cereb. Cortex 20, 1332–1340 (2010).

102. Wallace, G. L. et al. Longitudinal cortical development during adolescence and young adulthood in autism spectrum disorder: increased cortical thinning but comparable surface area changes. J. Am. Acad. Child Adolesc. Psychiatry 54, 464–469 (2015).

103. Bethlehem, R. A. I. et al. Brain charts for the human lifespan. Nature 604, 525–533 (2022).

104. Marquand, A. F., Rezek, I., Buitelaar, J. & Beckmann, C. F. Understanding Heterogeneity in Clinical Cohorts Using Normative Models: Beyond Case-Control Studies. Biol. Psychiatry 80, 552–561 (2016).

105. Wolfers, T. et al. Mapping the Heterogeneous Phenotype of Schizophrenia and Bipolar Disorder Using Normative Models. JAMA Psychiatry 75, 1146–1155 (2018).

106. Bedford, S. A., Seidlitz, J. & Bethlehem, R. A. I. Translational potential of human brain charts. Clinical and translational medicine vol. 12 e960 (2022).

107. Di Martino, A. et al. The autism brain imaging data exchange: towards a large-scale evaluation of the intrinsic brain architecture in autism. Mol. Psychiatry 19, 659–667 (2014).

108. Di Martino, A. et al. Enhancing studies of the connectome in autism using the autism brain imaging data exchange II. Sci Data 4, 170010 (2017).

109. Alexander, L. M. et al. An open resource for transdiagnostic research in pediatric mental health and learning disorders. Sci Data 4, 170181 (2017).

110. Fischl, B. & Dale, A. M. Measuring the thickness of the human cerebral cortex from magnetic resonance images. Proc. Natl. Acad. Sci. U. S. A. 97, 11050–11055 (2000).

111. Desikan, R. S. et al. An automated labeling system for subdividing the human cerebral cortex on MRI scans into gyral based regions of interest. (2006) doi:10.1016/j.neuroimage.2006.01.021.

112. Bedford, S. A. et al. The impact of quality control on cortical morphometry comparisons in autism. Imaging Neuroscience 1, 1–21 (2023).

113. Pardoe, H. R., Kucharsky Hiess, R. & Kuzniecky, R. Motion and morphometry in clinical and nonclinical populations. Neuroimage 135, 177–185 (2016).

114. Reuter, M. et al. Head motion during MRI acquisition reduces gray matter volume and thickness estimates. Neuroimage 107, 107–115 (2015).

115. Rosen, A. F. G. et al. Quantitative assessment of structural image quality. Neuroimage 169, 407–418 (2018).

116. Fortin, J. P. et al. Harmonization of cortical thickness measurements across scanners and sites. Neuroimage 167, 104–120 (2018).

117. Schabdach, J. M. et al. Brain growth charts of ‘clinical controls’ for quantitative analysis of clinically acquired brain MRI. bioRxiv (2023) doi:10.1101/2023.01.13.23284533.

118. Lai, M. C. et al. Imaging sex/gender and autism in the brain: Etiological implications. J. Neurosci. Res. 95, 380–397 (2017).

119. Lee, J. K. et al. Altered Development of Amygdala-Connected Brain Regions in Males and Females with Autism. J. Neurosci. 42, 6145–6155 (2022).

120. Alexander-Bloch, A. F. et al. On testing for spatial correspondence between maps of human brain structure and function. Neuroimage 178, 540–551 (2018).

121. Benjamini, Y. & Hochberg, Y. Controlling the false discovery rate: a practical and powerful approach to multiple testing. J. R. Stat. Soc. Series B Stat. Methodol. 57, 289–300 (1995).

122. Maechler, M. Package ‘diptest’. https://cran.r-project.org/web/packages/diptest/diptest.pdf (2022).

123. Swanson, J. M. et al. Categorical and Dimensional Definitions and Evaluations of Symptoms of ADHD: History of the SNAP and the SWAN Rating Scales. Int J Educ Psychol Assess 10, 51–70 (2012).

124. Wolfers, T. et al. Individual differences v. the average patient: mapping the heterogeneity in ADHD using normative models. Psychol. Med. 50, 314–323 (2020).

125. Yankowitz, L. D. et al. Evidence against the ‘normalization’ prediction of the early brain overgrowth hypothesis of autism. Mol. Autism 11, 1–17 (2020).

126. Prigge, M. B. D. et al. A 16-year study of longitudinal volumetric brain development in males with autism. Neuroimage 236, 118067 (2021).

127. Denier, N., Steinberg, G., van Elst, L. T. & Bracht, T. The role of head circumference and cerebral volumes to phenotype male adults with autism spectrum disorder. Brain Behav. 12, e2460 (2022).

128. Shen, M. D. Cerebrospinal fluid and the early brain development of autism. J. Neurodev. Disord. 10, 39 (2018).

129. Shiohama, T. et al. Small Nucleus Accumbens and Large Cerebral Ventricles in Infants and Toddlers Prior to Receiving Diagnoses of Autism Spectrum Disorder. Cereb. Cortex 32, 1200–1211 (2022).

130. van Rooij, D. et al. Cortical and Subcortical Brain Morphometry Differences Between Patients With Autism Spectrum Disorder and Healthy Individuals Across the Lifespan: Results From the ENIGMA ASD Working Group. Am. J. Psychiatry 175, 359–369 (2018).

131. Kyriakopoulou, V. et al. Characterisation of ASD traits among a cohort of children with isolated fetal ventriculomegaly. Nat. Commun. 14, 1550 (2023).

132. Ecker, C. et al. Interindividual Differences in Cortical Thickness and Their Genomic Underpinnings in Autism Spectrum Disorder. Am. J. Psychiatry 179, 242–254 (2022).

133. Kobayashi, A. et al. Increased grey matter volume of the right superior temporal gyrus in healthy children with autistic cognitive style: A VBM study. Brain Cogn. 139, 105514 (2020).

134. Jou, R. J., Minshew, N. J., Keshavan, M. S., Vitale, M. P. & Hardan, A. Y. Enlarged right superior temporal gyrus in children and adolescents with autism. Brain Res. 1360, 205–212 (2010).

135. Xiao, Y. et al. Atypical functional connectivity of temporal cortex with precuneus and visual regions may be an early-age signature of ASD. Mol. Autism 14, 11 (2023).

136. Kim, D. et al. Overconnectivity of the right Heschl’s and inferior temporal gyrus correlates with symptom severity in preschoolers with autism spectrum disorder. Autism Res. 14, 2314–2329 (2021).

137. Alaerts, K. et al. Underconnectivity of the superior temporal sulcus predicts emotion recognition deficits in autism. Soc. Cogn. Affect. Neurosci. 9, 1589–1600 (2014).

138. Chung, S. & Son, J.-W. Visual Perception in Autism Spectrum Disorder: A Review of Neuroimaging Studies. Soa Chongsonyon Chongsin Uihak 31, 105–120 (2020).

139. Wang, H. et al. Developmental brain structural atypicalities in autism: a voxel-based morphometry analysis. Child Adolesc. Psychiatry Ment. Health 16, 7 (2022).

140. Nickl-Jockschat, T. et al. Brain structure anomalies in autism spectrum disorder--a meta-analysis of VBM studies using anatomic likelihood estimation. Hum. Brain Mapp. 33, 1470– 1489 (2012).

141. Zoltowski, A. R. et al. Cortical Morphology in Autism: Findings from a Cortical Shape-Adaptive Approach to Local Gyrification Indexing. Cereb. Cortex 31, 5188–5205 (2021).

142. Zilbovicius, M., Saitovitch, A., Popa, T., Rechtman, E. & Boddaert, N. Autism, social cognition and superior temporal sulcus. Open Journal of Psychiatry 3, 46–55 (2013).

143. Baum, S. H., Stevenson, R. A. & Wallace, M. T. Behavioral, perceptual, and neural alterations in sensory and multisensory function in autism spectrum disorder. Prog. Neurobiol. 134, 140–160 (2015).

144. Lombardo, M. V. et al. Different functional neural substrates for good and poor language outcome in autism. Neuron 86, 567–577 (2015).

145. Lombardo, M. V. et al. Large-scale associations between the leukocyte transcriptome and BOLD responses to speech differ in autism early language outcome subtypes Europe PMC Funders Group. Nat. Neurosci. 21, 1680–1688 (2018).

146. Xiao, Y. et al. Neural responses to affective speech, including motherese, map onto clinical and social eye tracking profiles in toddlers with ASD. Nat Hum Behav 6, 443–454 (2022).

147. Alexander-Bloch, A. et al. Subtle in-scanner motion biases automated measurement of brain anatomy from in vivo MRI. Hum. Brain Mapp. 2397, 2385–2397 (2016).

148. Nakua, H. et al. Systematic comparisons of different quality control approaches applied to three large pediatric neuroimaging datasets. Neuroimage 274, 120119 (2023).

149. Swanson, M. R. et al. Subcortical Brain and Behavior Phenotypes Differentiate Infants with Autism versus Language Delay for the IBIS Network, Subcortical Brain and Behavior Phenotypes Differentiate Infants with Autism versus Language Delay. Biological Psychiatry: Cognitive Neuroscience and Neuroimaging (2017) doi:10.1016/j.bpsc.2017.07.007.

150. Zhang, W., et al. Revisiting subcortical brain volume correlates of autism in the ABIDE dataset: effects of age and sex. Psychol. Med. (2017) doi:10.1017/S003329171700201X.

151. Schuetze, M. et al. Morphological Alterations in the Thalamus, Striatum, and Pallidum in Autism Spectrum Disorder. Neuropsychopharmacology 41, 2627–2637 (2016).

152. Panizzon, M. S. et al. Distinct genetic influences on cortical surface area and cortical thickness. Cereb. Cortex 19, 2728–2735 (2009).

153. Strike, L. T. et al. Genetic Complexity of Cortical Structure: Differences in Genetic and Environmental Factors Influencing Cortical Surface Area and Thickness. Cereb. Cortex 29, 952–962 (2019).

154. Grasby, K. L. et al. The genetic architecture of the human cerebral cortex. Science 367, (2020).

155. Rakic, P. Specification of cerebral cortical areas. Science 241, 170–176 (1988).

156. Grotzinger, A. D. et al. Multivariate genomic architecture of cortical thickness and surface area at multiple levels of analysis. Nat. Commun. 14, 946 (2023).

157. Chen, C.-H. et al. Hierarchical genetic organization of human cortical surface area. Science 335, 1634–1636 (2012).

158. Chen, C.-H. et al. Genetic topography of brain morphology. Proc. Natl. Acad. Sci. U. S. A. 110, 17089–17094 (2013).

159. Makowski, C. et al. Discovery of genomic loci of the human cerebral cortex using genetically informed brain atlases. Science 375, 522–528 (2022).

160. Warrier, V. et al. Genetic insights into human cortical organization and development through genome-wide analyses of 2,347 neuroimaging phenotypes. Nat. Genet. 55, 1483– 1493 (2023).

161. Winkler, A. M. et al. Cortical thickness or grey matter volume? The importance of selecting the phenotype for imaging genetics studies. Neuroimage 53, 1135–1146 (2010).

162. Cadwell, C. R., Bhaduri, A., Mostajo-Radji, M. A., Keefe, M. G. & Nowakowski, T. J. Development and Arealization of the Cerebral Cortex. Neuron 103, 980–1004 (2019).

163. Li, M. et al. Integrative functional genomic analysis of human brain development and neuropsychiatric risks. Science 362, (2018).

164. Gandal, M. J. et al. Broad transcriptomic dysregulation occurs across the cerebral cortex in ASD. Nature 611, 532–539 (2022).

165. Bölte, S. et al. Sex and gender in neurodevelopmental conditions. Nat. Rev. Neurol. 19, 136–159 (2023).

166. Tunç, B. et al. Deviation from normative brain development is associated with symptom severity in autism spectrum disorder. Mol. Autism 10, (2019).

167. Courchesne, E. Brain development in autism: early overgrowth followed by premature arrest of growth. Ment. Retard. Dev. Disabil. Res. Rev. 10, 106–111 (2004).

168. Mahajan, R., Dirlikov, B., Crocetti, D. & Mostofsky, S. H. Motor Circuit Anatomy in Children with Autism Spectrum Disorder With or Without Attention Deficit Hyperactivity Disorder. Autism Res. 9, 67–81 (2016).

169. Nickel, K. et al. Inferior Frontal Gyrus Volume Loss Distinguishes Between Autism and (Comorbid) {Attention-Deficit/Hyperactivity} {Disorder-A} {FreeSurfer} Analysis in Children. Front. Psychiatry 9, 521 (2018).

170. Rong, Y., Yang, C.-J., Jin, Y. & Wang, Y. Prevalence of attention-deficit/hyperactivity disorder in individuals with autism spectrum disorder: A meta-analysis. Res. Autism Spectr. Disord. 83, 101759 (2021).

