## supplementary_materials for "Brain-charting autism and attention deficit hyperactivity disorder reveals distinct and overlapping neurobiology"

**Supplementary methods	1**

1. Dataset characteristics	1

1.1 Study and site details and parameters	1

1.2 Sample demographics and N before and after quality control (QC)	3

Table S1.2.1: Participant demographics by study (before QC)	3

Table S1.2.2: Participant demographics by study (post QC)	5

2. Sensitivity analysis: controlling for GMV, total SA and mean CT	6

3. Sensitivity analyses: different QC methods	7

4. Sensitivity analysis on subsample of matched male data	7

5. Multimodality of distributions by diagnostic group	7

6. Age interaction on subsample of age matched controls	7

7. Dimensional analyses of autistic and ADHD traits	8

7.1 Descriptions of each measure	8

7.2 Subsample demographic details	9

7.3 Sensitivity analysis using SRS T-scores	9

8. Correlation and overlap of brain maps between diagnostic groups	10

**Supplementary results	11**

1. Controlling for GMV, total SA and mean CT	11

2. Sensitivity analysis with varying QC methods	12

3. Spatial comparison of male and female diagnosis effect size maps	15

4. Sensitivity analysis on subsample of matched male data	16

5. Multimodality of data	17

6. Age interaction on subsample of age-matched controls	18

7. Dimensional analyses of autistic traits	19

7.1 Distribution of scores by group	19

7.2 Relationship by group in significant regions	20

7.3 Sensitivity analysis using SRS T-scores	21

8. Correlation and overlap of brain maps between diagnostic groups	21

### Supplementary methods

#### 1. Dataset characteristics

##### 1.1 Study and site details and parameters

**The Autism Brain Imaging Data Exchange (ABIDE)**

The ABIDE dataset (Di Martino et al, 2014) is a large, publicly available neuroimaging dataset of individuals with autism and controls, collected at multiple sites across Europe and North America. ABIDE contains two data releases (ABIDE I and II), amounting to a total of 2145 participants (1017 individuals with ASD (137 female/880 male) and 1128 controls (274 female/854 male)), aged 5-64 years. For more information about individual sites and imaging protocols, please see: <http://fcon_1000.projects.nitrc.org/indi/abide/>.

**The Province of Ontario Neurodevelopmental (POND) Network**

POND is an integrated discovery program based in Ontario, Canada aimed at improving our understanding of neurodevelopmental disorders and their underlying neurobiology. POND includes neuroimaging, clinical, behavioural and genetic data across multiple time points in children with neurodevelopmental diagnoses, including ADHD, autism, intellectual disability, obsessive compulsive disorder and Tourette syndrome, amongst others, from five sites across Ontario (Holland Bloorview Kids Rehabilitation Hospital, Toronto; The Hospital for Sick Children, Toronto; McMaster Children’s Hospital, Hamilton; Queen’s University, Kingston; and Lawson Health Research Institute, London). The majority of scans were collected on a 3T Siemens Trio TIM scanner, with a hardware upgrade to the Siemens Prisma scanner affecting the rest of the data. For the current project, data from participants with a diagnosis of autism and ADHD, as well as controls, were used. Neuroimaging data was available for 736 of these participants (179 TD/244 ADHD/313 autistic). Initially, the different POND data collection sites were treated as separate sites for the purposes of this study. However, some of these sites did not have any control participants, which is a necessity for running the out-of-sample centile models. Thus, ComBat was run on all data with the sites separated, and they were recombined after ComBat harmonisation and treated as one POND site for the out-of-sample centile estimation. For more details on the POND network, please see: <https://pond-network.ca/>.

**The Healthy Brain Network (HBN) at the Child Mind Institute (CMI)**

The HBN is an initiative based at the Child Mind Institute, in New York City, USA, aiming to create a biobank of data from children and adolescents, with the goal of advancing our understanding of childhood and adolescent psychiatric and neurodevelopmental disorders. Participants are referred by clinicians in the community, and thus are primarily children presenting with psychiatric symptoms or concerns, but may not all have an official diagnosis. Data collected includes clinical, behavioural and cognitive measures, as well as genetic information and neuroimaging, collected at three sites in and around New York City. Data included in the current project includes participants diagnosed with autism and ADHD, and controls. Neuroimaging data is available from 1592 participants (308 autism/149 ADHD/100 controls). Data from the HBN comes from three sites: (Rutgers University Brain Imaging Center (RU), Staten Island (SI), CitiGroup Cornell Brain Imaging Center (CBIC)), and were treated as three separate sites at every stage of analysis. Imaging data was collected using Siemens 3T TrioTim scanner with a 32-channel head coil at RU and SI, and a Siemens 3T Prisma scanner with a 32-channel head coil at CBIC, all using an MPRAGE sequence. For more details please see: <http://fcon_1000.projects.nitrc.org/indi/cmi_healthy_brain_network/>.

**The ADHD200 Consortium**

The publicly available ADHD200 Consortium consists of neuroimaging and phenotypic data from individuals with ADHD and controls across 8 sites internationally. A total of 947 children and adolescents are included in the dataset (362 ADHD/585 controls). For more information about individual sites and imaging protocols, please see: <http://fcon_1000.projects.nitrc.org/indi/adhd200/>.

**Multimodal Developmental Neurogenetics of Females with ASD (Female ASD)**

The Female ASD dataset, publicly available as part of the NIMH NDAR initiative (ID 2021), is the product of a collaboration between Yale University, the University of California Los Angeles, Harvard University, and the University of Washington, which aims specifically to investigate the underlying neurobiology and genetics of autism in females. The dataset comprises phenotypic, clinical, neuroimaging and genetic data from 397 children and adolescents with autism and controls (183 autism/ 214 control) with a good representation of female participants. Imaging data was collected on a Siemens 3T TrioTim or Prisma scanner at all sites. At two sites, scanners were upgraded to Prisma part way through data collection; these have thus been split up and counted as separate sites per scanner.

**University of California San Diego (UCSD): Biomarkers of Autism at 12 Months: From Brain Overgrowth to Genes**

The UCSD biomarkers of autism dataset, publicly available as part of the NIMH NDAR initiative, includes phenotypic, clinical and neuroimaging data from infants age 12-36 months who are at risk of developing autism, language delay, or developmental delay, as well as typical controls. Data is collected at multiple time points, with diagnoses tracked across visits. Here, we used only the first time point for each participant. Imaging data was collected on a General Electric Signa HDxt 1.5T scanner while the infants were sleeping.

**The UK Medical Research Council (MRC) Autism Imaging Multi-centre Study (AIMS)**

The UK MRC-AIMS dataset is a multi-site initiative aiming to improve our understanding of females with autism. The dataset consists of clinical and neuroimaging data from a sex balanced sample of autistic and non-autistic adults, collected at the University of Cambridge, the University of Oxford and the Institute of Psychiatry, Psychology and Neuroscience (IoPPN) at King’s College London. In the current study, only data from the University of Cambridge site was used, which included 61 autistic adults (31 female/30 male) and 63 controls (31 female/32 male), aged 18-52. Imaging data was collected using a 3T General Electrical Medical Systems HDx scanner with an 8-channel receive-only head coil. For details see [[1–3]](https://paperpile.com/c/jj3h1Z/5l2r+KQ6k+ZOHT).

##### 1.2 Sample demographics and N before and after quality control (QC)

Before quality control, our sample included data from 4736 participants (2006 typically developing (TD) controls [1289 male/717 female], 1103 individuals with ADHD [809 male; 294 female], and 1627 autistic individuals [1296 male; 331 female]), with an age range of 2-64 (mean age = 13.67; median age = 11.98).

##### Table S1.2.1: Participant demographics by study (before QC)

| **Study** | **Group** | **N** | **Median age** | **Age range** |
| --- | --- | --- | --- | --- |
| ADHD200 | ADHD Female | 72 | 9.7 | 7.3-19.7 |
|  | ADHD Male | 230 | 11.3 | 7.2-20.9 |
|  | TD Female | 230 | 11.6 | 7.1-21.8 |
|  | TD Male | 263 | 11.8 | 7.2-25 |
| CMI HBN | ADHD Female | 189 | 9.4 | 5-21.7 |
|  | ADHD Male | 460 | 9.7 | 5.1-20.3 |
|  | Autistic Female | 19 | 10.4 | 5.7-16 |
|  | Autistic Male | 86 | 10.7 | 5.7-20.4 |
|  | TD Female | 73 | 9.6 | 5.5-19.5 |
|  | TD Male | 72 | 9.1 | 5-21.2 |
| Female ASD | Autistic Female | 85 | 11.9 | 8-18 |
|  | Autistic Male | 98 | 12 | 8-17.9 |
|  | TD Female | 113 | 12.4 | 8.1-17.9 |
|  | TD Male | 101 | 13 | 8-17.8 |
| POND | ADHD Female | 72 | 11.1 | 6.2-21.5 |
|  | ADHD Male | 208 | 11.2 | 5.8-21.9 |
|  | Autistic Female | 82 | 11.5 | 2.6-22 |
|  | Autistic Male | 316 | 11.3 | 2.2-22 |
|  | TD Female | 88 | 12.7 | 3.6-21.4 |
|  | TD Male | 99 | 11.2 | 3.7-21.7 |
| UCSD | Autistic Female | 18 | 3.4 | 3.1-4.5 |
|  | Autistic Male | 76 | 3.3 | 3-5.2 |
|  | TD Female | 15 | 3.4 | 3-3.8 |
|  | TD Male | 32 | 3.4 | 3-5 |
| UK AIMS | Autistic Female | 31 | 24.7 | 18.1-49.5 |
|  | Autistic Male | 30 | 24.5 | 18-41 |
|  | TD Female | 31 | 25.8 | 19-45.1 |
|  | TD Male | 32 | 26 | 18-42 |
| ABIDE I | Autistic Female | 62 | 14 | 8.1-45 |
|  | Autistic Male | 448 | 15 | 7-64 |
|  | TD Female | 99 | 13.8 | 7.8-46 |
|  | TD Male | 461 | 15.3 | 6.5-56.2 |
| ABIDE II | Autistic Female | 56 | 11.7 | 5.2-54 |
|  | Autistic Male | 328 | 12.7 | 5.1-62 |
|  | TD Female | 113 | 11 | 5.9-46.6 |
|  | TD Male | 278 | 13.8 | 5.9-64 |

##### Table S1.2.2: Participant demographics by study (post QC)

| **Study** | **Group** | **N** | **Median age** | **Age range** | **Attrition (% lost to QC)** |
| --- | --- | --- | --- | --- | --- |
| ADHD200 | ADHD Female | 61 | 9.8 | 7.3-19.7 | 15.28 |
|  | ADHD Male | 198 | 11.3 | 7.2-20.9 | 13.91 |
|  | TD Female | 197 | 11.5 | 7.1-21.8 | 14.35 |
|  | TD Male | 231 | 11.8 | 7.2-25 | 12.17 |
| CMI HBN | ADHD Female | 147 | 10 | 5-21.7 | 22.22 |
|  | ADHD Male | 337 | 10.1 | 5.3-20.3 | 26.74 |
|  | Autistic Female | 14 | 11.7 | 5.7-16 | 26.32 |
|  | Autistic Male | 67 | 11.2 | 5.7-20.4 | 22.09 |
|  | TD Female | 66 | 9.7 | 5.6-19.5 | 9.59 |
|  | TD Male | 63 | 10.2 | 5-21.2 | 12.5 |
| Female ASD | Autistic Female | 75 | 12.5 | 8.2-18 | 11.76 |
|  | Autistic Male | 78 | 13.1 | 8.3-17.9 | 20.41 |
|  | TD Female | 109 | 12.6 | 8.1-17.9 | 3.54 |
|  | TD Male | 92 | 13 | 8-17.8 | 8.91 |
| POND | ADHD Female | 62 | 11.2 | 6.2-21.5 | 13.89 |
|  | ADHD Male | 182 | 11.3 | 6.4-21.9 | 12.5 |
|  | Autistic Female | 67 | 11.9 | 2.6-22 | 18.29 |
|  | Autistic Male | 246 | 11.6 | 2.2-22 | 22.15 |
|  | TD Female | 87 | 12.8 | 3.6-22 | 1.14 |
|  | TD Male | 92 | 11.5 | 3.7-21.7 | 7.07 |
| UCSD | Autistic Female | 18 | 3.4 | 3.1-4.5 | 0 |
|  | Autistic Male | 76 | 3.3 | 3-5.2 | 0 |
|  | TD Female | 15 | 3.4 | 3-3.8 | 0 |
|  | TD Male | 31 | 3.4 | 3-5 | 0 |
| UK AIMS | Autistic Female | 30 | 25.9 | 18.1-49.5 | 3.23 |
|  | Autistic Male | 28 | 25 | 18-41 | 6.67 |
|  | TD Female | 28 | 26 | 19-45.1 | 9.68 |
|  | TD Male | 32 | 26 | 18-42 | 0 |
| ABIDE I | Autistic Female | 39 | 16.5 | 9.3-45 | 37.1 |
|  | Autistic Male | 333 | 16.9 | 7.1-64 | 25.67 |
|  | TD Female | 80 | 14.2 | 8-46 | 19.19 |
|  | TD Male | 392 | 16.1 | 6.5-56.2 | 14.97 |
| ABIDE II | Autistic Female | 45 | 12 | 5.2-54 | 19.64 |
|  | Autistic Male | 283 | 13 | 5.1-62 | 13.72 |
|  | TD Female | 105 | 12 | 5.9-46.6 | 7.08 |
|  | TD Male | 249 | 15 | 5.9-64 | 10.43 |

#### 2. Sensitivity analysis: controlling for GMV, total SA and mean CT

In the main paper analyses, we chose not to control for total brain volume or global brain measures in our analyses, due to concerns that controlling for global measures would obscure sex differences, which was one of our primary areas of interest. However, in light of lack of consensus about whether to control for total brain volume or global measures in analyses or regional cortical measures, we have also re-run these analyses while controlling for the respective global measure: total grey matter volume, total surface area, or mean cortical thickness. Analyses were identical to the main analyses for group differences, sex by diagnosis interaction, and sex-stratified analyses, other than the addition of these measures in the model.

#### 3. Sensitivity analyses: different QC methods

All interaction and main effect analyses were rerun using various combinations and methods of accounting for QC, including using FSQC only as a cut off (N = 4255; without controlling for Euler number), using Euler number only as a cut off (N = 4367), and using both FSQC and Euler number as a cut off (N = 4015). The main analyses were run in exactly the same way with the exception of these changes in QC method, to assess the impact of QC procedure on results. The cut off point for Euler number was at 2 median absolute deviations, which was a Euler number of 254.

#### 4. Sensitivity analysis on subsample of matched male data

Due to the imbalance in sample size between males and females, we wanted to assess the extent to which apparent sex differences in cortical alterations related to autism and ADHD in the interaction and sex stratified analyses may be attributable to differences in power. Thus, we re-ran these analyses using a subset of the male data, that was equal in size and matched to the female sample. To do this, we used the MatchIt package in R for propensity matching between the male and female samples, based on age, diagnostic group and site. This resulted in a total sample of 2896, with 1448 each of males (754 TD/330 ADHD/364 autistic) and females (762 TD/333 ADHD/353 autistic). The main analyses were then re-run on this sample, including main effect of diagnosis, interaction with sex, and the sex stratified analyses. Correlations were run on the subthreshold effect size maps between these and the original analyses on the full sample to assess consistency of results.

#### 5. Multimodality of distributions by diagnostic group

To account for the fact that neurodevelopmental conditions are heterogeneous and may comprise multiple subgroups, we assessed multimodality of the centile scores in each diagnostic group. We explored the number of peaks in each probability density function using the *density* function in R [[4]](https://paperpile.com/c/jj3h1Z/HLkV) using a Gaussian smoothing kernel. The number of peaks was derived from the inflection points of the density curves. Multimodality was tested using Hartigan’s dip test [[5]](https://paperpile.com/c/jj3h1Z/ZpWZ), which provides a p-value for the alternate hypothesis that the data are multimodal. Distributions were examined separately for each diagnostic group, and for each cortical phenotype and region separately. Significance and number of peaks for each parcellation were displayed using ggseg in R [[6]](https://paperpile.com/c/jj3h1Z/4Jt6). Density plots were generated using the geom_flat_violin option in the PupillometryR package [[7]](https://paperpile.com/c/jj3h1Z/cBlO).

#### 6. Dimensional analyses of autistic and ADHD traits

We investigated dimensional associations between neuroanatomical measures and autistic and ADHD traits across all three groups, in subsets of the data with these measures available. For autistic traits, we used the Autism Diagnostic Observation Schedule calibrated severity score (ADOS CSS; N = 19 TD/13 ADHD/732 autistic), the Restricted Behaviour Scale Revised (RBS-R; N = 442 TD/456 ADHD/594 autistic), the Social Responsiveness Scale (SRS; N = 702 TD/372 ADHD/602 autistic) total raw scores (analyses were also run using scaled T-scores for comparison). For ADHD measures, we used the Strengths and Weaknesses of Attention-Deficit/Hyperactivity-symptoms and Normal-behaviours ^121^ (SWAN; N = 200 TD/397 ADHD/358 autistic) inattentive and hyperactivity/impulsivity subscale scores. Details of each measure and subsample are described below. Linear models examined the relationship between each behavioural measure and each regional cortical phenotype, in separate models, controlling for age, sex, and Euler number. We repeated each analysis controlling for global brain measures. Given that significant sex differences were observed for each measure, sex-by-trait interactions were also examined.

##### 6.1 Descriptions of each measure

**ADOS.** The Autism Diagnostic Observation Schedule (ADOS [[8]](https://paperpile.com/c/jj3h1Z/FCsIS)) is the gold standard instrument for diagnosing autism and assessing autistic symptoms. The ADOS is a semi-structured interview and series of activities, which differs depending on the age, developmental and language level of the individual. The ADOS 2 allows for the calculation of a standardised score across modules (calibrated severity score; CSS). This score quantifies autistic traits (independent of age and IQ), allowing for the comparison and pooling of scores across modules, which would not otherwise be possible, and is useful for research purposes. CSS scores can also be calculated from ADOS 1 scores if item level data is available, according to the algorithm by Hus and colleagues [[9]](https://paperpile.com/c/jj3h1Z/4Gk5K). This was done in the current dataset using the methods implemented by Hammill and colleagues [[10]](https://paperpile.com/c/jj3h1Z/m2sr).

**RBS-R.** The Repetitive Behaviour Scale-Revised (RBS-R) is a parent-report measure designed to comprehensively assess the range of restricted, repetitive behaviours characteristic of autism. The RBS-R provides an overall total, as well as 6 subscales reflecting different domains of restricted, repetitive behaviours: Stereotyped Behavior, Self-injurious Behaviour, Compulsive Behaviour, Routine Behaviour, Sameness Behavior, and Restricted Behavior (with no overlapping content). Each item is assessed in the form of a 4-point Likert scale from 0 (behaviour does not occur) to 3 (behaviour occurs and is a severe problem). A 5 subscale scoring system for the RBS-R also exists, based on factor analysis [[11]](https://paperpile.com/c/jj3h1Z/s1JwX). The 6 subscale version was provided for the majority of datasets, and as this was also the original conceptualisation of the measure, this version was used. For HBN however, the 5 subscale version was provided. The original 6 subscales were recalculated using the raw data from HBN, and the scoring instructions from the original paper by Bodfish and colleagues [[12]](https://paperpile.com/c/jj3h1Z/gK8Le).

**SRS.** The Social Responsiveness Scale (SRS [[13]](https://paperpile.com/c/jj3h1Z/7xBrz)) is a 65 item self- or parent- report measure assessing the presence and level of social difficulty related to autism, as well as RRBs. The SRS captures even subtle traits, and can be used as a screener for autism, as well as to assess variation in autistic traits in the general population. There are two total scores for the SRS: the raw score, which is simply the sum of the score for each question, and the T-score, which is scaled for the respondent’s gender and age. A total T-score of 76 or higher is strongly associated with a diagnosis of autism; and T-scores of 59 and below are considered to not be associated with clinically significant autism.

**SWAN.** The Strengths and Weaknesses of Attention-Deficit/Hyperactivity-symptoms and Normal-behaviours [[14]](https://paperpile.com/c/jj3h1Z/8wkEI) (SWAN) is an 18-item parent report measures of ADHD-related behaviours for children, focusing on both positive and negative behaviours. The SWAN contains two subscales: the Hyperactive/Impulsive scale and the Inattentive scale, which each comprise 9 questions. Questions are rated on a scale of -3 - 3 (far below average; below average; slightly below average; average; slightly above average; above average; far above average). Questions rated -2 or below (below or far below average) are scored as one point, and if a child gets 6 or more points on either subscale, they are deemed to have met the clinical cut-off for ADHD, and considered likely to have that ADHD subtype. It has been widely used in studies of genetics, imaging, neuropsychology, and co-occurring diagnosis, among others [[15]](https://paperpile.com/c/jj3h1Z/lz17).

##### 6.2 Subsample demographic details

**ADOS.** In the current dataset, ADOS CSS was available (or recoded from item level data) for 764 individuals (732 autistic participants, 13 individuals with ADHD, and 19 TD individuals) post QC. Age distributions for each group are seen in figure S6.2.1.

**RBS-R.** The RBS-R was available from 1492 in our data set (post-QC; 456 ADHD, 594 autistic, 442 TD). Age distributions for each group are seen in figure S6.2.1.

**SRS.** In our dataset, the SRS was available for 1676 participants (post-QC; 372 ADHD, 602 autistic individuals and 702 TD). Age distributions for each group are seen in figure S6.2.1.

**SWAN.** The SWAN was available for 801 participants (post QC; 326 ADHD, 286 autistic individuals and 189 TD). Age distributions for each group are seen in figure S6.2.1.

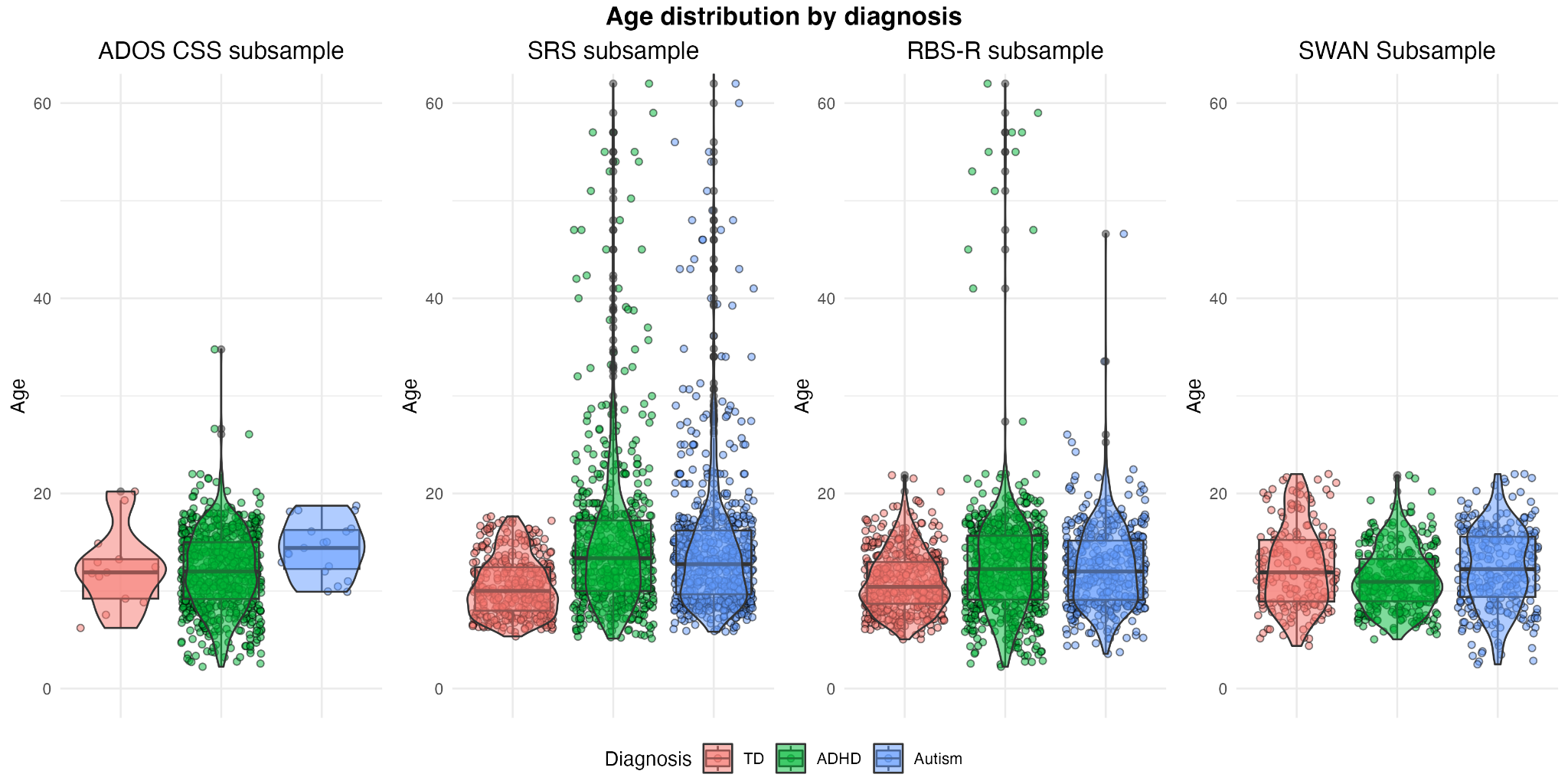

**Figure S.6.2.1.** Age distribution by diagnosis for each measure subsample.

##### 6.3 Sensitivity analysis using SRS T-scores

Raw total scores were used for the main analysis examining the association between SRS score and cortical measures so as not to obscure any potential sex differences. However, as there is not a clear consensus about which score is best to use for research purposes, we also conducted the same analysis using the SRS total T-score for comparison. The analyses were conducted in exactly the same manner as those in the “Associations with autistic traits” section in the main paper, except replacing the raw total score for total T-score.

#### 7. Age interaction on subsample of age matched controls

We observed a significant age by sex interaction for ADHD diagnosis; however, to assess to what extent this interaction was driven by the uneven age range in the ADHD (5-22) and control groups, we re-ran this analysis in a subset of controls with the same age range. Any TD participants below the age of 5 and over the age of 22 were excluded, and the identical analysis was then run.

#### 8. Autism+ADHD comparison and replication analyses

##### 8.1 Correlation and overlap of brain maps between diagnostic groups

To examine similarity and overlap between the autism, ADHD and autism+ADHD groups, we performed pairwise spearman correlations between the effect sizes across brain regions, for each cortical phenotype. Correlations were plotted for each cortical phenotype and each pairwise comparison. Brain regions were assigned to a lobe based on the division in [[16]](https://paperpile.com/c/jj3h1Z/p74p), and colour coded according to their corresponding lobe in the correlation plots, in order to visualise whether some lobes show more similar effects across conditions. Next, in order to visualise these effects across the cortex, we plotted the pairwise overlap of effect size direction and significance in ggseg [[6]](https://paperpile.com/c/jj3h1Z/4Jt6). For each pair, regions were selected that had effect sizes in the same direction in both conditions (ie. positive or negative), and colour coded accordingly. We also identified regions that displayed a positive effect in both conditions. Spin permutation testing was used to assess significance with appropriate control of the potentially confounding effect of spatial autocorrelation of each map ^118^.

##### 8.2 Sensitivity analysis controlling for GMV, total SA and mean CT

We have also re-run these analyses for the autism+ADHD group while controlling for the respective global measure: total grey matter volume, total surface area, or mean cortical thickness. Analyses were identical to the main analyses for group differences, sex by diagnosis interaction, and sex-stratified analyses, other than the addition of these measures in the model.

##### 8.3 Replication in subset based on dimensional clinical measures

Secondary diagnosis data was not available for all datasets, and can be unreliable; thus there are likely many individuals with co-occurring diagnoses who were therefore not included in this analysis. Due to this, we attempted to replicate this result using continuous clinical measures. For this analysis, we used the SWAN inattentive and hyperactivity/impulsivity subscale scores in the POND and HBN datasets, which both had this measure available. We created a second autism+ADHD subgroup of autistic individuals who also met the clinical cut off of scoring 6 or higher on either the Inattentive or Hyperactive/Impulsive SWAN subscales (N=118 post QC), and repeated the same analysis described in the “Co-occurring autism and ADHD” methods section. We compared the effect size maps for the overall main effect to the original autism+ADHD analysis results by conducting spearman correlations between maps, and using spin tests to assess significance.

### Supplementary results

#### 1. Controlling for GMV, total SA and mean CT

Controlling for global brain measures (mean cortical thickness, total cortical grey matter volume, and total surface area) had a much greater impact on the results for ADHD than for autism. The results of the main effect of autism for cortical thickness and volume did not substantially change, though significant results were also observed for SA, which were not observed in previous analyses. These effects were similar to those seen in CT and CV, with increased SA in the superior temporal gyrus, as well as in the superior frontal gyrus and inferior temporal cortex. In ADHD, the results for CT disappeared entirely. Results for CV and SA were also dramatically altered. No significant regions of decrease were observed after controlling for total cortical GMV or SA, and instead increased CV and SA were observed in frontal regions. This highlights the fact that the autism effects are far more localised, with more global differences observed in ADHD.

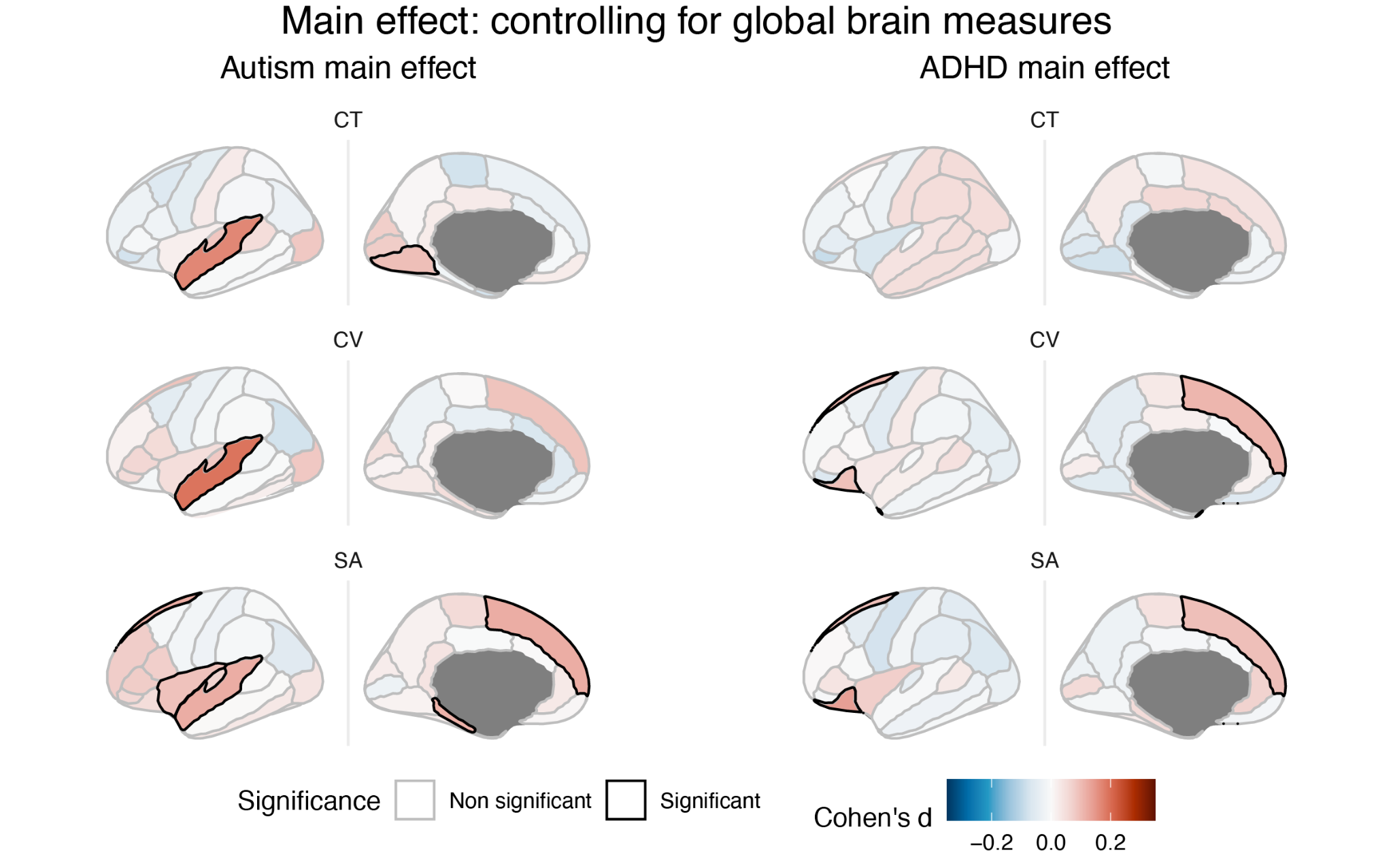

**Figure S1.1.** Main effect of autism and ADHD after controlling for global brain measures.

Interaction effects for autism were altered slightly, such that no regions remained significant after controlling for global measures. However, subthreshold effect size maps looked largely similar. ADHD again showed more differences for CV and SA, but still no regions reached significance. In the sex stratified analyses, male analyses were again very similar to those in the whole sample, with effects in autism having been less impacted than those in ADHD. Results for females were also slightly altered for both autism and ADHD, with only the temporal pole and occipital lobe regions reaching significance in ADHD (for CT and CV) after controlling for global measures, and for autism the post central gyrus and bank of the superior temporal gyrus became significant for CT (autism>TD).

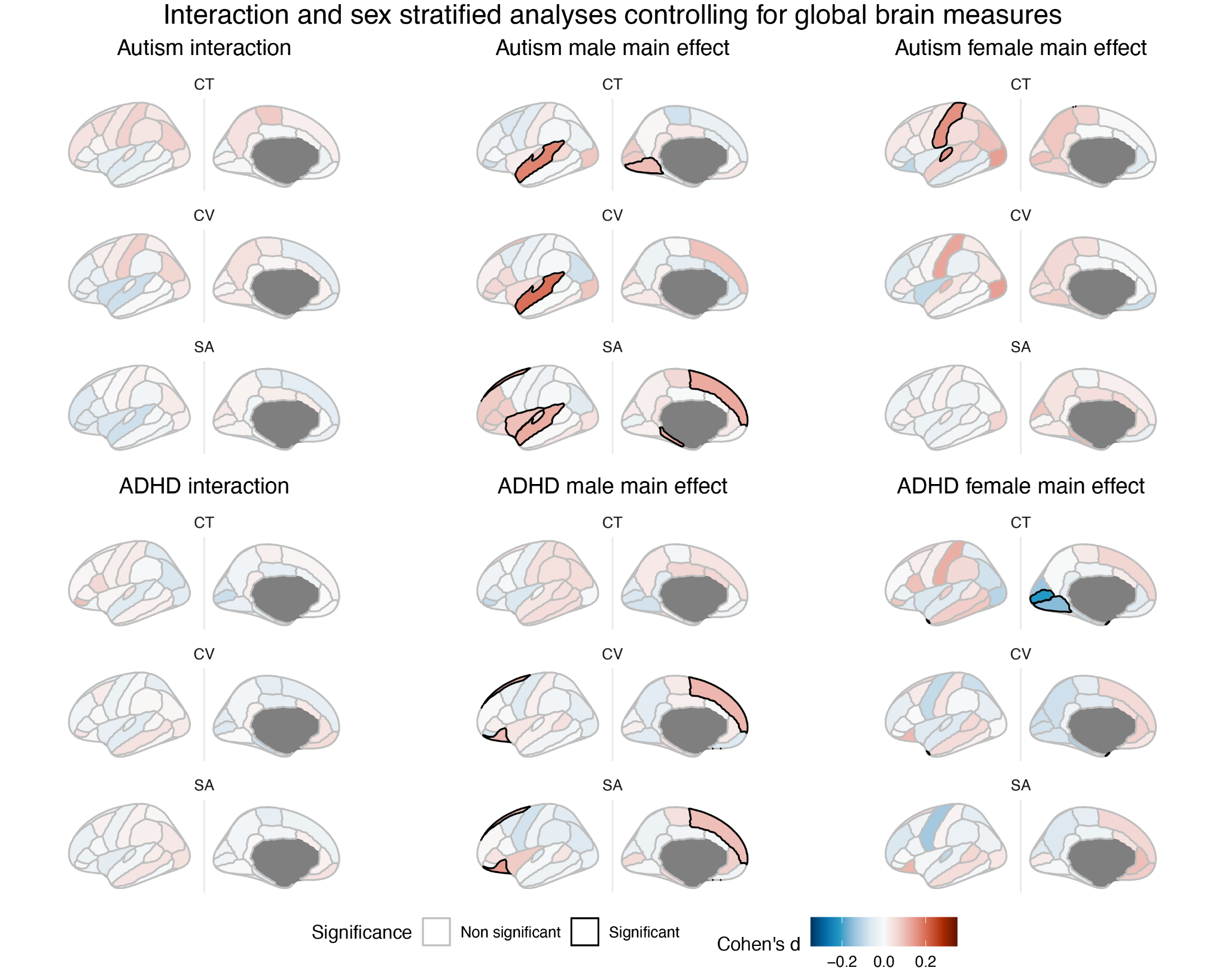

**Figure S1.2.** Interaction and sex stratified effects of autism and ADHD after controlling for global brain measures.

#### 2. Sensitivity analysis with varying QC methods

QC method and combination did not substantially impact results, in particular for main effects. There were some differences in which regions reached significance in each analysis; most notably, when thresholding by both FSQC and Euler number, there were no significant effects for the autism diagnosis by sex interaction, and when thresholding by Euler only, there was a significant interaction between sex and autism diagnosis for SA as well as CV. However, all subthreshold effect size maps looked almost identical across QC conditions (figure S2.1-2.3).

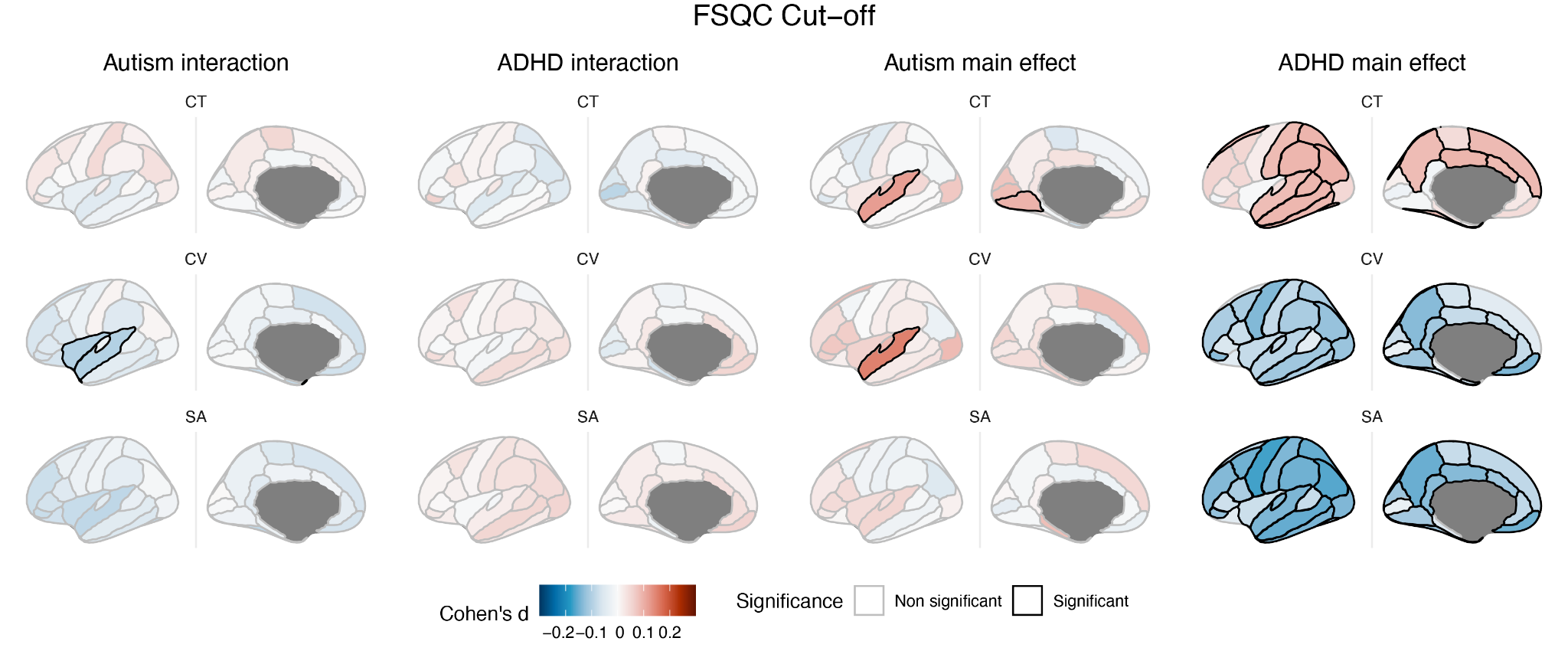

**Figure S2.1.** Sex-by-diagnosis interactions and main effects for autism and ADHD when thresholding by FSQC only. Results were extremely similar to the original analysis.

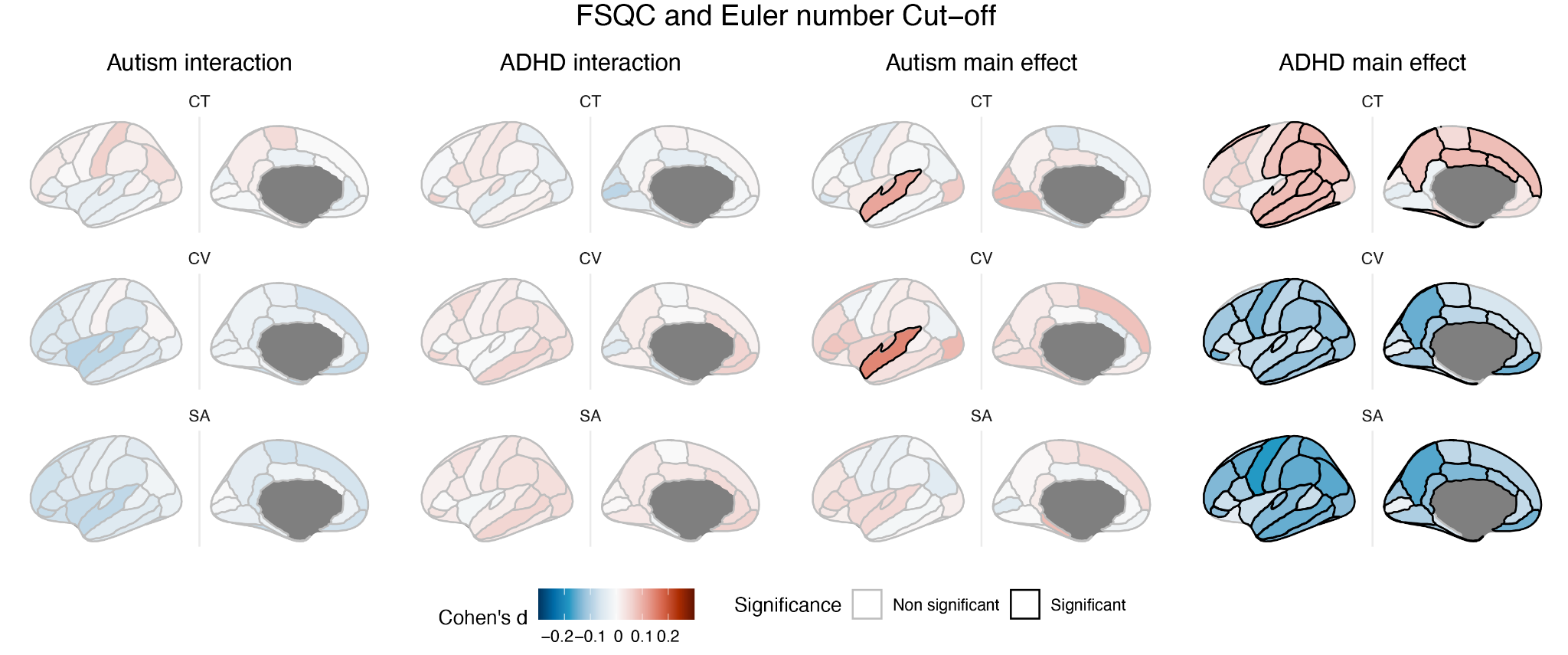

**Figure S2.2.** Sex by diagnosis interactions and main effects for autism and ADHD when thresholding by FSQC and Euler number. Results were extremely similar to the original analysis, however no regions reached significance in the interaction between autism diagnosis and sex.

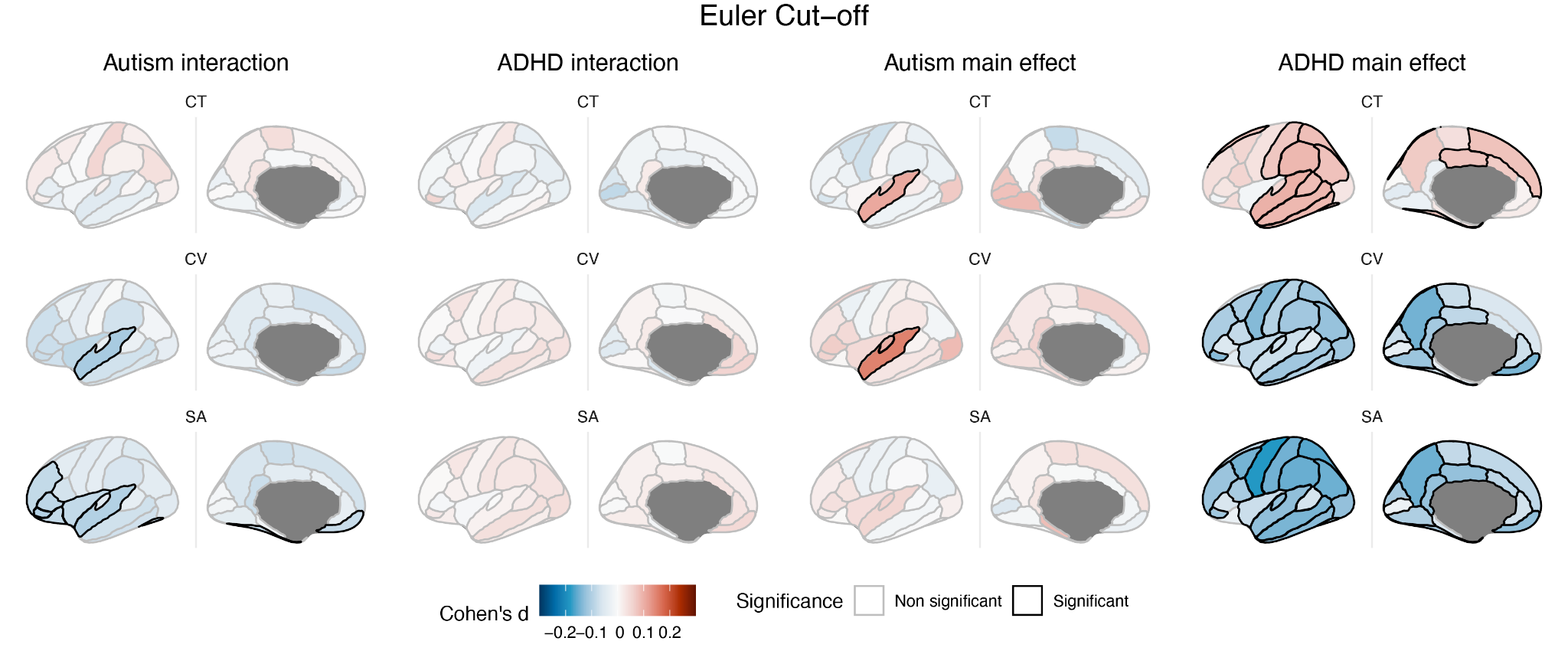

**Figure S2.3.** Sex by diagnosis interactions and main effects for autism and ADHD when thresholding by Euler number only, at 2 median absolute deviations. Results were extremely similar to the original analysis, however regions reached significance in the interaction between autism diagnosis and sex for surface area, which they did not in the other analyses.

#### 3. Spatial comparison of male and female diagnosis effect size maps

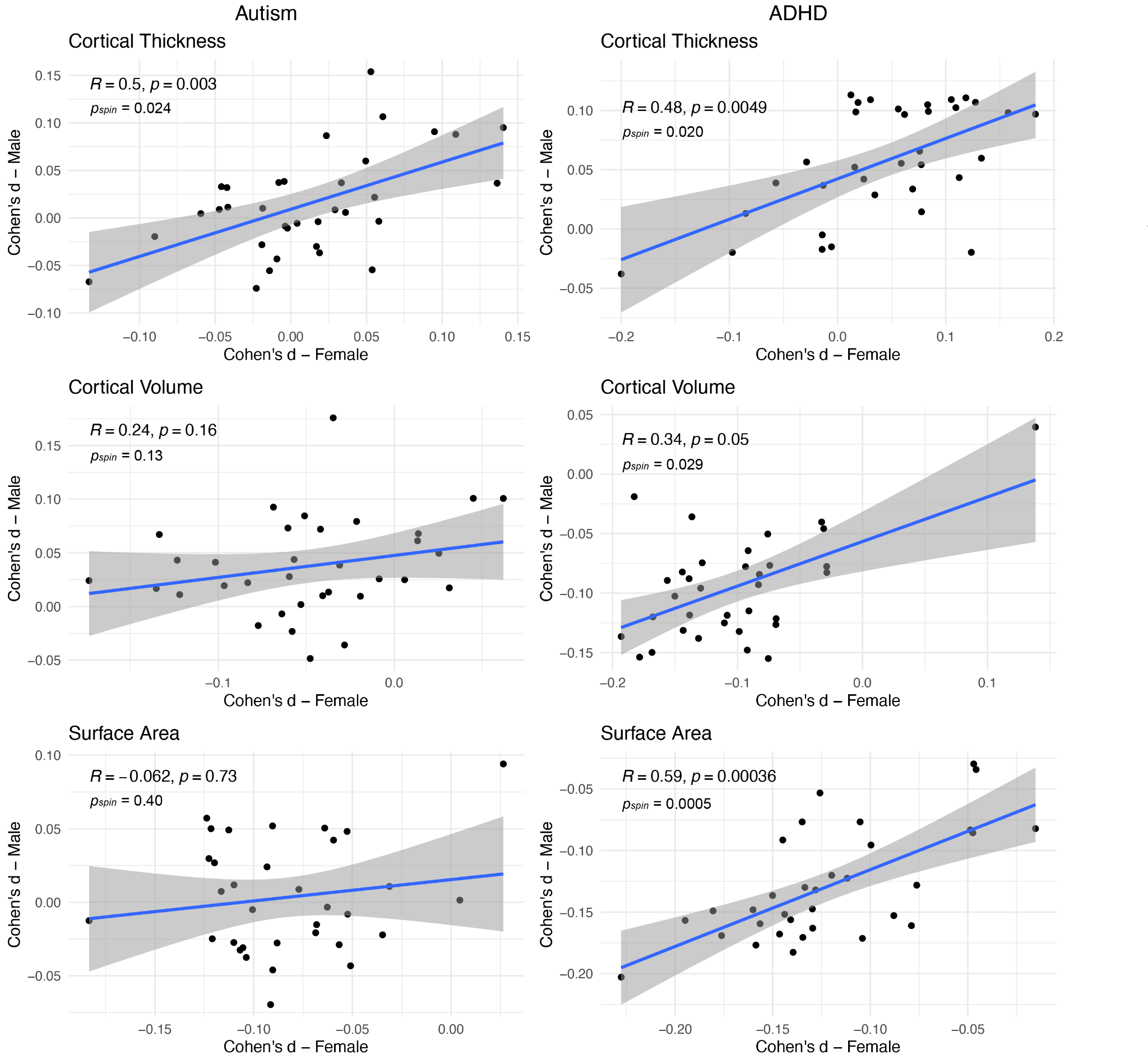

**Figure S3.1.** Correlations between male and female effect size maps for main effect of autism (left) and ADHD (right) diagnosis, for cortical thickness (top), volume (middle) and surface area (bottom). Regional effect size maps for ADHD diagnosis were extremely similar between males and females, and all cortical phenotypes were significantly correlated even based on the more conservative spin permutation test. The same is true of autism diagnosis effect size maps only for cortical thickness; surface area and cortical volume maps differed between males and females and were not significantly correlated.

#### 4. Sensitivity analysis on subsample of matched male data

Results examining the main effects of diagnosis in the full sample were largely similar in terms of spatial mapping of effect sizes, but with some differences in which regions met significance. For autism, the superior temporal gyrus remained significant only for CV, but subthreshold effect size maps were largely the same. For ADHD, more regions reached significance for CT (ADHD>TD), though for CV almost no regions remained significant; significance and effect sizes were slightly attenuated for SA as well, though not to the same extent (Figure S4.1). Correlations between effect size maps of the original sample and the new sex-matched sample revealed significant, positive correlations of rho=0.57-0.78 for autism and rho=0.73-0.89 for ADHD (all p_spin_<0.001).

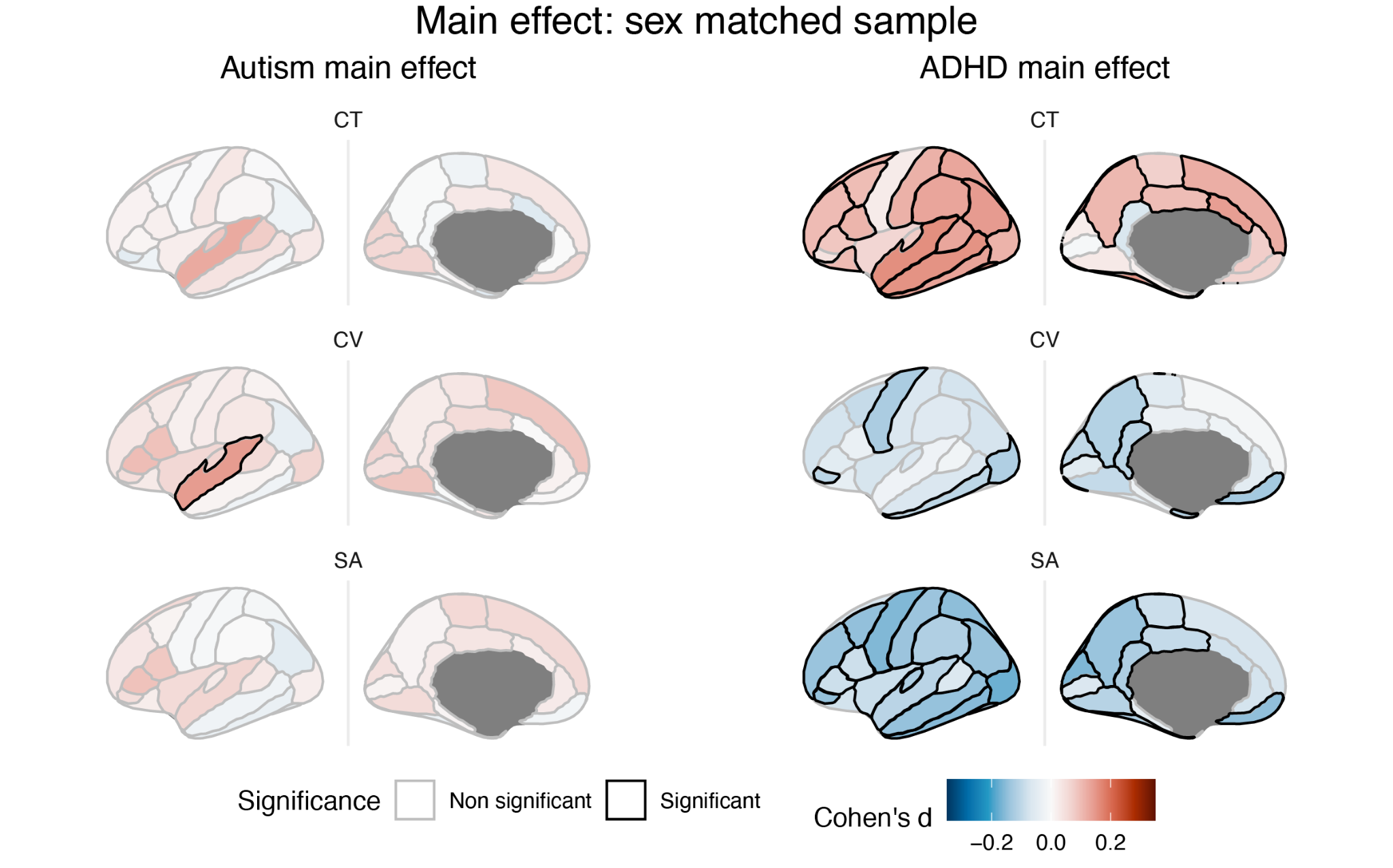

**Figure S4.1.** Main effect of autism (left) and ADHD (right) diagnosis in a sex matched subsample with equal numbers of males and females. Results are largely unchanged from the original analyses, though with fewer regions reaching significance.

No regions reached significance for the interaction between either diagnosis and sex. Again, subthreshold effect size maps for an interaction with sex looked similar and correlations with maps from the full sample were rho=0.72-0.87 for ADHD and rho=0.78-0.87 for autism (p_spin_<0.001). In the male only analysis, results were essentially the same as for the whole group, though with fewer still significant regions for SA in ADHD, and no significant regions for CV. Correlations with previous maps for the male analysis were rho=0.73-0.91 for ADHD and rho=0.60-0.81 for autism (all p_spin_<0.001). Female results are unchanged as the sample remained the same, but they are shown below for the sake of comparison (Figure S4.2).

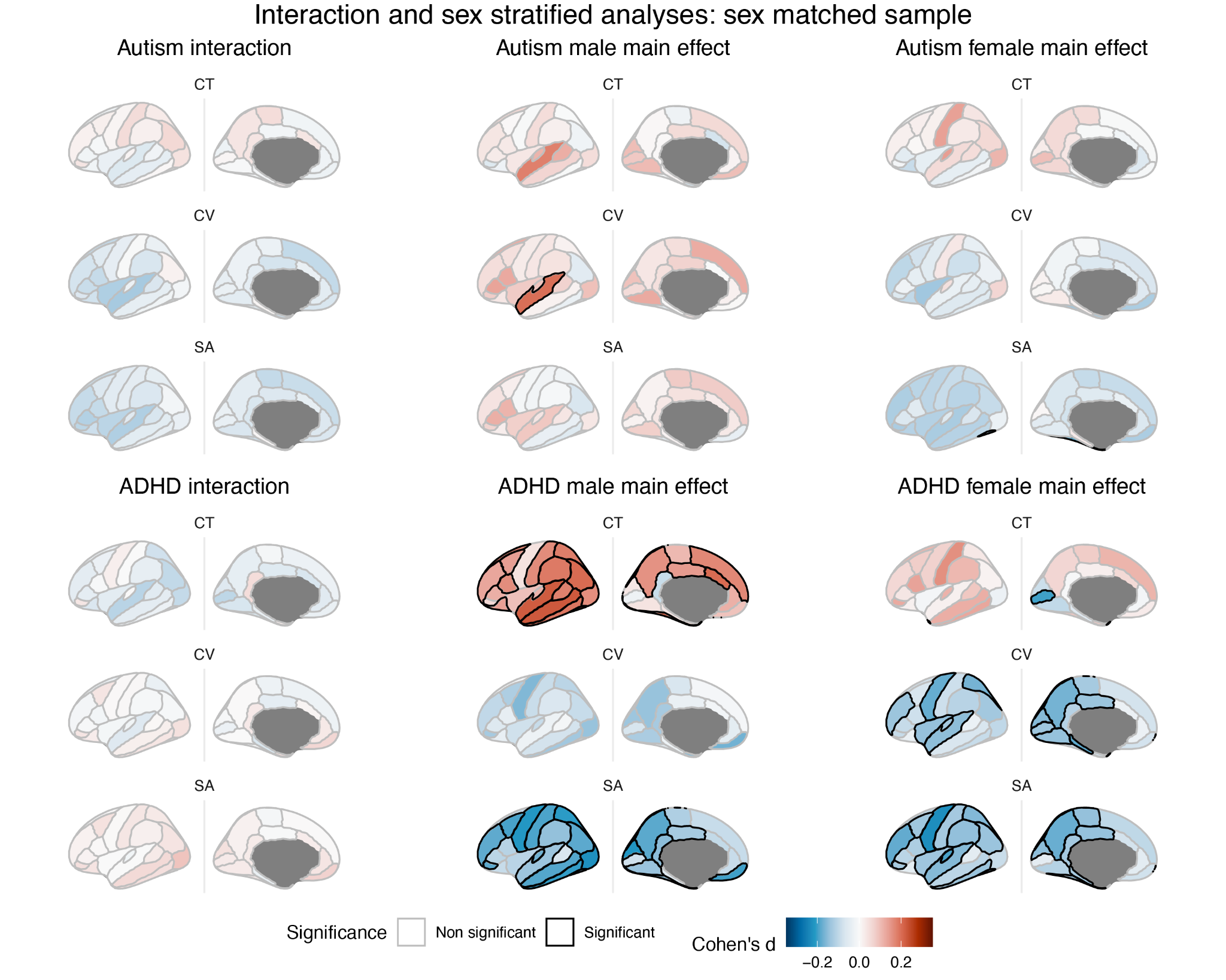

**Figure S4.2.** Interaction with sex, and sex-stratified analyses for autism (top) and ADHD (bottom) in a sex matched subsample with equal numbers of males and females. Results are largely unchanged from the original analyses, though with fewer regions reaching significance.

#### 5. Multimodality of data

Hartigan dip test analyses revealed that most regions did not reach significance for multimodality, even if they were deemed to have multiple peaks. Autism had the most regions with a multimodal distribution, with the majority being significant for all three cortical phenotypes. In ADHD, only a few regions for CT were significant, and for controls there was only one or two regions for each phenotype.

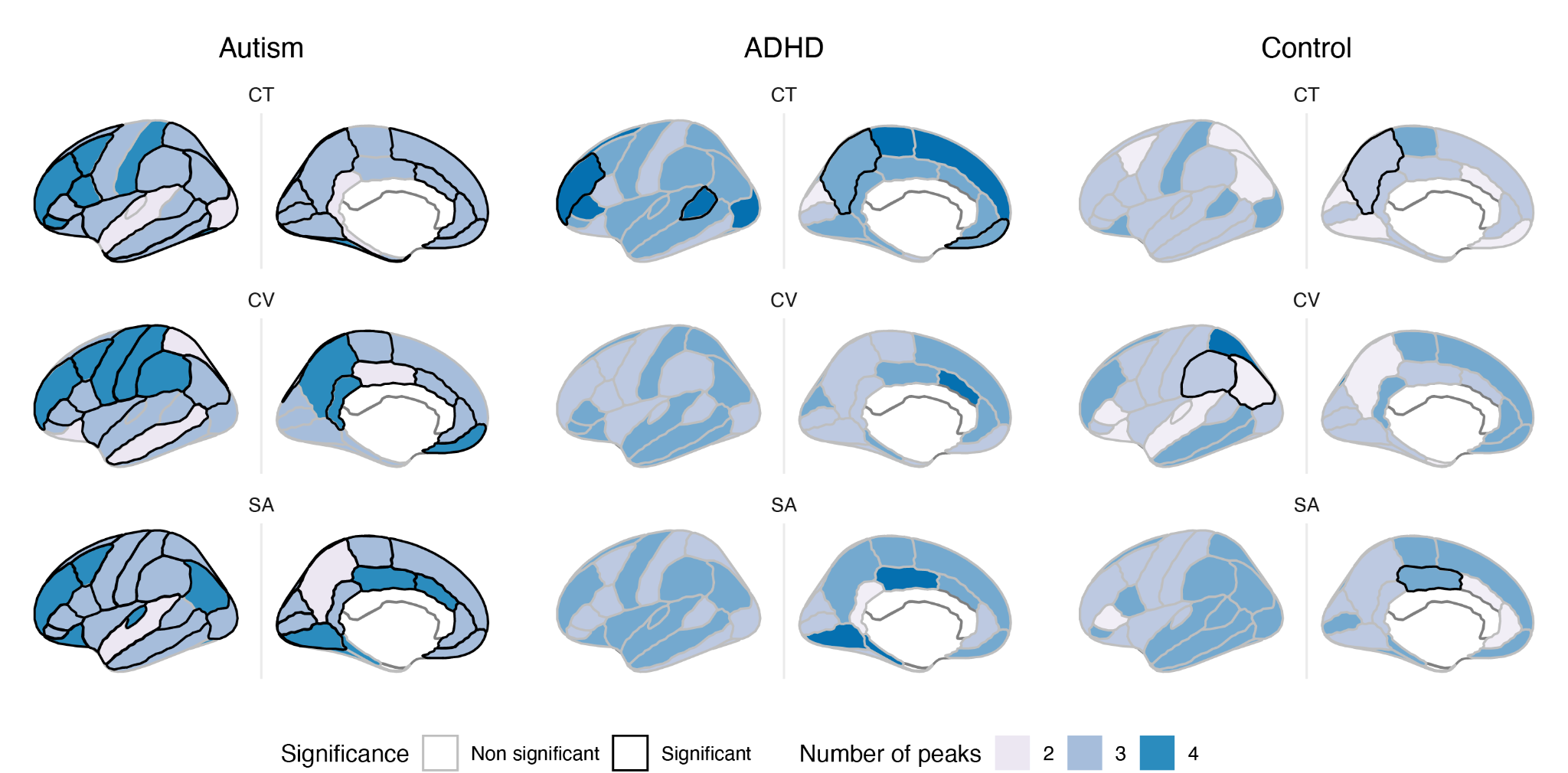

**Fig S5.1.** Significance of multimodality and number of peaks in the probability density function across cortical regions and phenotypes for each diagnosis. Colour represents the number of peaks in the distribution (darker colours represent higher numbers). Significance of multimodality is signified by the black outline.

#### 6. Dimensional analyses of autistic traits

##### 6.1 Distribution of scores by group

We examined the means and distributions of each measures’ scores, plotted separately by diagnostic group. There was a significant difference between each pairwise combination of groups, for every measure. ADOS was primarily available for autistic participants, with a limited number of control participants and participants with ADHD available. For all measures, including the SWAN which is a measure of ADHD behaviours, individuals with ADHD displayed an intermediate phenotype in which they showed elevated traits on both scales relative to controls, but less than the autistic group.

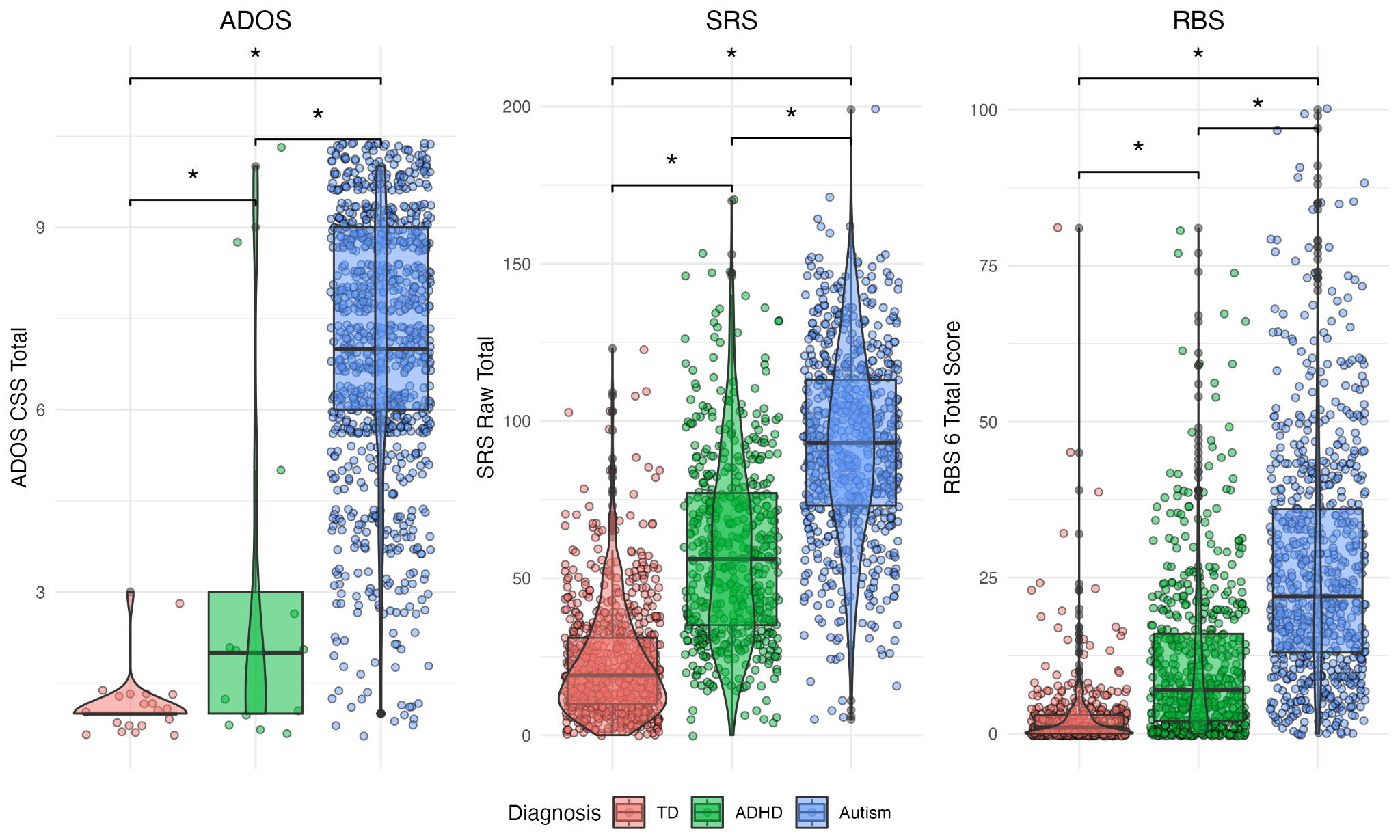
**Figure S6.1.1.** Score distributions for ADOS, SRS (raw total) and RBS, shown by diagnostic group. For all measures, individuals with ADHD display an intermediate phenotype between controls and autistic participants.

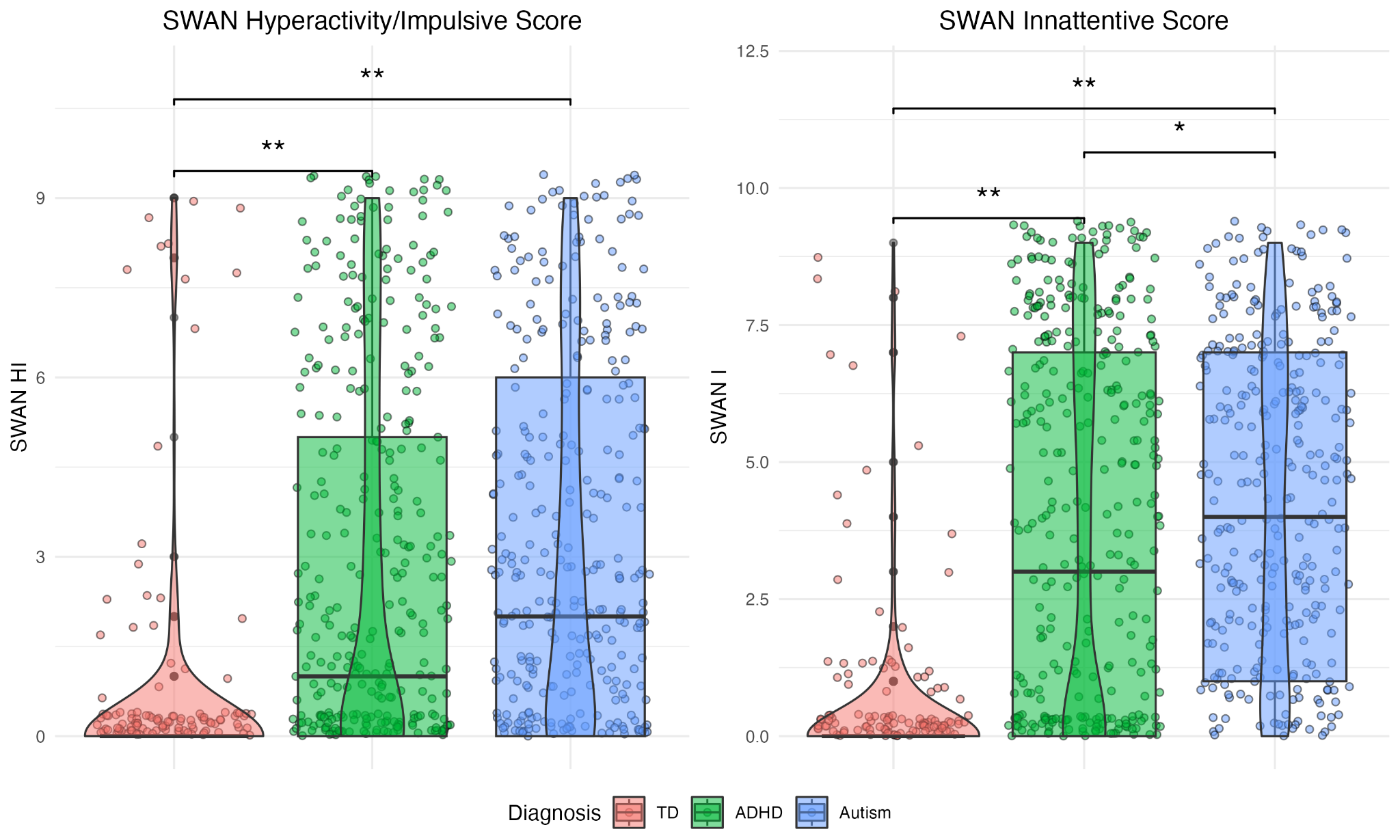

**Figure S6.1.2.** Score distributions for SWAN Hyperactivity/Impulsivity (HI) and Inattentive (I) subscales, shown by diagnostic group. Even for these measures, autistic individuals had higher scores than the ADHD group (though the difference was only significant for the I scale), and both groups had significantly higher scores than the control group.

##### 6.2 Associations between autistic/ADHD traits and cortical measures

###### Associations with autistic traits

The ADHD group displayed an intermediate phenotype between the control and autistic groups for all scores (Figure S6.1.1-6.1.2). Males had significantly higher levels of autistic traits as per the ADOS CSS (*d* = 0.29), SRS raw (*d* = 0.21) and RBS-R scores (*d* = 0.25), and the SWAN HI (*d* = 0.31) and SWAN I (*d* = 0.30; all *P* < 0.0001).

There were no significant cortical associations with ADOS CSS, but RBS-R showed weak but significant negative correlations with cortical surface area and volume (partial r = -0.06 - -0.12), and SRS with surface area only (partial r = -0.06 - -0.10; Figure S6.2.1). No measure-by-sex interactions were significant. Associations with cortical measures were similar for SRS T-scores, though fewer regions reached significance for SA (Figure S6.5).

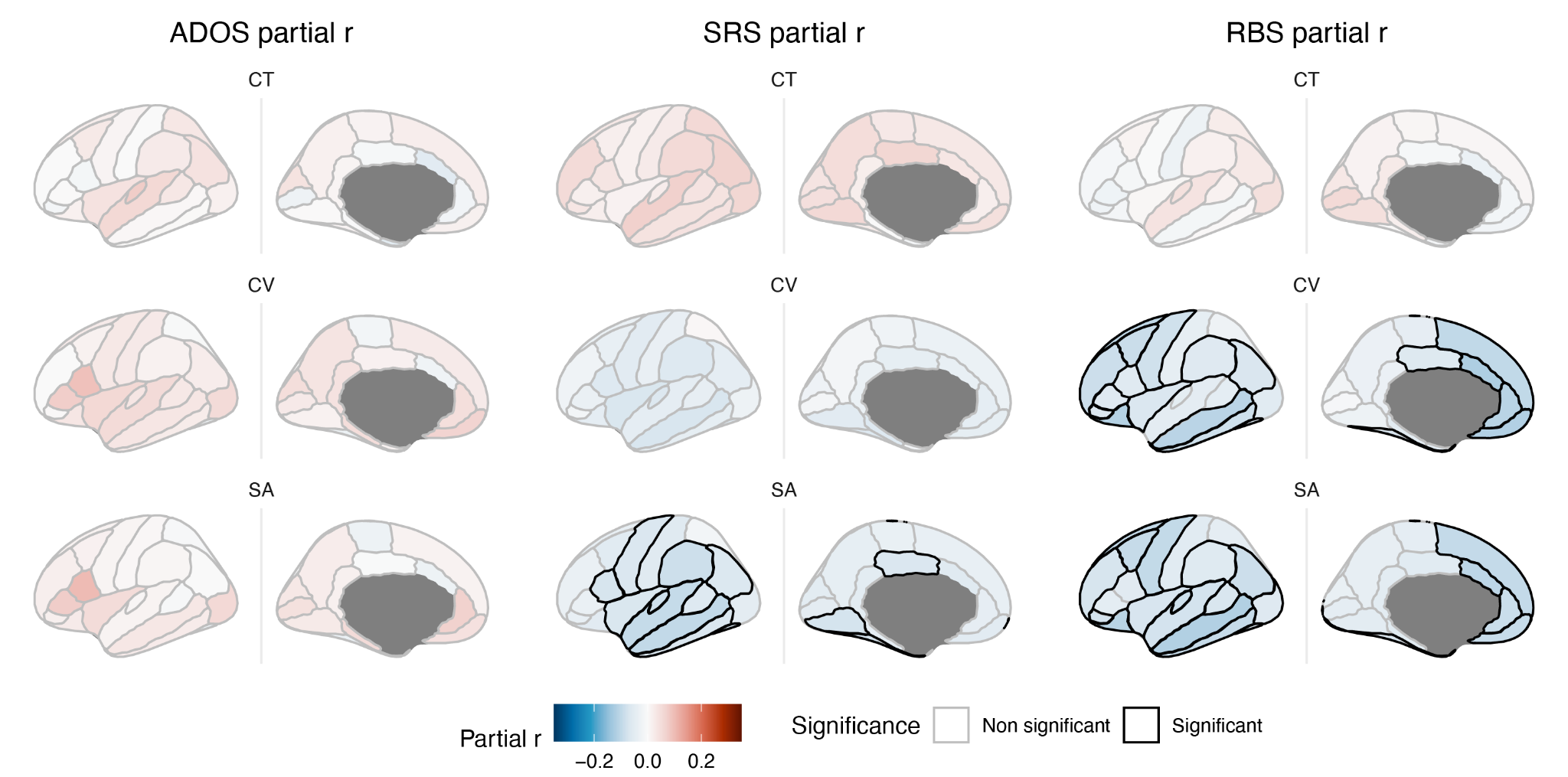
**Figure S6.2.1.** Association between autistic traits and cortical measures. Maps show partial correlation for each measure (ADOS, SRS, RBS-R), with significant regions (passing 5% FDR) outlined in black. Red represents positive correlations (increasing cortical measure with increasing levels of autistic traits), and blue represents negative effect sizes (decreasing cortical measure with increasing levels of autistic traits).

###### Associations with ADHD traits

As with the autistic trait measures, significant but weak associations between SWAN subscale scores and cortical measures were observed. A significant positive association was observed between both the SWAN HI and I scales and CT in the STG and transverse temporal gyrus (partial r = 0.10-0.14), as well as a significant negative association in the frontal pole for CV (partial r = -0.12- -0.14). For the SWAN I scale, a significant positive association was also observed for CT in the lingual gyrus (partial r =0.12), as well as significant negative associations with SA in frontal, temporal and occipital regions (partial r = -0.10- 0 0.13). No measure-by-sex interactions were significant.

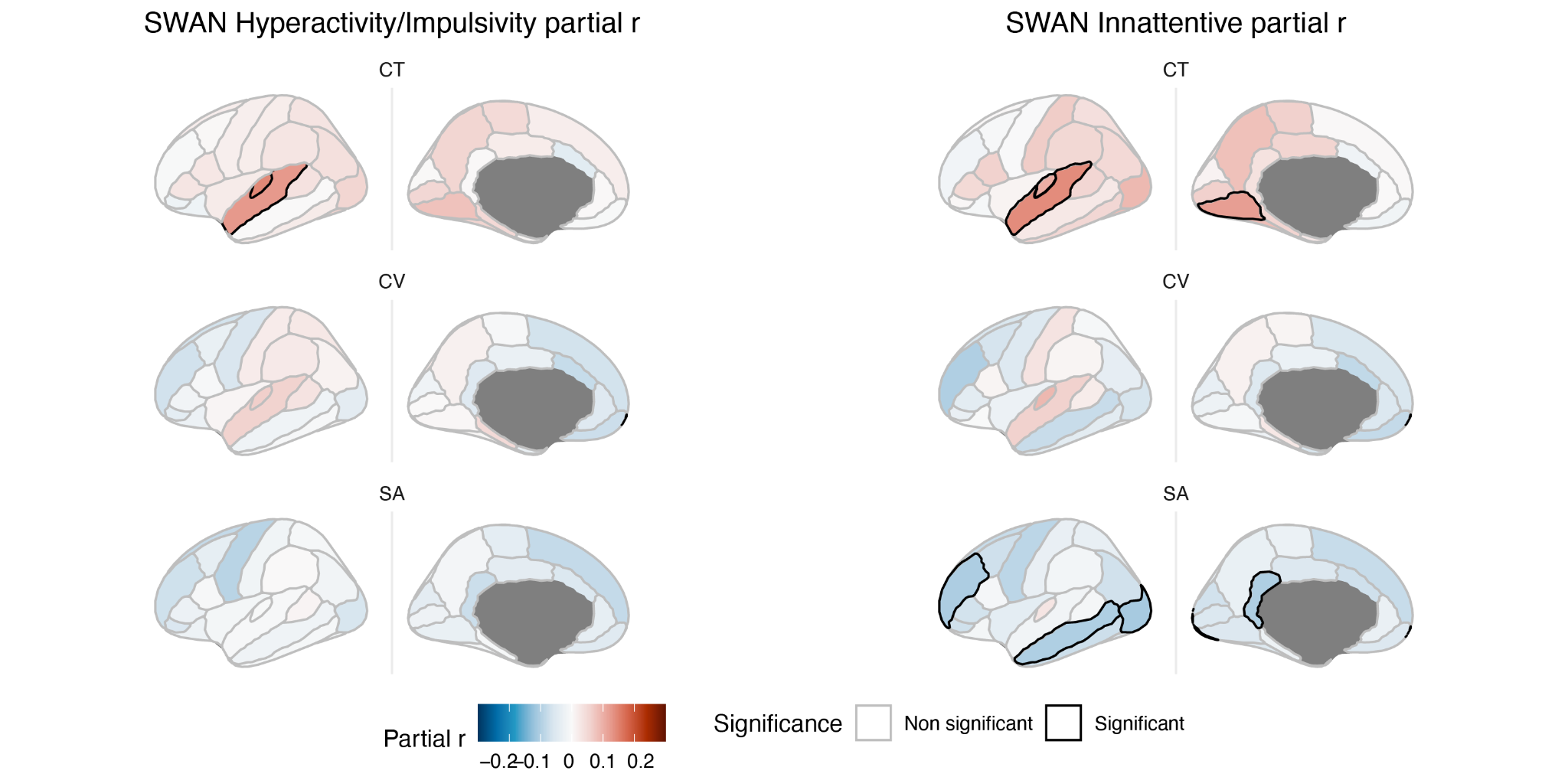

##### **Figure S6.2.2.** Association between ADHD traits and cortical measures. Maps show partial correlation for SWAN Hyperactivity/Impulsivity and Inattentive scales, with significant regions (passing 5% FDR) outlined in black. Red represents positive correlations (increasing cortical measure with increasing levels of ADHD traits), and blue represents negative effect sizes (decreasing cortical measure with increasing levels of ADHD traits).

##### 6.3 Associations between autistic/ADHD traits and cortical measures controlling for global brain measures

No associations for autistic traits (ADOS, SRS or RBS) remained significant after controlling for global cortical measures (total GMV, mean CT or total SA). However, associations with ADHD traits (SWAN subscales) were less impacted. Associations with SA were no longer significant after controlling for total SA; however, CT results were unchanged, and additional regions of significance were observed for CV, with the STG and transverse temporal gyrus again also a significant positive relationship, and a negative relationship in the prefrontal cortex.

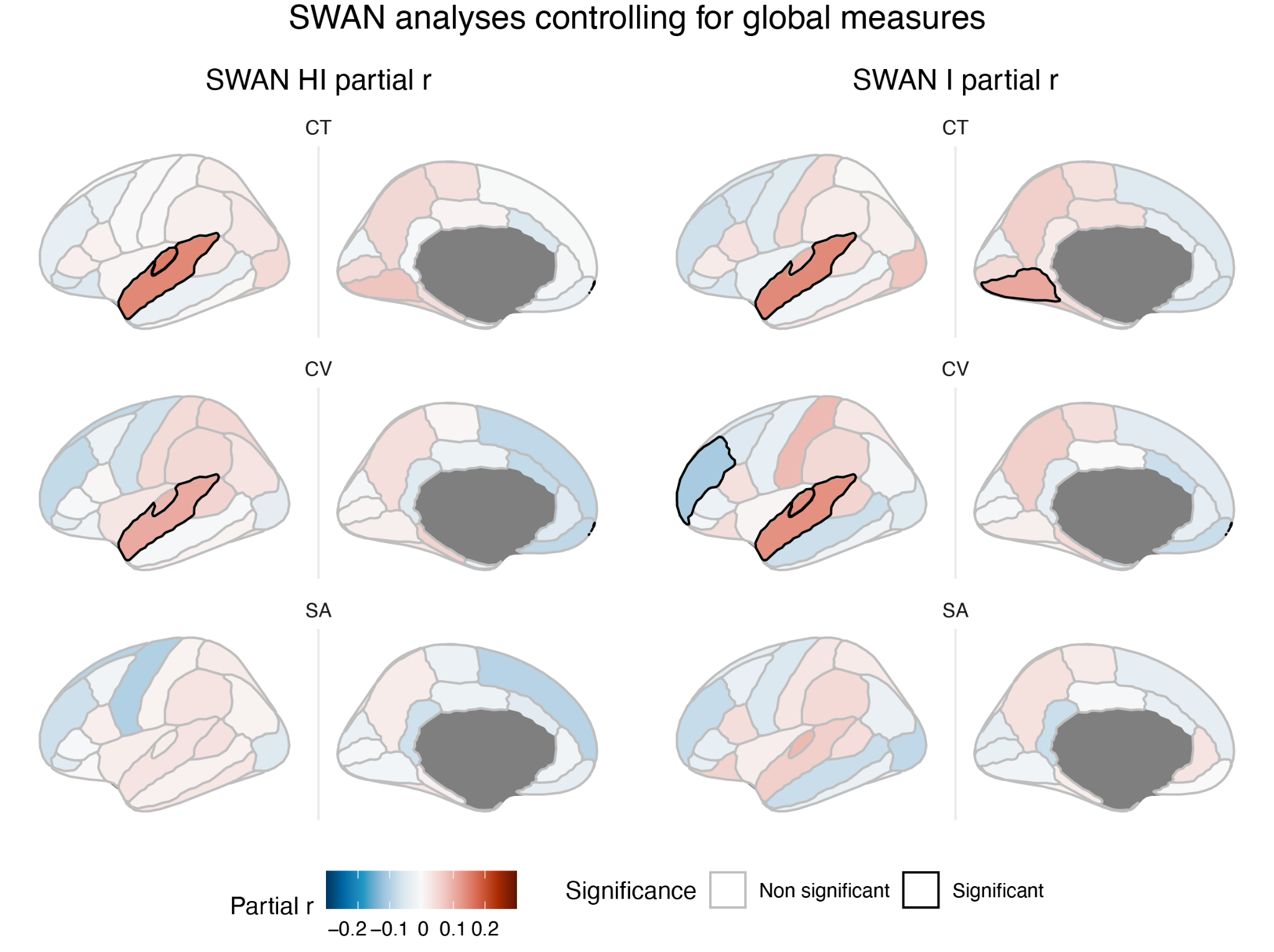

**Figure S6.3.1.** Associations between ADHD traits and regional cortical measures, controlling for global cortical measures.

##### 6.4 Relationship by group in significant regions

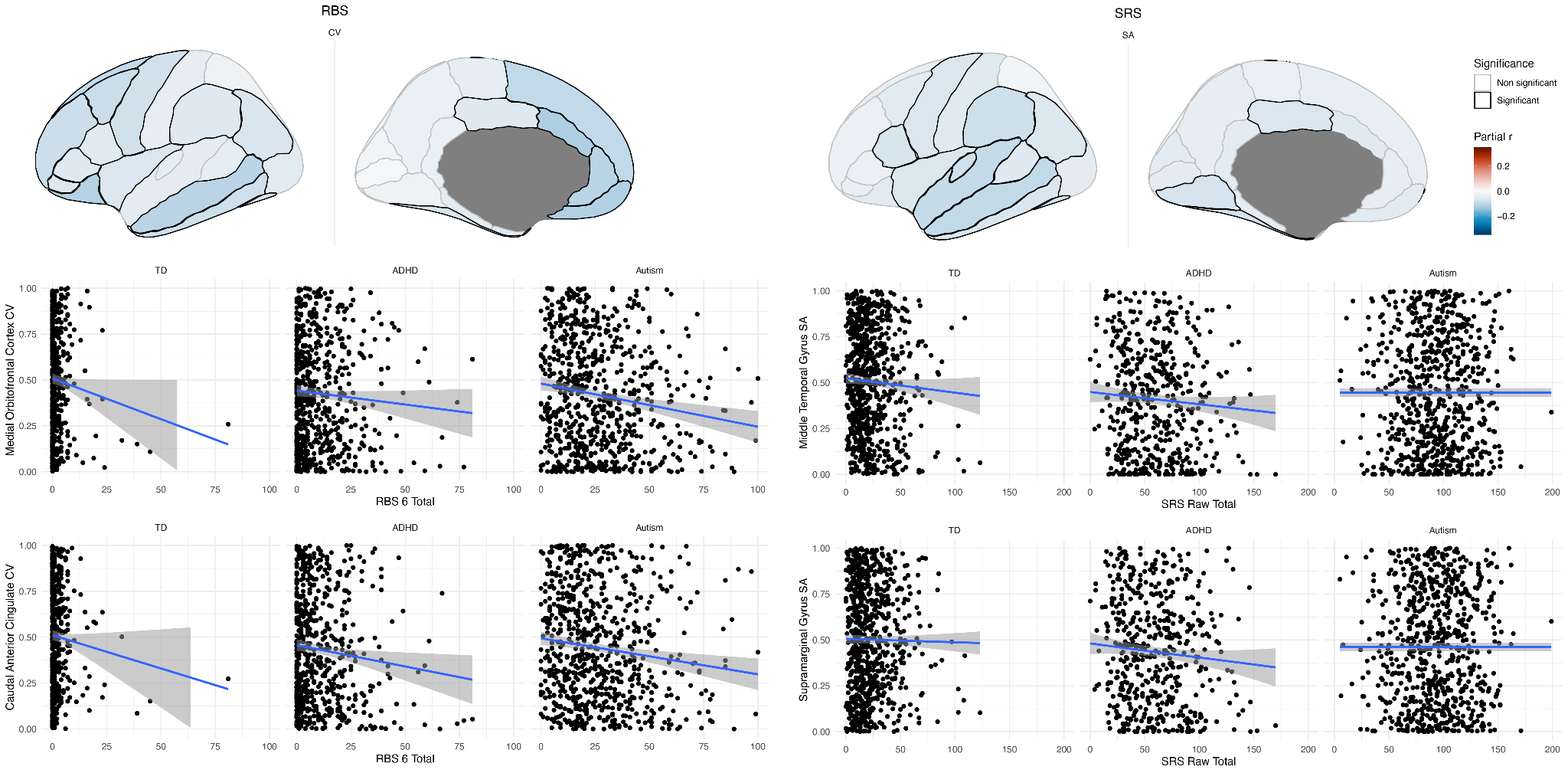

**Figure S6.4.1.** Maps showing associations between CV and RBS total (left) and SA and SRS total (right). Scatterplots show the relationship between significant cortical regions and total score, separately by diagnostic group.

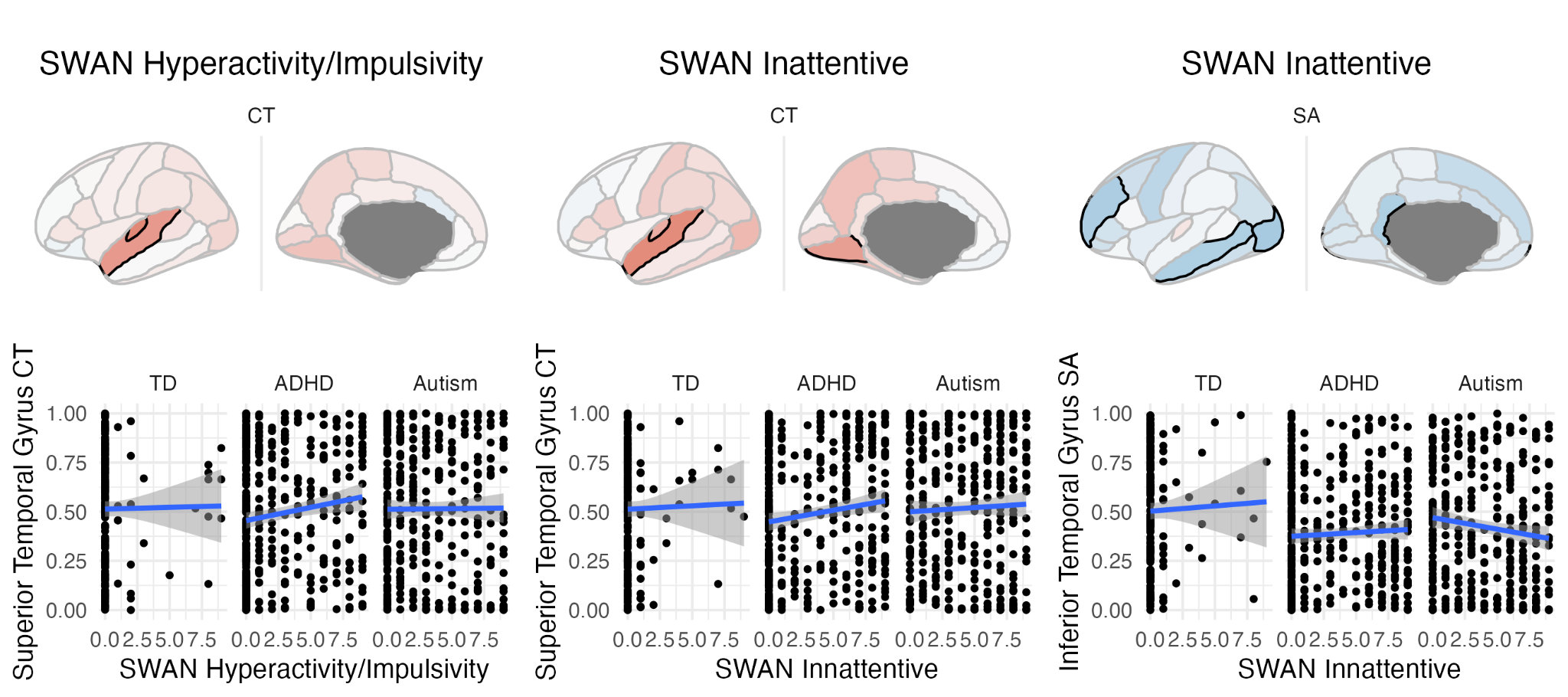

**Figure S6.4.2.** Maps showing association between CT (left two) SWAN H/I and I scales, and SA and SWAN I scale (top) and scatterplots showing the relationship between significant cortical regions and score, separately by diagnostic group.

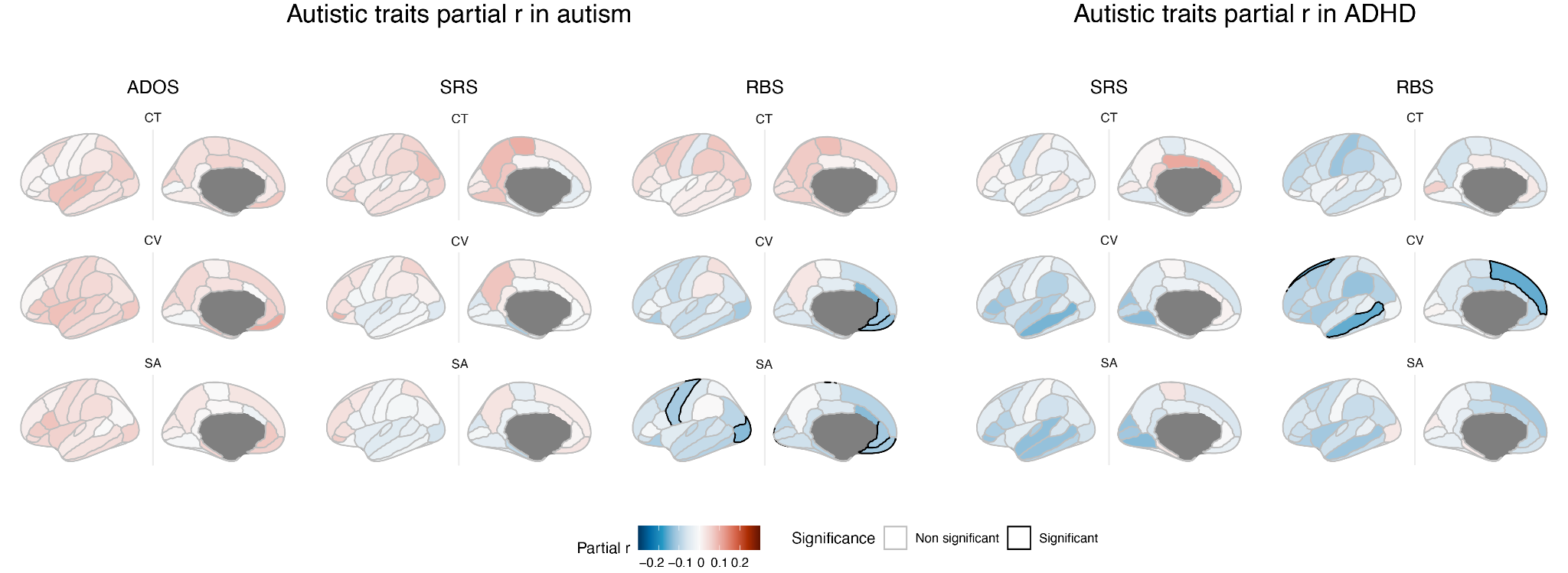

**Figure S6.4.3.** Association between autistic traits and cortical measures in autism (right) and ADHD (left) separately. Maps show partial correlation for each measure (ADOS, SRS, RBS-R), with significant regions (passing 5% FDR) outlined in black. Note that there were not enough individuals with ADHD who had ADOS scores to complete this analysis.

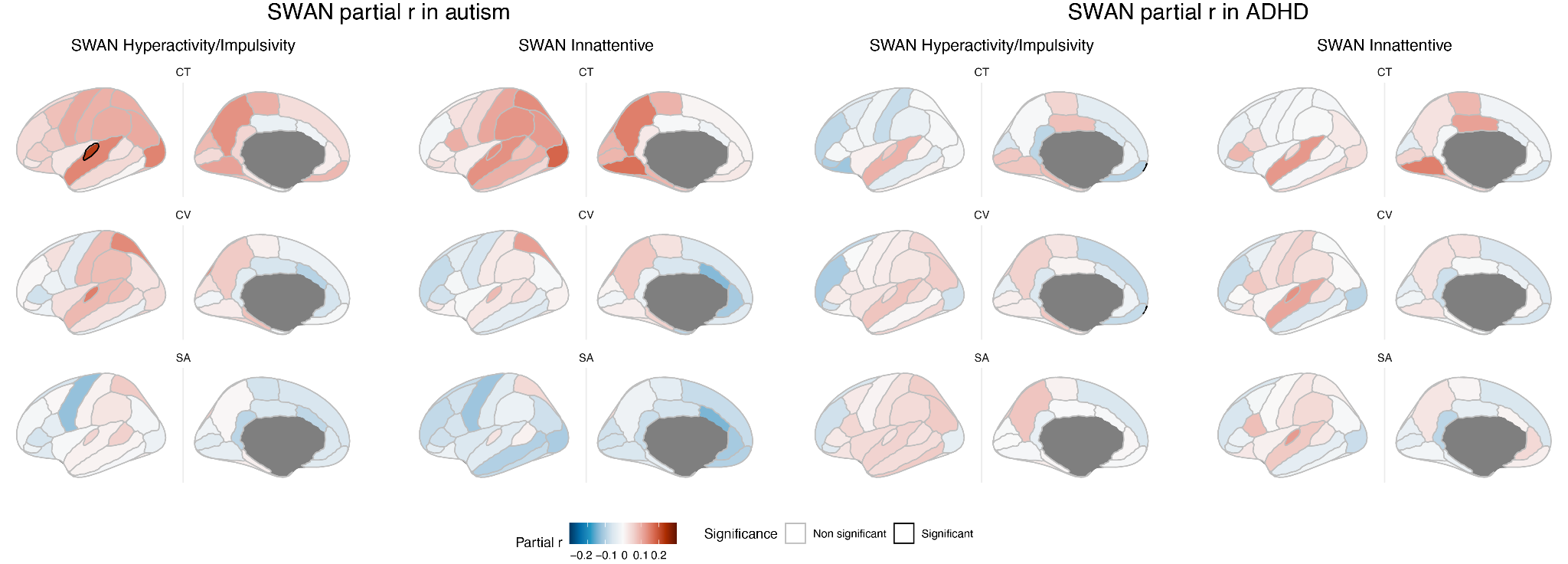

**Figure S6.4.4.** Association between ADHD traits and cortical measures in autism (right) and ADHD (left) separately. Maps show partial correlation for each measure (ADOS, SRS, RBS-R), with significant regions (passing 5% FDR) outlined in black.

##### 6.5 Sensitivity analysis using SRS T-scores

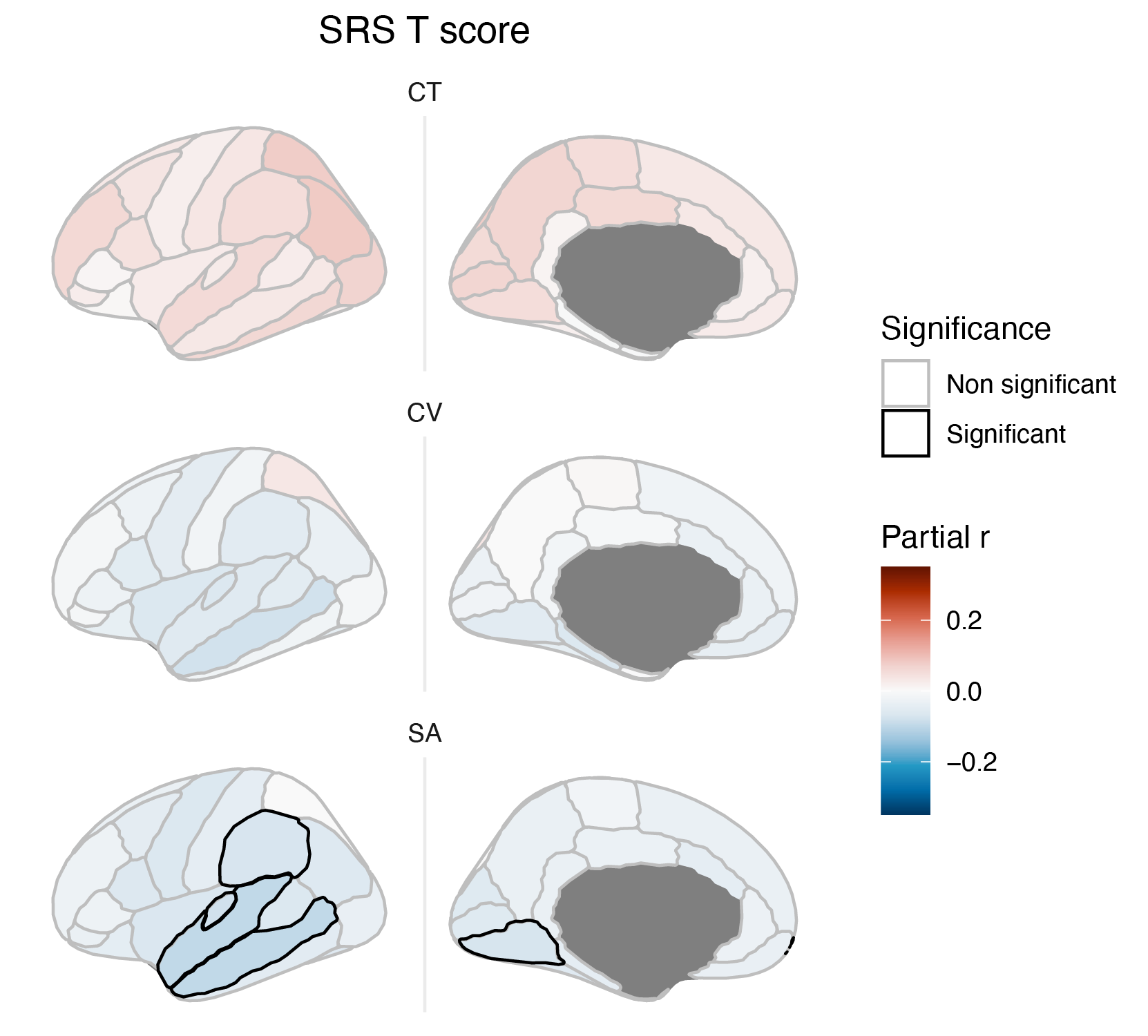

**Figure S6.5.** Association between SRS T-score and cortical measures. As in the raw score analyses, only SA showed significant associations, but here fewer regions reached significance. The SRS T-score, which is scaled by sex, was the only measure that was not significantly different between males and females*.*

#### 7. Age interaction on subsample of age-matched controls

Results largely held consistent with the whole sample analysis for CT, with more regions reaching significance for the ADHD diagnosis by age interaction when restricting the TD group to the same age range. However, no regions remained significant for CV. Positive interactions were observed in frontal, parietal and some occipital regions, reflecting increased CT with age in the ADHD group but decreases or no effect in the TD group. Negative interactions were observed in the insula and bank of the superior temporal sulcus, reflecting a more negative relationship between age and CT in ADHD than the control group (Figure S7.1).

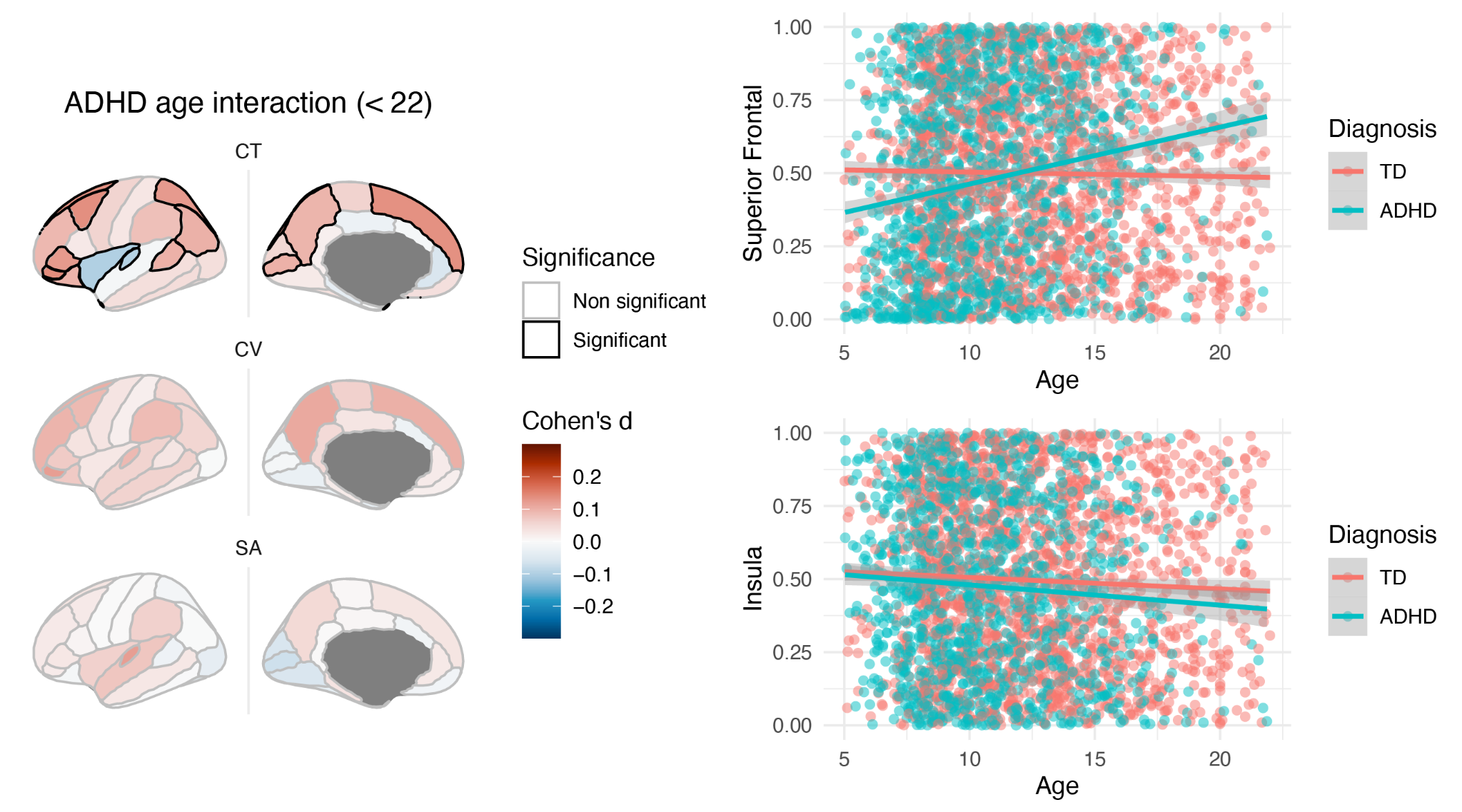

#### **Figure S7.1.** Significant interaction between age and ADHD diagnosis in cortical thickness, with age range in controls limited to the same age range as the ADHD sample.

#### 8. Autism+ADHD comparison and replication analysis

##### 8.1 Correlation and overlap of brain maps between diagnostic groups

Perhaps unsurprisingly, the autism+ADHD group overlapped more with the autism and ADHD alone groups alone than did the autism and ADHD group with each other. Almost all correlations between the autism+ADHD group and other groups, for each phenotype, were significant (p<0.01). Only the correlation between ADHD and autism+ADHD for CT did not survive the more stringent spin test (p*_spin_* = 0.05). Only SA effect sizes were significantly correlated between autism and ADHD (Figure S8.1).

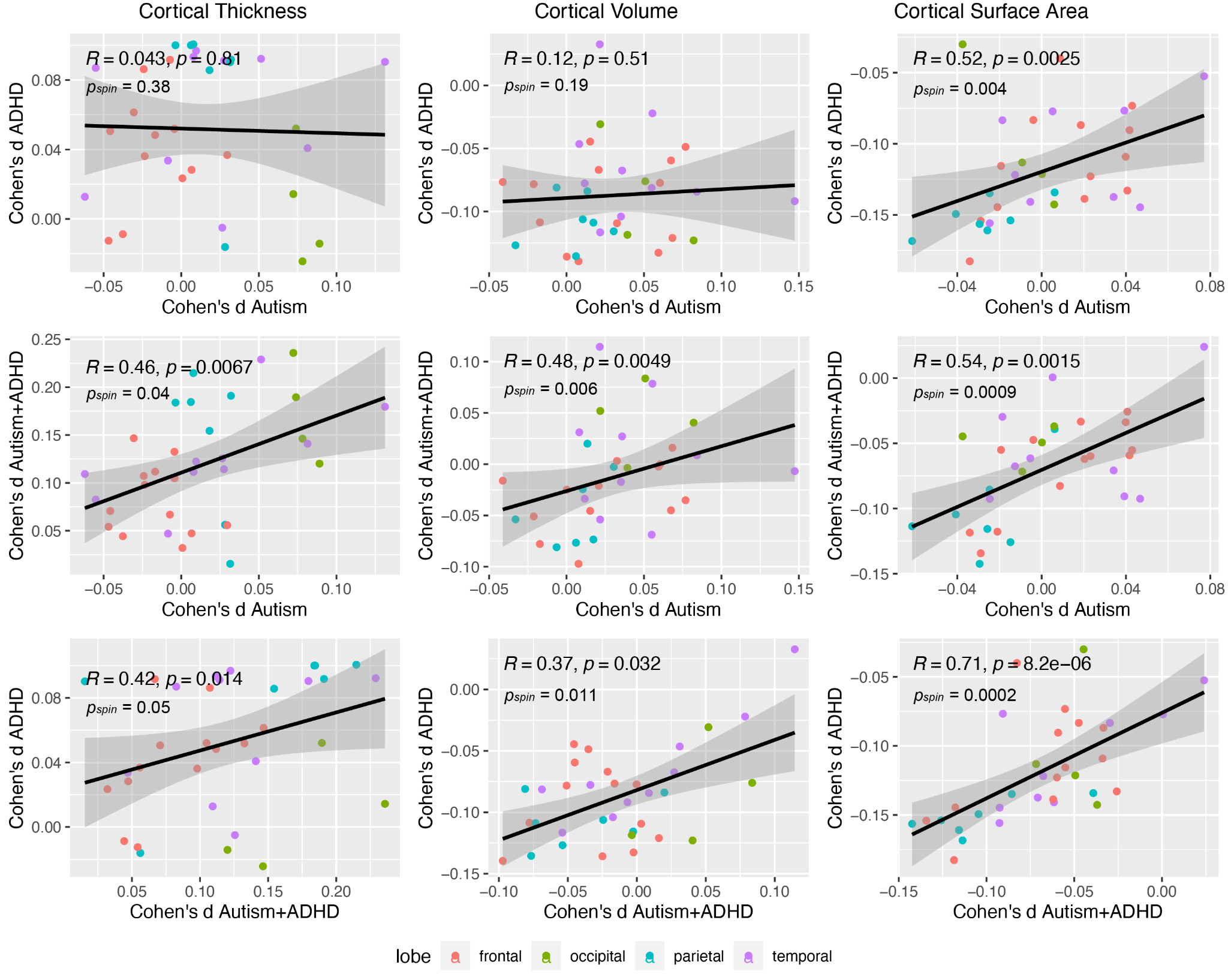

**Fig S8.1.** Pairwise correlations of Cohen’s d effect sizes between pairwise combinations of autism, ADHD, and autism+ADHD groups, across regions of the brain, coloured by lobe. Significance was tested using the more conservative spin test method.

##### 8.2 Autism+ADHD analysis controlling for global measures

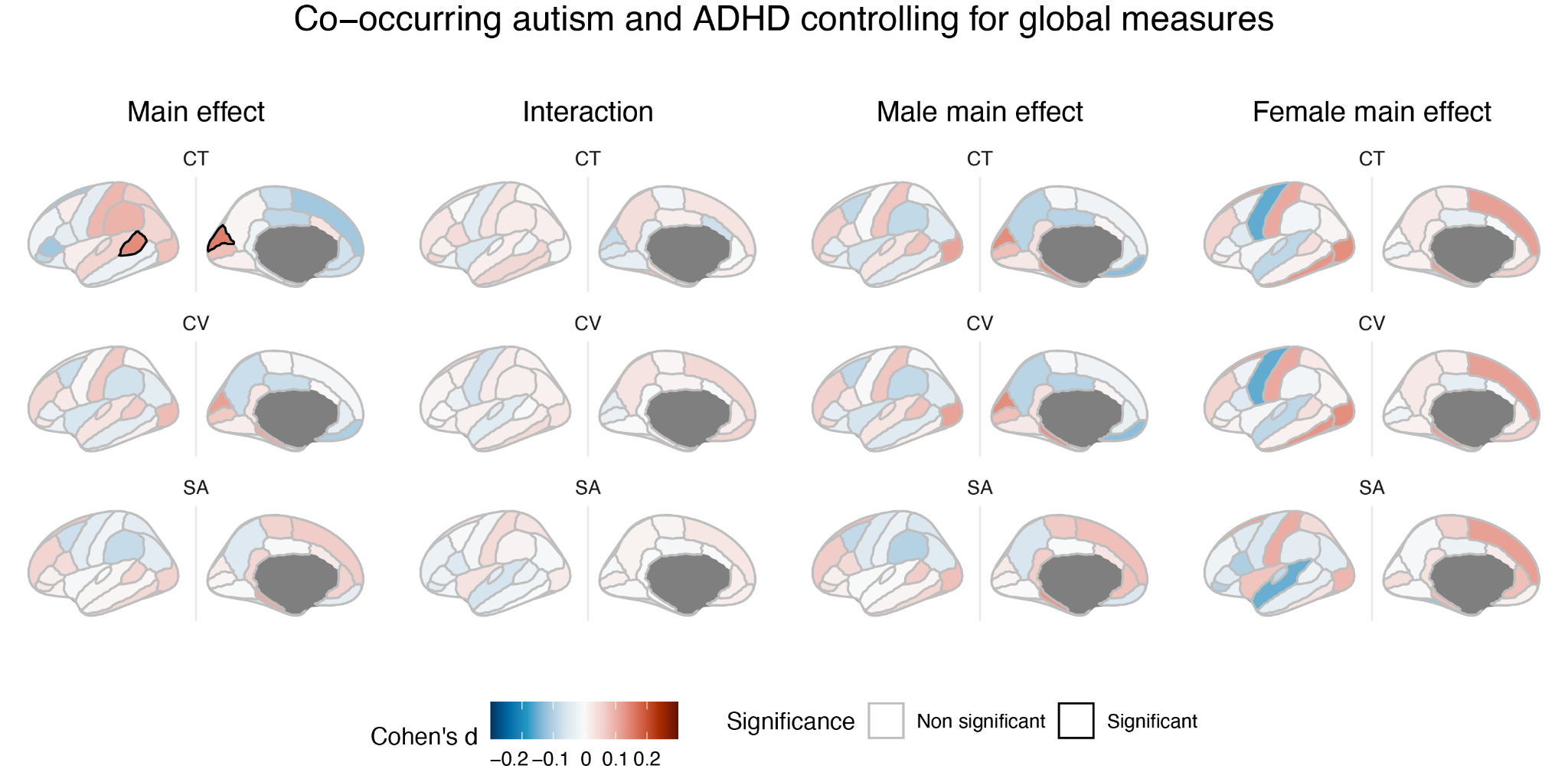

**Figure S8.2.** Cortical alterations (relative to controls) in individuals with co-occurring autism and ADHD controlling for global measures. Main effects of diagnosis relative to controls; interaction with sex; main effects in males, and main effects in females. All maps show Cohen’s *d* effect sizes, with significant regions (passing 5% FDR) outlined in black. Red represents positive effect sizes (autism+ADHD > controls), and blue represents negative effect sizes (autism+ADHD < controls). Only the cuneus and bank of superior temporal sulcus for CT remained significantly different between the autism+ADHD group and controls.

##### 8.3 Replication using SWAN subscale cut-off

The replication analysis based on the SWAN cut-off yielded similar results. As in the main analysis, there were no significant differences in CV centiles, and the autism+ADHD group displayed increases in CT centiles, and decreases in SA centiles in primarily frontal and parietal regions (Supplementary Figure S8.2). Slightly fewer regions reached significance in this analysis, but this may be due to a lack of power, as the number of autism+ADHD participants in this analysis was substantially less (N=118), and effect size maps looked similar. For the overall main effect across phenotypes, effect size maps were correlated at rho = 0.71. Maps were significantly correlated for CT (rho = 0.61; p*_spin_* = 0.012) and CV (rho = 0.35; p*_spin_* = 0.016), but not SA (rho = 0.21; p*_spin_* = 0.2).

The greatest difference between the two analyses is in the female main effect maps, for CV and SA. It should be noted however, that in the SWAN cut off analysis, the female and male effect size maps are more similar, with many overlapping regions in SA both showing significant decreases. It is possible that the SWAN-based analysis is less impacted by sex biases in diagnostic criteria which cause females with ADHD to be underdiagnosed, leading to more similarity between the male and female groups.

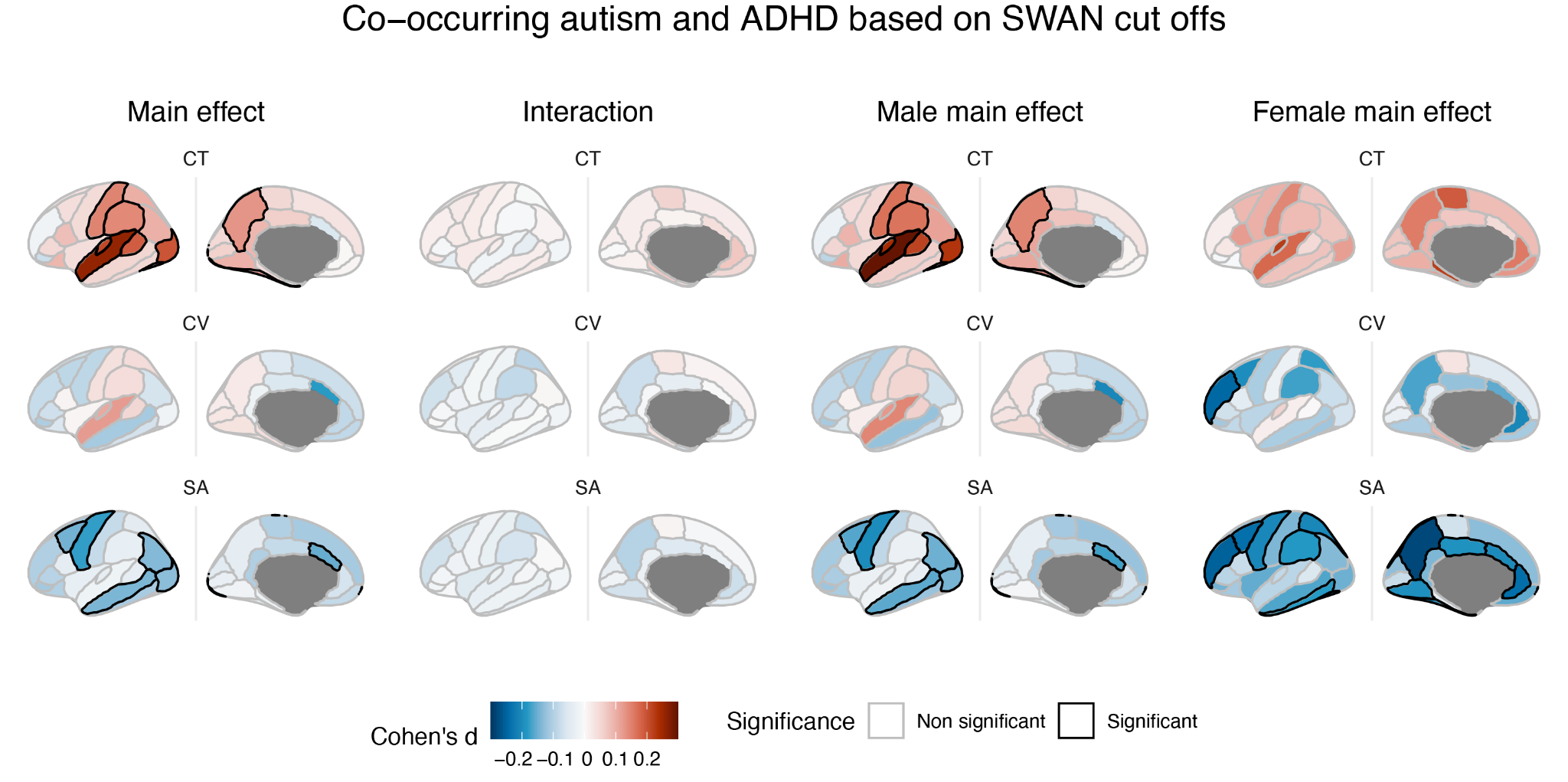

**Figure S8.3.** Cortical alterations (relative to controls) in individuals with co-occurring autism and ADHD based on the SWAN subscale cutoff. Main effects of diagnosis relative to controls; interaction with sex; main effects in males, and main effects in females. All maps show Cohen’s *d* effect sizes, with significant regions (passing 5% FDR) outlined in black. Red represents positive effect sizes (autism+ADHD > controls), and blue represents negative effect sizes (autism+ADHD < controls).

#### References

1.	Lai MC, Lombardo MV, Suckling J, Ruigrok ANV, Chakrabarti B, Ecker C, et al. Biological sex affects the neurobiology of autism. Brain. 2013;136: 2799–2815.

2.	Ecker C, Andrews DS, Gudbrandsen CM, Marquand AF, Ginestet CE, Daly EM, et al. Association Between the Probability of Autism Spectrum Disorder and Normative Sex-Related Phenotypic Diversity in Brain Structure. JAMA Psychiatry. 2017;74: 329.

3.	Zeestraten EA, Gudbrandsen MC, Daly EM, De Schotten MT, Catani M, Dell ’acqua F, et al. Sex differences in frontal lobe connectivity in adults with autism spectrum conditions. Transl Psychiatry. 2017;7: e1090.

4.	[R Core Team (2013) R A Language and Environment for Statistical Computing. R Foundation for Statistical Computing, Vienna. - References - Scientific Research Publishing. [cited 30 Jun 2023]. Available:](http://paperpile.com/b/jj3h1Z/HLkV) <https://www.scirp.org/(S(i43dyn45teexjx455qlt3d2q))/reference/ReferencesPapers.aspx?ReferenceID=1787696>

5.	Maechler M. Package “diptest.” 2022. Available: <https://cran.r-project.org/web/packages/diptest/diptest.pdf>

6.	Mowinckel AM, Vidal-Piñeiro D. Visualization of Brain Statistics With R Packages ggseg and ggseg3d. Advances in Methods and Practices in Psychological Science. 2020;3: 466–483.

7.	Forbes S. PupillometryR: An R package for preparing and analysing pupillometry data. J Open Source Softw. 2020;5: 2285.

8.	Lord C, Rutter M, Goode S, Heemsbergen J, Jordan H, Mawhood L, et al. Austism diagnostic observation schedule: A standardized observation of communicative and social behavior. J Autism Dev Disord. 1989;19: 185–212.

9.	Hus V, Gotham K, Lord C. Standardizing ADOS domain scores: Separating severity of social affect and restricted and repetitive behaviors. J Autism Dev Disord. 2014;44: 2400–2412.

10.	Hammill C, Lerch JP, Taylor MJ, Ameis SH, Chakravarty MM, Szatmari P, et al. Quantitative and Qualitative Sex Modulations in the Brain Anatomy of Autism. Biol Psychiatry Cogn Neurosci Neuroimaging. 2021;6: 898–909.

11.	Lam KSL, Aman MG. The repetitive behavior scale-revised: Independent validation in individuals with autism spectrum disorders. J Autism Dev Disord. 2007;37: 855–866.

12.	Bodfish JW, Symons FJ, Parker DE, Lewis MH. Varieties of repetitive behavior in autism: comparisons to mental retardation. J Autism Dev Disord. 2000;30: 237–243.

13.	Constantino JN, Davis SA, Todd RD, Schindler MK, Gross MM, Brophy SL, et al. Validation of a brief quantitative measure of autistic traits: comparison of the social responsiveness scale with the autism diagnostic interview-revised. J Autism Dev Disord. 2003;33: 427–433.

14.	Swanson JM, Schuck S, Porter MM, Carlson C, Hartman CA, Sergeant JA, et al. Categorical and Dimensional Definitions and Evaluations of Symptoms of ADHD: History of the SNAP and the SWAN Rating Scales. Int J Educ Psychol Assess. 2012;10: 51–70.

15.	Brites C, Salgado-Azoni CA, Ferreira TL, Lima RF, Ciasca SM. Development and applications of the SWAN rating scale for assessment of attention deficit hyperactivity disorder: a literature review. Braz J Med Biol Res. 2015;48: 965–972.

16.	Klein A, Tourville J. 101 labeled brain images and a consistent human cortical labeling protocol. Front Neurosci. 2012;6: 171.
